# Comparative analysis of anticholinergic burden scales to explain iatrogenic cognitive impairment and self-reported side effects in the euthymic phase of bipolar disorders: results from the FACE-BD cohort

**DOI:** 10.1101/2023.04.10.23288347

**Authors:** N Vidal, E Brunet-Gouet, S Frileux, B Aouizerate, V Aubin, R Belzeaux, P Courtet, T D’Amato, C Dubertret, B Etain, E Haffen, D Januel, M Leboyer, A Lefrere, PM Llorca, E Marlinge, E Olié, M Polosan, R Schwan, M Walter, the FACE-BD (FondaMental Academic Centers of Expertise for Bipolar Disorders) group, C Passerieux, P Roux

## Abstract

Bipolar disorders (BD) are characterized by cognitive impairment during the euthymic phase, to which treatments can contribute. The anticholinergic properties of medications, i.e., the ability of a treatment to inhibit cholinergic receptors, are associated with cognitive impairment in elderly patients and people with schizophrenia but this association has not been well characterized in individuals with remitted bipolar disorders. Moreover, the validity of anticholinergic burden scales designed to assess the anticholinergic load of medications has been scarcely tested in bipolar disorders. We aimed to test the concurrent validity of several scales by assessing their associations with objective cognitive impairment and subjective anticholinergic side-effects in BD. We hypothesized that the scale is valid if its association with cognitive impairment or self-reported anticholinergic side-effects is significant. A sample of 2,031 individuals with euthymic bipolar disorders was evaluated with a neuropsychological battery to identify cognitive impairment. Two scales among 27 were significantly positively associated with cognitive impairment in multiple logistic regressions, whereas chlorpromazine equivalents, lorazepam equivalents, the number of antipsychotics, or the number of treatments were not. The two scales significantly correlated with worse performance in processing speed and verbal memory. In addition, 14 scales showed good concurrent validity to assess self-reported peripheral anticholinergic side-effects and 13 were valid for evaluating self-reported central anticholinergic side-effects. Thus, we identified valid scales to monitor the anticholinergic burden in BD, which may be useful in assessing iatrogenic cognitive impairment in studies investigating cognition in BD.

## 1. Introduction

Type I and II bipolar disorders (BD) affect 1.06 and 1.57% of the population, respectively (Clemente et al., 2015) and can be associated with cognitive impairment in all phases of the illness (Cipriani et al., 2017). During manic and depressive episodes, bipolar patients show globally impaired verbal memory, language, attention, and executive functions (Cipriani et al., 2017; Ha et al., 2014; Kurtz & Gerraty, 2009). In the euthymic phase, patients also show impairment in all cognitive domains in 12.4 to 39.7% of cases (Burdick et al., 2014; Jensen et al., 2016; Roux et al., 2019). In BD, impaired cognition is associated with more frequent affective episodes (Sánchez-Morla et al., 2019), lower quality of life (Cotrena et al., 2016), and impaired social functioning (Baune & Malhi, 2015). Current management of BD consists largely of pharmacological interventions, and the use of highly anticholinergic drugs has increased over the past 25 years (Reinold et al., 2021; Sumukadas et al., 2014). However, the cognitive effects of medication in BD have been under-investigated. Psychotropic drugs are generally associated with cognitive improvement when they treat affective episodes (Kurtz & Gerraty, 2009) but may be associated with residual cognitive impairment during the euthymic period (Bourne et al., 2013; Roux et al., 2019; Xu et al., 2020). Anticholinergic properties of the treatment, which refer to its inhibitory activity on acetylcholine receptors, have been suspected of contributing to iatrogenic effects on cognition in healthy subjects (Mintzer & Griffiths, 2003). Such findings led to the design of anticholinergic burden scales to quantify the cumulative anticholinergic burden of treatment (Al Rihani et al., 2021). The association between the scores of such scales and cognitive impairment has been largely studied in populations with cognitive frailty, such as the elderly and individuals with schizophrenia (Lisibach et al., 2021; Georgiou et al., 2021), whereas it has only been characterized once in BD (Eum et al., 2017). Eum et al. reported the absence of a significant association between the Anticholinergic Drug Scale (ADS; Carnahan et al., 2006) and the composite score on a brief cognitive battery for 146 clinically stable patients with BD and psychotic features. The concurrent validity of other scales to evaluate cognitive impairment has never been tested in BD, despite large discrepancies between existing scales that are mainly due to the different methods used to design them (Rudd et al., 2005). For example, some scales were designed based exclusively on in vitro objective measures of serum anticholinergic activity of the drugs (Chew et al., 2008), whereas others were designed based on a literature review of the anticholinergic properties of the drugs discussed by experts (Rudolph et al., 2008). In addition, the same drug can be classified as highly anticholinergic by one scale and not anticholinergic by another: perphenazine is highly anticholinergic on the Anticholinergic Risk Scale (Rudolph et al., 2008) but is not anticholinergic on Chew’s scale (Chew et al., 2008). Although most recent scores are more convergent than older ones (Al Rihani et al., 2021), the considerable differences between scales argue for the need to identify the most optimal tools to assess the anticholinergic cognitive burden, specifically in BD. In addition to the cognitive burden, anticholinergic burden scales should monitor other anticholinergic side effects. Thus, this study also aimed to identify the scales with the best concurrent validity against common peripheral (such as dry mouth, constipation, and blurred vision) and central anticholinergic side effects (such as drowsiness and confusion) reported by individuals with BD.

## 2. Experimental procedures

The preregistration of the article is available at https://osf.io/zsrph/?view_only=753ecc167870464a8d88e0cd654e5085.

### 2.1. Study design and characteristics of the recruiting network

This multicenter transversal study included patients recruited into the FACE-BD (FondaMental Advanced Centers of Expertise for Bipolar Disorders) cohort within a French national network of 10 centers (Bordeaux, Colombes, Créteil, Grenoble, Marseille, Monaco, Montpellier, Nancy, Paris, and Versailles). This network was created by the Fondation FondaMental (https://www.fondation-fondamental.org), which organized and provided the necessary resources to follow cohorts of patients with BD, promoting comparative research. The local ethics committee approved the study (Comité de Protection des Personnes Ile de France IX) on January 18, 2010, under French law for non-interventional studies (observational studies without any risk, constraint, supplementary or unusual procedure concerning diagnosis, treatment, or monitoring). The board required that all patients be given an informational letter but waived the requirement for written informed consent. However, verbal consent was witnessed and formally recorded.

### 2.2. Participants

The diagnosis of BD was based on the Structured Clinical Interview for DSM-IV-TR (SCID) criteria (First et al., 1997). Outpatients between 18 and 65 years of age with type I, II, or not otherwise specified (NOS) BD were eligible for the present study. We selected patients who were euthymic at the time of testing according to the DSM-IV-R criteria and who had scores on the Montgomery Asberg Depression Rating Scale (MADRS; Montgomery and Asberg, 1979) ≤10 and the Young Mania Rating Scale (YMRS; Young et al., 1978) ≤12. We excluded non-euthymic participants, as previous studies reported that mood symptoms may influence cognitive performance (Kurtz & Gerraty, 2009) and self-reported side effects (Gao et al., 2008; Vaccarino et al., 2009) beyond the iatrogenic load.

We also excluded patients with a history of neurological disorders, dyslexia, dyscalculia, dysphasia, dysorthographia, dyspraxia, any episode of substance abuse disorder in the previous months, or electro-convulsive therapy in the past year, again to remove other sources of cognitive impairment other than medications.

### 2.3. Measures

#### 2.3.1. Anticholinergic burden scales

We identified 36 available scales in a literature review performed in November 2022 (see Supplementary Information for more details). Among the 36 scales, we excluded seven: the exclusion rules and the description of the selected scales are reported in Supplementary Information.

Drugs not included in a scale were scored 0, as if they had no anticholinergic properties according to the scale, as in a previous study (Lisibach et al., 2022a). Two methods to compute the total anticholinergic burden of the treatment were used: summing the scores (“sum”), as recommended by Carnahan et al., (2006), and using the highest score (“max”), as recommended by Sittironnarit et al. (2011).

#### 2.3.2. Cognition

Experienced neuropsychologists administered the tests in a fixed order that was the same for every center. Testing lasted approximately 120 min, including 5-to-10-min breaks. The standardized test battery complied with the recommendations of the International Society for Bipolar Disorders (Yatham et al., 2010) and included 11 tests, including five subtests from the Wechsler Adult Intelligence Scale version III (WAIS III) (Wechsler, 1997a) or version IV (Wechsler et al., 2008), as the French version of the WAIS-IV started to be used in the FACE-BD cohort as it became available. The battery evaluated six domains:

- Processing speed: Digit symbol coding (WAIS-III) or coding (WAIS-IV), WAIS symbol search, and Trail Making Test, part A (Reitan, 1958)
- Verbal memory: California Verbal Learning Test short and long delay-free recall and total recognition (Delis, 2000)
- Attention: Conners’ Continuous Performance Test II V.5 (omission, commission, variability, and detectability) (Conners and Staff, 2000)
- Working memory: WAIS digit span (total score) and spatial span (forward and backward scores) from the Wechsler Memory Scale version III (Wechsler, 1997b)
- Executive function: color/word condition of the Stroop test (Golden, 1978), semantic and phonemic verbal fluency (Lezak, 2004), and Trail Making Test, part B (Reitan, 1958)
- Verbal and perceptual reasoning: WAIS vocabulary and matrices

Higher scores reflect better performance. We converted the raw scores of each test to standardized scores based on normative data (Conners & Staff, 2000; Godefroy, 2008; Golden, 1978; Poitrenaud et al., 2007). We computed a global deficit score or GDS (Heaton et al., 2004) to determine a threshold for cognitive impairment (see Supplementary information for details about the computation method). We also computed a mean z-score for each cognitive domain.

#### 2.3.3. Drug side effects

Adverse effects were assessed using the Patient-Rated Inventory of Side Effects Modified (PRISE-M; Rush et al., 2004; Rush & O’Neal, 1999). The PRISE-M evaluates the tolerance level for 32 side effects in nine domains (gastro-intestinal, cardiovascular, skin, nervous, sensory systems, urogenital systems, sleep, sexual function, and others). Each item was coded 0 for absent, 1 for bearable, and 2 for painful. We computed two scores for anticholinergic side effects based on the usual anticholinergic side effects reported in the literature (Feinberg, 1993; Fond et al., 2019; Giuliano & Droupy, 2013; Lieberman, 2004). The peripheral score was the sum of the following item scores: “dry mouth”, “constipation”, “blurred vision”, “dry skin”, “itching”, “difficulty urinating”, “frequent urination”, “erectile dysfunction”, and “orgasm disorders”. The central score was the sum of the scores of “increased sleep time”, “loss of energy”, “asthenia”, “impaired concentration”, “general malaise”, “reduced sex drive”, and “orgasm disorders”.

#### 2.3.4. Clinical covariates and alternative measures of iatrogenic cognitive burden

Socio-demographic (sex, age, education level) and clinical variables, such as the age at onset of BD, the total number of mood episodes, the subtype of BD, and a history of psychotic symptoms were recorded. Mania was evaluated by the Young Mania Rating Scale (YMRS), depression by the Montgomery Asberg Depression Rating Scale (MADRS), and symptoms severity by the Clinical Global Impressions scale (CGI; Guy, 1976). We also recorded the class of treatment (antidepressants, anticonvulsants, lithium, antipsychotics, anxiolytics, and antiparkinsonian drugs prescribed for extrapyramidal side effects). These variables were all screened as potential covariates of the association of anticholinergic load with cognitive deficit, as well as with peripheral and central anticholinergic side effects. We additionally recorded whether a patient used multiple classes of treatment, i.e., two or more different classes, for information.

We collected alternative measures associated with iatrogenic cognitive burden: the number of medications (reported in Dias et al., 2012), the number of antipsychotics (reported in Bourne et al., 2013), chlorpromazine (reported in Jamrozinski et al., 2009) equivalents (CPZeq, computed from the formulas proposed by Andreasen et al., 2010 and Leucht et al., 2015), and lorazepam (reported in Savić et al., 2021) equivalents (computed from the formulas proposed by Kane, 2017). As for the anticholinergic burden scales, we assessed whether these measures were associated with cognitive impairment.

### 2.4. Statistical analysis

Statistical analyses were carried out using R. For the multiple analyses, missing data were estimated using multivariate imputation by chained equations (50 imputations, *mice* package of R, Van buuren & Groothuis-Oeudshoorn, 2011). Each variable had < 30% missing data, allowing the use of multiple imputations (Marshall et al., 2010). We compared imputed and non-imputed datasets to ensure the validity of the imputed values (Nguyen et al., 2017). The fraction of missing information (fmi) computed by the *pool* function of the *mice* package is reported in the results.

First, we assessed associations between the various anticholinergic scale scores and the presence of a cognitive deficit using successive bivariable logistic regressions on the complete cases.

For scales associated with a cognitive deficit at a 0.05 significance level in bivariable analyses, we ran successive multiple logistic regressions of the scores on the presence of a cognitive deficit that included a subset of covariates. Covariates were included in the model if they were associated with the scale validated by the most studies, i.e., the Anticholinergic Cognitive Burden scale (Boustani et al., 2008; Lisibach et al., 2021), with a p-value < 0.2, as pre-established in the preregistration. We also ran multiple logistic regression analyses using the four alternative measures of iatrogenic cognitive burden (number of medications, number of antipsychotics, chlorpromazine equivalents, and lorazepam equivalents) with the same set of covariates.

We then conducted receiver operating characteristic (ROC) curve analysis for scales associated with cognitive impairment in the multiple analysis at a 0.05 significance level. The area under the curve (AUC) was interpreted as follows: 0.5: unpredictive, 0.5-0.7: poorly predictive, 0.7-0.9: moderately predictive, > 0.9: strongly predictive.

In addition, we computed the correlations (Spearman *ρ*) between the scales significantly associated with a cognitive deficit in multiple regression models and the z-scores in each cognitive domain to explore whether the scales are sensitive to specific profiles of cognitive impairment. For the correlation analysis, we applied the false-discovery procedure of Benjamini-Hochberg to p-values, with the significance level set to 0.05 (Benjamini & Hochberg, 1995).

We also assessed the associations between the scales and peripheral and central anticholinergic side-effects scores using the same analytical framework as above but using linear regressions.

Finally, we computed the correlations (Spearman *ρ*) between the scales significantly associated with peripheral or central anticholinergic side-effect subscores in multiple regression models and each item of the PRISE-M to explore which self-reported side effects were associated with the scales and whether the scales were sensitive to non-anticholinergic adverse effects. Similarly, we applied the false-discovery procedure of Benjamini-Hochberg to p-values, with the significance level set to 0.05 (Benjamini & Hochberg, 1995).

## 3. Results

### 3.1. Description of the sample

We included 2,031 participants with euthymic bipolar disorders (62% female), among whom 393 (19%) were considered to be cognitively impaired by the GDS criterion (**Table 1**). The characteristics of the treatments are presented in **Table 2**. The anticholinergic scales reported between 1.13% and 67.01% patients with a non-zero score, i.e., with an anticholinergic burden, indicating a large discrepancy between scales (**Supplementary Figure 2**).

**Table 1.** Description of the sample (n=2031).

| Category | *n* | *Mean (SD)* | *% missing data* |
| --- | --- | --- | --- |
| Female, n (%) | 1254 (62%) |  | 1% |
| Age (mean, ±SD) | | 39.6 (12.3) | 0% |
| Education level (years) |  | 14.4 (2.6) | 6% |
| Cognitive impairment, n (%) | 393 (19%) |  | 15% |
| MADRS (mean, ±SD) | | 4.13 (3.22) | 4% |
| YMRS (mean, ±SD) | | 1.70 (2.65) | 4% |
| CGI (mean, ±SD) | | 2.6 (1.4) | 6% |
| Type I BD, n (%) | 1050 (52%) |  | 0% |
| Type II BD, n (%) | 790 (39%) |  | 0% |
| Unspecified BD, n (%) | 191 (9%) |  | 0% |
| End of the last characterized mood episode > 3 months before, n (%) | 1589 (78%) |  | 10% |
| Age at the first mood episode (mean, ±SD) | | 23.9 (9.2) | 10% |
| Number of depressive episodes (mean, ±SD) | | 5.4 (6.4) | 20% |
| Number of manic episodes (mean, ±SD) | | 1.2 (2.1) | 7% |
| Number of hypomanic episodes (mean, ±SD) | | 3.4 (6.1) | 29% |
| Number of mixed episodes (mean, ±SD) | | 0.4 (1.5) | 22% |
| Patients with history of psychosis, n (%) | 724 (36%) |  | 18% |

MADRS: Montgomery-Asberg Depression Rating Scale
YMRS: Young Mania Rating Scale
CGI: Clinical Global Impressions scale.

**Table 2.** Characteristics of the treatments (n=2031).

| Variable | *n* | *Mean (SD)* | *% missing data* |
| --- | --- | --- | --- |
| Number of medications |  | 2 (1) | 30% |
| Number of antipsychotics |  | 0.5 (0.6) | 30% |
| Number of patients using antidepressants, n (%) | 492 (34%) |  | 30% |
| Number of patients using anticonvulsant, n (%) | 702 (49%) |  | 30% |
| Number of patients using lithium, n (%) | 507 (36%) |  | 30% |
| Number of patients using antipsychotic, n (%) | 617 (43%) |  | 30% |
| Number of patients using anxiolytic, n (%) | 344 (24%) |  | 30% |
| Number of patients using antiparkinsonian drug, n (%) | 22 (2%) |  | 30% |
| Number of patients using multiple classes, n (%) | 456 (32%) |  | 30% |
| Chlorpromazine equivalent, mg/24h (mean, ±SD) | | 103.6 (183.8) | 31% |
| Lorazepam equivalent, mg/24h (mean, ±SD) | | 0.11 (0.58) | 30% |

### 3.2. Cognitive impairment and anticholinergic burden scales

The bivariable regressions of cognitive impairment, defined by the GDS criterion, on the anticholinergic burden scales are reported in **Figure 1**. The values of 21 scales were significantly positively associated with cognitive impairment when using the “sum” method (1.06 ≤ OR ≤ 1.69) and the “max” method (1.06 ≤ OR ≤ 3.21). We then adjusted the bivariable models of the 21 scales with a set of covariates (see the covariate selection process in Supplementary Information). The values from two of the 21 scales were significantly associated with cognitive impairment after inclusion of the covariates: Chew’s scale (Chew et al., 2008), computed using the “max” method, and the Anticholinergic Toxicity Scale (ATS; Xu et al., 2017), computed using the “sum” and “max” methods (**Table 3**, see **Supplementary Table 2** for the results of the other 21 scales). An increase of 1 point in the maximum Chew’s score or the Anticholinergic Toxicity Scale was associated with a 33% or 11 to 14% increase, respectively, in the risk of being cognitively impaired. The number of medications, antipsychotics, CPZeq, and lorazepam equivalents were not significantly associated with cognitive impairment after adjusting on the same set of covariates (**Supplementary Table 3**). Among the covariates, the use of lithium was significantly associated with more frequent cognitive impairment (1.62 ≤ OR ≤ 1.98) and education level was significantly associated with less frequent cognitive impairment (OR = 0.87) in all multiple regression models. The use of antipsychotics and antiparkinsonian drugs was significantly associated with more frequent cognitive impairment in 96% and 52% of the multiple regression models, respectively (1.47 ≤ OR ≤ 1.68 and 2.58 ≤ OR ≤ 2.78, respectively). The AUC for Chew’s scale and the Anticholinergic Toxicity Scale was 0.57 and 0.53, respectively (**Figure 2**), thus suggesting poor predictability (0.5 < AUC < 0.7) of cognitive impairment by the two scales at an individual level.

**Figure 1.**
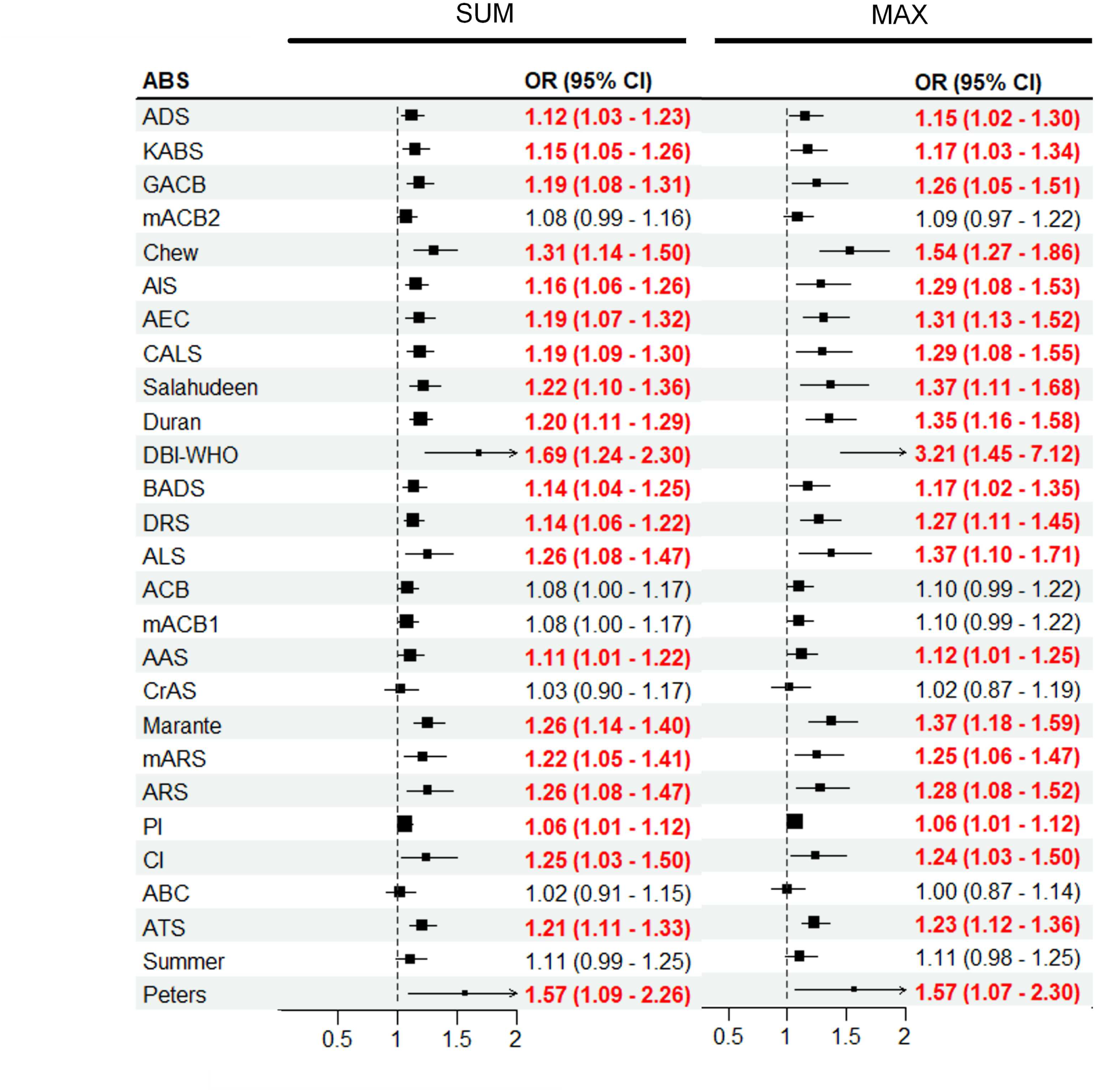
Results of the bivariable logistic regression models of cognitive impairment with the scales as the predictor. Significant (p < 0.05) odds ratios are shown in red. The total anticholinergic burden score was computed by either summing the scores of each treatment (SUM) or by using the maximum score (MAX).

**Figure 2.**
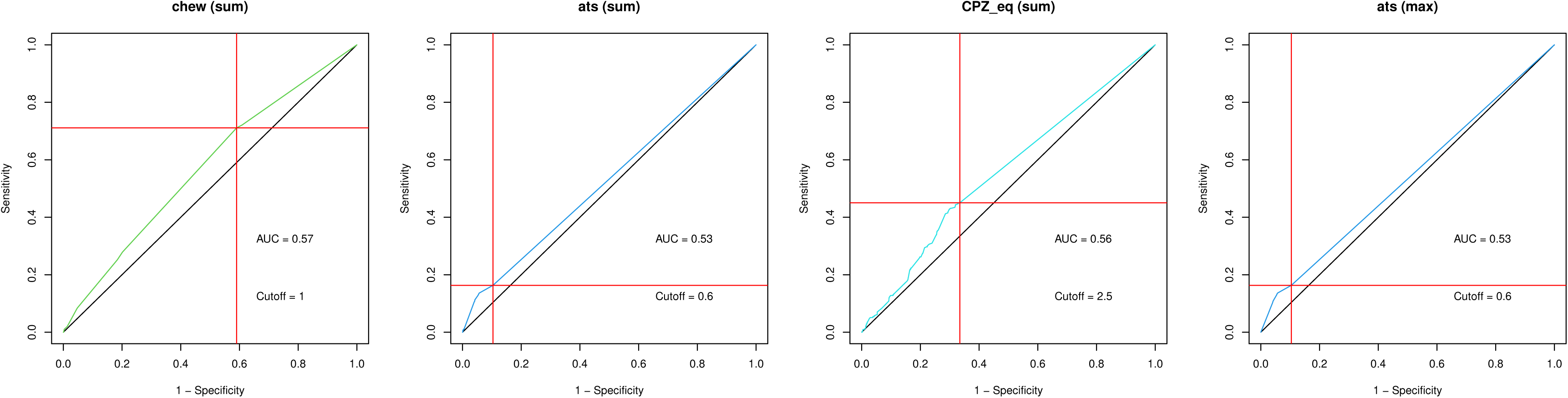
ROC curves for Chew’s scale, the Anticholinergic Toxicity Scale, and chlorpromazine equivalents as classifiers of cognitive impairment. |Cutoffs were computed using Unal’s guidelines (2017): the cutoff represents the value for which the sensitivity and specificity are the closest to the value of the area under the ROC curve such that the absolute value of the difference between the sensitivity and specificity values is minimum. Red lines indicate the specificity and sensitivity at the cutoff. ats: Anticholinergic Toxicity Scale, AUC: area under the curve, chew: Chew’s Scale, CPZ_eq: chlorpromazine equivalents.

**Table 3.** Multiple logistic regression models of cognitive impairment with Chew’s scale and the Anticholinergic Toxicity Scale as main predictors.

|  | Chew (max) |  |  | ATS (sum) |  |  | ATS (max) |  |  |
| --- | --- | --- | --- | --- | --- | --- | --- | --- | --- |
|  | OR (95% CI) | p-value | fmi | OR (95% CI) | p-value | fmi | OR (95% CI) | p-value | fmi |
| Scale | **1.33 (1.06-1.67)** | **0.013** | 0.12 | **1.11 (1.01-1.23)** | **0.039** | 0.14 | **1.13 (1.01-1.27)** | **0.027** | 0.20 |
| MADRS | 0.98 (0.94-1.03) | 0.41 | 0.12 | 0.98 (0.94-1.02) | 0.35 | 0.07 | 0.98 (0.93-1.02) | 0.291 | 0.12 |
| History of psychosis | 0.93 (0.67-1.29) | 0.673 | 0.21 | 0.9 (0.64-1.28) | 0.562 | 0.30 | 0.91 (0.65-1.26) | 0.567 | 0.21 |
| Antidepressants | 0.76 (0.54-1.05) | 0.095 | 0.15 | 0.82 (0.6-1.12) | 0.21 | 0.14 | 0.83 (0.61-1.14) | 0.25 | 0.16 |
| Lithium | **1.66 (1.21-2.29)** | **0.002** | 0.16 | **1.88 (1.4-2.51)** | **<.001** | 0.15 | **1.93 (1.44-2.59)** | **<.001** | 0.14 |
| Antipsychotics | **1.51 (1.11-2.04)** | **0.008** | 0.13 | **1.55 (1.15-2.07)** | **0.004** | 0.10 | **1.56 (1.16-2.09)** | **0.003** | 0.11 |
| Anxiolytics | 1.26 (0.9-1.74) | 0.176 | 0.18 | 1.21 (0.87-1.68) | 0.249 | 0.15 | 1.22 (0.88-1.69) | 0.24 | 0.18 |
| Antiparkinsonian | **2.67 (1.05-6.79)** | **0.039** | 0.06 | 2.11 (0.8-5.56) | 0.131 | 0.07 | 2.19 (0.85-5.67) | 0.105 | 0.07 |
| Education level | **0.87 (0.82-0.91)** | **<.001** | 0.12 | **0.87 (0.82-0.91)** | **<.001** | 0.13 | **0.86 (0.82-0.91)** | **<.001** | 0.12 |
| Age at first episode | 1 (0.99-1.02) | 0.808 | 0.28 | 1 (0.99-1.02) | 0.641 | 0.22 | 1 (0.99-1.02) | 0.775 | 0.29 |
| Number of manic episode | 1.06 (0.99-1.14) | 0.086 | 0.21 | 1.07 (0.99-1.14) | 0.082 | 0.23 | 1.06 (0.99-1.14) | 0.086 | 0.21 |
| Number of depressive episode | 1 (0.98-1.03) | 0.757 | 0.27 | 1 (0.98-1.03) | 0.85 | 0.34 | 1 (0.98-1.03) | 0.832 | 0.28 |
| CGI | 1.1 (0.99-1.21) | 0.065 | 0.16 | 1.09 (0.99-1.21) | 0.093 | 0.21 | 1.1 (0.99-1.21) | 0.071 | 0.17 |

Significant associations are in bold. Chew: Chew's scale (Chew et al., 2008), ATS: Anticholinergic Toxicity Scale (Xu et al., 2017), fmi: fraction of missing information.

### 3.3. Associations between cognitive domains and anticholinergic burden

After identifying two valid scales to measure the anticholinergic cognitive burden, we calculated pairwise correlations between the two scales and the patients’ z-scores in each of the six cognitive domains (processing speed, verbal memory, attention, working memory, executive function, and reasoning). Chew’s score was significantly but weakly associated with lower performance in processing speed (*ρ* = -0.08), verbal memory (*ρ* = -0.09), attention (*ρ* = -0.11), and executive function (*ρ* = -0.12) (**Figure 3**). The Anticholinergic Toxicity Scale was significantly but weakly associated with worse performance in processing speed (*ρ* = -0.08) and verbal memory (*ρ* = -0.04).

**Figure 3.**
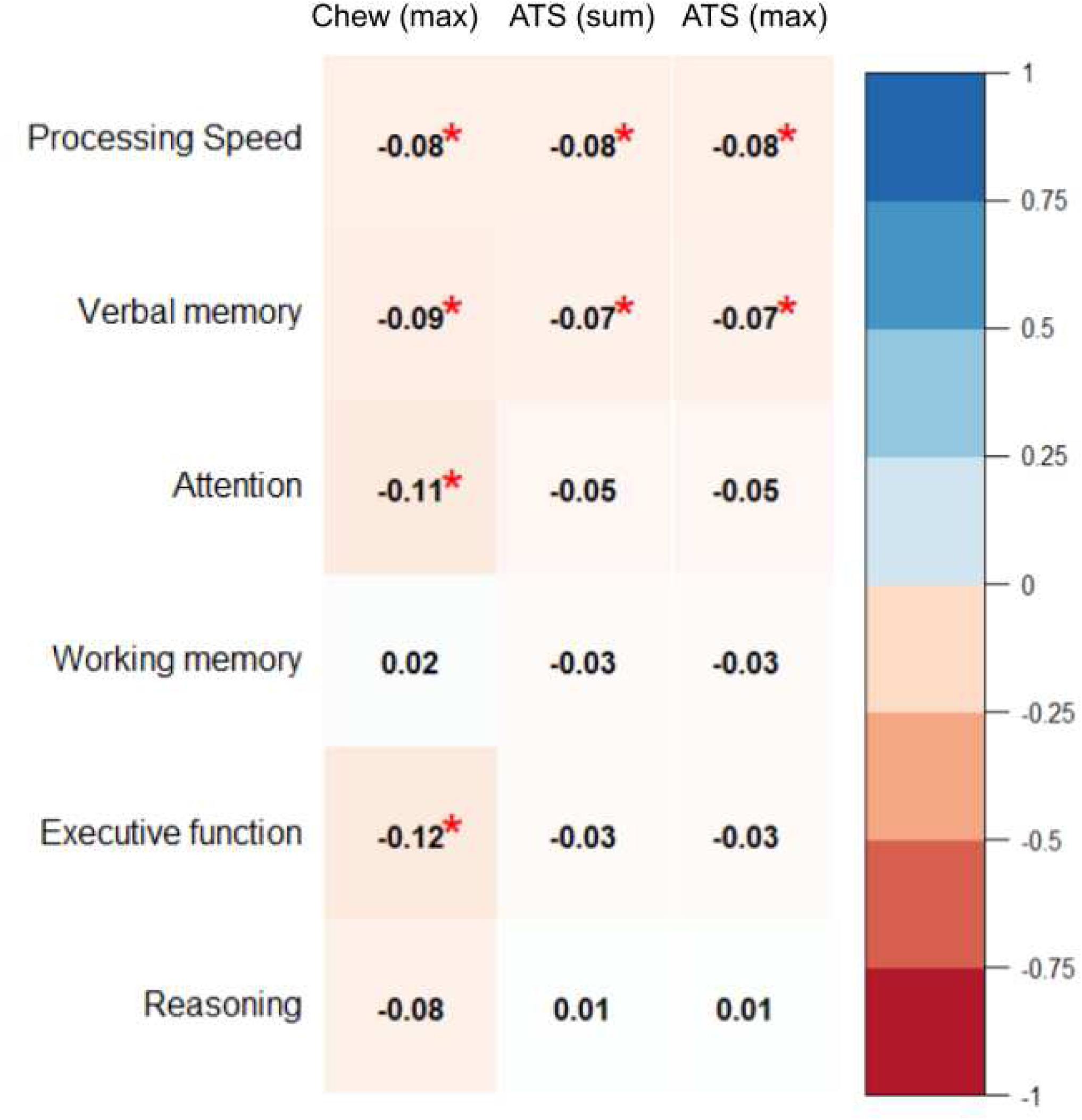
Pairwise correlations (Spearman coefficients) between z-scores in six cognitive domains and Chew’s scale and the Anticholinergic Toxicity Scale. A star indicates significance at the 5% level after Benjamini-Hochberg/false-discovery rate correction. Chew: Chew’s Scale; ATS: Anticholinergic Toxicity Scale.

### 3.4. Self-reported side effects and anticholinergic burden scales

Among the 27 scales, 21 were significantly associated with the subscore for peripheral anticholinergic side effects (Standardized β: 0.055 ≤ β≤ 0.158; 0.003 ≤ R^2^ ≤ 0.025) and 22 were significantly associated with the subscore for central anticholinergic side effects (Standardized β: 0.058 ≤ β ≤ 0.190; 0.003 ≤ R^2^ ≤ 0.036) in bivariable logistic regression models (**Supplementary Figure 3**). We then adjusted the 21 models for the peripheral subscore and the 22 for the central subscore with the same set of previously selected covariates. Fourteen scales were significantly associated with the peripheral subscore in multiple logistic regression models when using the “max” method, the “sum” method, or both (Standardized β: 0.032 ≤ β ≤ 0.157; 0.09 ≤ R^2^ ≤ 0.10) (**Supplementary Table 4**): the Anticholinergic Burden Classification (Ancelin et al., 2006), the Anticholinergic Risk Scale (Rudolph et al., 2008), the Anticholinergic Cognitive Burden scale (Boustani et al., 2008), the Anticholinergic Impregnation Scale (Briet et al., 2017), the Brazilian Anticholinergic Activity Drug Scale (Nery & Reis, 2019), the CRIDECO Anticholinergic Load Scale (Ramos et al., 2022), the Delirogenic Risk Scale (Hefner et al., 2015), Durán’s scale (Durán et al., 2013), the German Anticholinergic Burden Scale (Kiesel et al., 2018), the Korean Anticholinergic Burden Scale (Jun et al., 2019), the Muscarinic Acetylcholinergic Receptor ANTagonist Exposure Scale (Klamer et al., 2017), the modified Anticholinergic Cognitive Burden Scale of Joshi et al. (2021), the modified Anticholinergic Risk Scale of Sumukadas et al. (2016), and Salahudeen’s scale (Salahudeen et al., 2015). Thirteen scales were significantly associated with the central subscore in multiple logistic regression models (Standardized β: 0.039 ≤ β ≤ 0.153; 0.21 ≤ R^2^ ≤ 0.22) (**Supplementary Table 5**) (a list of the 13 scales is available in Supplementary Information). Pairwise correlations between the scales were significantly associated with one of the two scores in multiple regression analyses and each item of the PRISE-M is reported in **Figure 4**. All scales were significantly associated with an increase in patient-reported dry mouth (0.08 ≤ *ρ* ≤ 0.16) and weight gain (0.08 ≤ *ρ* ≤ 0.17). More than 12/15 scales were significantly associated with increased sleep time, more constipation, asthenia, loss of energy, and reduced sex drive. Certain scales were also significantly positively associated with side effects not included in the peripheral or central anticholinergic subscores, such as diarrhea, palpitations, tinnitus, tremor, dizziness, vertigo, anxiety, and agitation.

**Figure 4.**
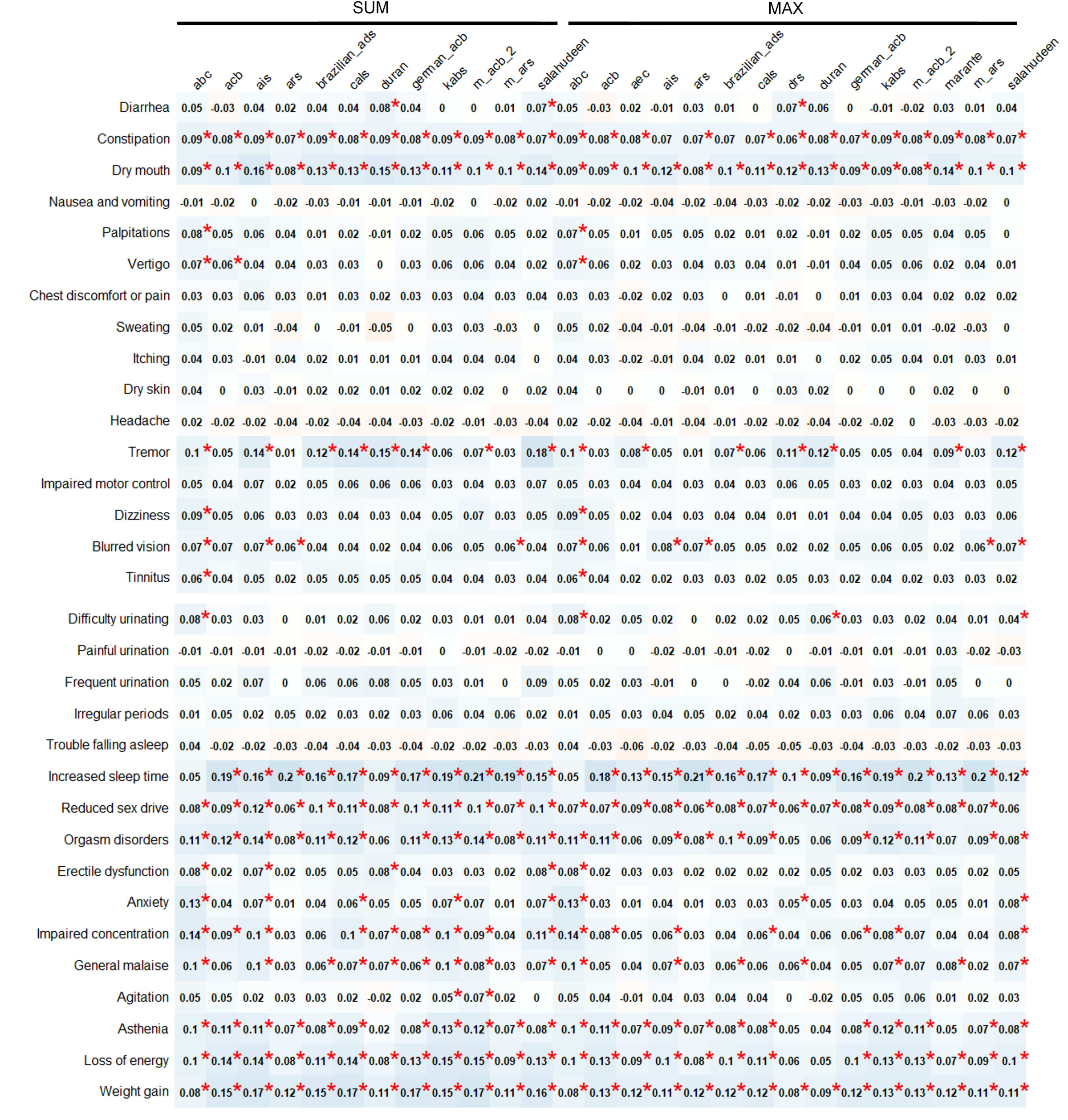
Pairwise correlations (Spearman coefficients) between items of the PRISE-M questionnaire and 15 anticholinergic burden scales. Colors indicate the strength and valence of the correlation. A star indicates significance at the 5% level after Benjamini-Hochberg/false-discovery rate correction. abc: Anticholinergic Burden Classification; acb: Anticholinergic Cognitive Burden Scale; aec: Anticholinergic Effect on Cognition; ais: Anticholinergic Impregnation Scale; ars: Anticholinergic Risk Scale; brazilian_ads: Brazilian Anticholinergic Activity Drug Scale; cals: the CRIDECO Anticholinergic Load Scale; drs: Delirogenic Risk Scale; duran: Durán’s Scale; german_acb: German Anticholinergic Burden Scale; kabs: Korean Anticholinergic Burden Scale; m_acb_2: Modified Anticholinergic Cognitive Burden Scale 2; marante: Muscarinic Acetylcholinergic Receptor ANTagonist Exposure Scale; m_ars: Modified Anticholinergic Risk Scale; salahudeen: Salahudeen’s Scale.

## 4. Discussion

We tested the concurrent validity of 27 existing anticholinergic scales against cognitive impairment and self-reported anticholinergic side effects of medication in a large sample of individuals with bipolar disorders who were euthymic at the time of testing. We found marked discrepancies in the number of drugs with anticholinergic properties listed in each scale, as reported in a previous study that included 11 (Al Rihani et al., 2021) of the 27 scales used in the present study.

We identified two scales with acceptable concurrent validity to assess cognitive impairment: Chew’s scale using the “max” method and the Anticholinergic Toxicity Scale using either the “max” or “sum” method. Chew’s scale and the Anticholinergic Toxicity Scale were more robust indicators of cognitive impairment than antipsychotic load measured by chlorpromazine equivalents, which is associated with poorer performance in attention in BD (Jamrozinski et al., 2009). We replicated the lack of association between cognitive impairment in bipolar patients and Anticholinergic Drug Scale values reported by Eum et al. (2017). Previous use of Chew’s scale showed its association with cognitive impairment in elderly patients (Lampela et al., 2013; Lisibach et al., 2021), whereas the Anticholinergic Toxicity Scale has been shown to be strongly associated with delirium and mortality in elderly patients (Lisibach et al., 2022a & 2022b), but no study has explored their association with cognitive performance for individuals with psychiatric disorders. Indeed, Chew’s scale was designed based exclusively on in vitro measures of serum anticholinergic activity and the Anticholinergic Toxicity Scale was designed based on the affinity for muscarinic receptors deduced from the molecular structure of the drug: neither of the two scales were designed based on clinical observations, making our results quite unexpected. Our results suggest that completely objective scales based on computational or in vitro receptor affinities might be better indicators of cognitive impairment than other scales that account for patients’ reports of anticholinergic side effects or expert opinions. In addition, the use of completely objective scales provides for interesting perspectives for research. Indeed, new pharmacological agents could be positioned on the scale from the earliest phases of their development, before pharmacovigilance and expert consensus. Despite significant associations of Chew’s scale and the Anticholinergic Toxicity Scale with cognitive impairment, ROC curve analyses showed them to be insufficiently strong to establish a threshold for the risk of iatrogenic cognitive impairment at the individual level, which would be useful in selecting participants for interventional studies targeting cognitive improvement in BD. However, we recommend the use of these scales for studies that require controlling for the burden of medication on cognition at the group level. These scales may also be useful in clinical settings to anticipate the cognitive impact of medication changes.

Beyond the anticholinergic burden, we identified other iatrogenic sources of cognitive impairment in BD, such as the use of lithium and antipsychotics. Other cross-sectional studies reported a weak and inconsistent negative association between cognition and the use of lithium in euthymic BD (Wingo et al., 2009). However, it was not possible to conclude a causal link between the use of antipsychotics or lithium and cognitive impairment, as we did not investigate the dose effect of these medications on cognition. Longitudinal studies are needed to clarify the effect of antipsychotics and lithium on cognition in BD and should take into account the daily dose and serum levels, duration of exposure, and therapeutic response. Any decision to discontinue antipsychotics or lithium due to cognitive side effects should be made with care after a careful clinical medication review and evaluation of the benefit-to-risk ratio.

Considering each cognitive domain separately, Chew’s scale and the Anticholinergic Toxicity Scale were associated with worse performance in processing speed and verbal memory, albeit weakly. Moreover, Chew’s scale was associated with worse performance in executive function and attention. This is the first evidence of an association between anticholinergic burden and executive function and verbal memory in BD. However, executive function and verbal memory are globally impaired in BD and associated with poor functional outcomes, hospitalization, and suicide attempts (Martínez-Arán et al., 2004; Mur et al., 2007). Our results advocate for assessment of the anticholinergic burden of treated euthymic bipolar patients.

In addition, we identified 14 scales with good concurrent validity against peripheral anticholinergic side effects and 13 with good concurrent validity against central anticholinergic side effects in BD. The 15 scales with good validity for at least one of the scores correlated, albeit weakly, with common patient-reported peripheral (such as dry mouth) and central (loss of energy, increased sleep time, and reduced sex drive) anticholinergic side effects. Furthermore, the same scales were all significantly associated with weight gain, which is likely not attributed to anticholinergic properties but to other effects of antipsychotics, mood stabilizers, and antidepressants, which score high on these scales (Vanina et al., 2002). Certain scales were also associated with diarrhea, tremor, vertigo, anxiety, and agitation, which are not generally considered to be anticholinergic side effects. These results suggest that these scales may lack divergent validity, as they were associated with non-anticholinergic side-effects. These unexpected associations may arise from the fact that the PRISE-M is a subjective scale that refers to the tolerance of the patients for what they consider to be drug-induced side-effects. Patients may have mistakenly considered a symptom of their disorder, such as agitation, as a side-effect of the treatment. Considering that PRISE-M items are negatively related with self-esteem and psychosocial functioning in resistant depression (Levy et al., 2021), reducing polypharmacy and minimizing the score of the scales associated with peripheral or central subscores could improve the anticholinergic tolerability of psychotropic treatments and patient functioning.

The main limitation of our study was the cross-sectional design. Although we controlled for a history of psychosis, the subtype of BD, and the severity of the disorder, our results could be confounded by the indication of the treatment. Cognitively impaired patients may have more anticholinergic treatment than patients with no cognitive impairment because they experience more intense bipolar symptoms or are more regularly hospitalized. Finally, we limited the study to one definition of cognitive impairment, which might have been restrictive. However, using a conservative criterion such as the GDS enabled us to identify the most cognitively impaired patients, i.e., the most challenging patients to treat.

## Conclusion

The current management of BD consists largely of pharmacological intervention and our results suggest that the anticholinergic burden should be monitored in BD. The variety of available anticholinergic burden scales is considerable, and we recommend two scales to help identify individuals with remitted BD at risk of iatrogenic cognitive impairment: Chew’s scale and the Anticholinergic Toxicity Scale. In addition, 14 scales showed concurrent validity with self-reported peripheral anticholinergic side effects and 13 showed good validity for measuring self-reported central anticholinergic side effects. The use of anticholinergic burden scales appears to be appropriate for guiding clinicians and researchers in measuring the iatrogenic effects of prescribed treatments.

## Data Availability

All data produced in the present study are available upon reasonable request to the authors

