## Supplementary Material for "Comparative analysis of anticholinergic burden scales to explain iatrogenic cognitive impairment and self-reported side effects in the euthymic phase of bipolar disorders: results from the FACE-BD cohort"

Supplementary information

### 1. Measures

#### 1.2. Anticholinergic burden scales

We identified scales by conducting a literature review in Google Scholar, PubMed, and Cochrane in November 2022 with the following keywords:
("anticholinergic" [Title/Abstract]) OR ("cholinergic antagonists" [Title/Abstract]) OR ("antimuscarinic" [Title/Abstract]) OR ("muscarinic antagonists" [Title/Abstract]) OR ("atropinic" [Title/Abstract]) AND ("scale"[Title/Abstract]) OR ("measure"[Title/Abstract]) OR ("tool"[Title/Abstract])) OR ("score"[Title/Abstract])

We used the references of the articles and the citations to the articles to identify additional scales according to the “snowballing” procedure. We found 36 scales designed to assess, at least partially, the anticholinergic properties of a treatment (**Supplementary Figure 1**).

**Excluded scales:**
We excluded Cancelli’s anticholinergic burden score because it excluded antipsychotics (Cancelli et al., 2007) and Tune’s list because the authors did not report any score (Tune et al., 1992). We also excluded the Clinician-Rated Anticholinergic Score (the CR-ACh; Han et al., 2001) and the modified Clinician-Rated Anticholinergic Score (the CR-ACh-mod; Carnahan et al., 2002) because we favored the most recent versions of these scales: the Clinician-Rated Anticholinergic score (CrAS; Han et al., 2008) and the Anticholinergic Drug Scale (ADS; Carnahan et al., 2006), respectively. We excluded the Drug Delirium Scale (DDS; Nguyen et al., 2016) because the reported score is not based exclusively on the anticholinergic properties of the drug. Finally, we excluded the Drug Burden Index (DBI; Hilmer et al., 2007) and Cao’s score (Cao et al., 2008), which introduced an equation to compute anticholinergic load. We favored the version of the equation of Dauphinot et al. (2014), called the DBI-WHO, which relies on the defined daily dose of the WHO instead of the US FDA dosage norms, thus suggesting better generalizability of this version of the score. Among the 29 remaining scales, two were unavailable upon request to the authors (Aizenberg et al., 2002; Whalley et al., 2012).

**Included scales:**
Among the 27 scales, 23 were numerical. Nineteen numerical scales were discrete:

- **Summers**’s scale (Summers, 1978)
- Anticholinergic Drug Scale (**ADS**; Carnahan et al., 2006, updated in 2021 by the authors)
- Anticholinergic Burden Classification (**ABC**; Ancelin et al., 2006)
- Clinician-rated Anticholinergic Score (**CrAS**; Han et al., 2008)
- Anticholinergic Risk Scale (**ARS**; Rudolph et al., 2008)
- Anticholinergic Activity Scale (**AAS**; Ehrt et al., 2010)
- Anticholinergic Cognitive Burden scale (**ACB**; Boustani et al., 2008, updated in 2012)
- Anticholinergic Loading Scale (**ALS**; Sittironnarit et al., 2011)
- Delirogenic Risk Scale (**DRS**; Hefner et al., 2015)
- Anticholinergic Effect on Cognition **(AEC**; Bishara et al., 2016, updated regularly on the website https://medichec.com/; we used the online version available in November 2022)
- modified Anticholinergic Risk Scale (**mARS**; Sumukadas et al., 2016)
- Anticholinergic Impregnation Scale (**AIS**; Briet et al., 2017, updated in 2022 by the authors (Javelot et al., 2022))
- Muscarinic Acetylcholinergic Receptor ANTagonist Exposure Scale (**MARANTE**; Klamer et al., 2017, update of the scale received from the authors in October 2022)
- German Anticholinergic Burden Scale (**German ACB**; Kiesel et al., 2018)
- Korean Anticholinergic Burden Scale (**KABS**; Jun et al., 2019)
- Brazilian Anticholinergic Activity Drug Scale (**Brazilian ADS**; Nery et al., 2019)
- First modified Anticholinergic Cognitive Burden Scale (**mACB1** by Kable et al., 2019)
- Second modified Anticholinergic Cognitive Burden Scale (**mACB2** by Joshi et al., 2021)
- CRIDECO Anticholinergic Load Scale (**CALS**; Ramos et al., 2022)

Most discrete scales scored between 0 (drugs with no anticholinergic potency) and 3 (drugs with high anticholinergic potency). Four numerical scales were continuous:

- the Clinical and Pharmacological indices assess anticholinergic burden with benztropine equivalents (**CI** and **PI**; Minzenberg et al., 2004)
- the Drug Burden Index uses the WHO Defined Daily Doses to compute a score between 0 and 1 (**DBI-WHO**; Dauphinot et al., 2014)
- the Anticholinergic Toxicity Scale uses a score between 0 and 5 (**ATS**; Xu et al., 2017)

For the DBI-WHO, we used the list of anticholinergic drugs used by Dispenette et al. (2014) because they were the first to provide a list of drugs for the DBI-WHO.

In addition, four scales were categorical:

- **Peters**’s Scale (Peters, 1989): We coded “low” anticholinergic activity as 1, “intermediate” as 2, and “high” as 3.
- **Chew**’s Scale (Chew et al., 2008): We used the same scoring as the AAS scale, which is derived from Chew’s scale, and coded “0” as 0, “+/0” as 0.5, “+” as 1, “++” as 2, and “+++” as 3.
- **Durán**’s Scale (Durán et al., 2013): We used the same scoring as a recent comparative analysis of scales (Lisibach et al., 2021) and coded “0” as 0, “0 or 1” as 0.5, drugs listed in “Table 4” as 1, “weak” as 2, and “strong” as 3.

- **Salahudeen**'s Scale (Salahudeen et al., 2015) : We took the minimum score when several scores were proposed, for example, “1 or 2” was coded as 1 and “2 or 3” was coded as 2.

Among the 27 scales, five were dose-dependent (DBI-WHO, CI/PI, MARANTE, and Summer’s scale). The AIS scale provides information on the ability of the drugs to cross the brain-blood barrier/BBB (Javelot et al., 2022). However, only six of 2,031 patients took a drug that could not cross the BBB. Therefore, we did not account for the drug’s ability to cross the BBB in the scoring.

For the proportion of treated patients with a non-zero score in each scale: see **Supplementary Figure 2**.

##### 1.2. Cognitive impairment criteria

To compute the GDS criterion, we followed the guidelines of Heaton et al., 2004. T-scores were converted to deficit scores (ds) for each cognitive test, knowing that:

- ds = 0, if T score ≥ 40 (no impairment)
- ds = 1, if T score of 35–39 (mild impairment)
- ds = 2, if T score of 30–34 (mild-to-moderate impairment)
- ds = 3, if T score of 25–29 (moderate impairment)
- ds = 4, if T score of 20–24 (moderate-to-severe impairment)
- ds = 5, if T score < 20 (severe impairment)

We averaged ds over all cognitive tests. A participant was considered to be cognitively impaired if the average ds was ≥ 0.5, i.e., when half of the neuropsychological variables indicated mild impairment.

#### 2. Multiple logistic regression analysis of cognitive impairment

Statistical analyses were conducted using R version 4.2.2.

##### 2.1. Covariate selection

We ran pairwise correlation analyses for the 50 imputed datasets between the score in the Anticholinergic Cognitive Burden Scale (ACB; Boustani et al., 2008), the scale validated by the most studies (Lisibach et al., 2021), and each variable among age, sex, education level, the use of antidepressants, antipsychotics, anticonvulsants, lithium, anxiolytics, or antiparkinsonian drugs, MADRS score, YMRS score, the subtype of bipolar disorder, history of psychosis, the number of previous depressive, manic, hypomanic, or mixed episodes, age at first thymic episode, and CGI score (**Supplementary Table 1**).

Variables associated with ACB with a p-value < 0.2, i.e., MADRS score, history of psychosis, the use of antidepressants, lithium, antipsychotics, anxiolytics, or antiparkinsonian drugs, the number of previous depressive and manic episodes, age at first thymic episode, CGI score, and the education level, were used as covariates in the analysis.

#### 2.2. Results of multiple logistic regression analysis

For the multiple logistic regression analyses of scales with a non-significant association with cognitive impairment, see **Supplementary Table 2**.

For multiple logistic regression analyses of other correlates of iatrogenic cognitive burden (number of medications, number of antipsychotics, chlorpromazine equivalents, lorazepam equivalents), see **Supplementary Table 3**.

### 3. Multiple logistic regression analysis of peripheral and central anticholinergic side effect subscores

For the bivariable logistic regression analyses of peripheral and central anticholinergic side effect subscores with scales as the predictors, see **Supplementary Figure 3**.

For the multiple logistic regression analyses of peripheral anticholinergic side effect subscores with scales as the main predictors, see **Supplementary Table 4**.

For the multiple logistic regression analyses of central anticholinergic side effect subscores with scales as the main predictors, see **Supplementary Table 5**.

The 13 scales significantly associated with the central subscore in multiple regression analyses were: the Anticholinergic Cognitive Burden Scale, the Anticholinergic Risk Scale, the Anticholinergic Effect on Cognition, the Anticholinergic Impregnation Scale, the Brazilian Anticholinergic Activity Drug Scale, the CRIDECO Anticholinergic Load Scale, the Delirogenic Risk Scale, Durán's scale, the German Anticholinergic Burden Scale, the Korean Anticholinergic Burden Scale, the modified Anticholinergic Cognitive Burden Scale of Joshi et al. (2021), the modified Anticholinergic Risk Scale of Sumukadas et al. (2016), and Salahudeen's Scale.

**Supplementary Table 1. Pairwise correlations between the score in the Anticholinergic Cognitive Burden scale and the variables screened as potential covariates.**

| Variable | *Spearman coefficient* $\varrho$ | *p-value* |
|---|---|---|
| Sex | 0.002 | 0.946 |
| Age | -0.008 | 0.767 |
| Education level | -0.045 | 0.092 |
| MADRS | 0.050 | 0.064 |
| YMRS | -0.017 | 0.523 |
| CGI | 0.071 | 0.009 |
| Subtype of BD | -0.028 | 0.295 |
| Age at the first mood episode | 0.035 | 0.199 |
| Number of depressive episodes | 0.038 | 0.198 |
| Number of manic episodes | 0.025 | 0.151 |
| Number of hypomanic episodes | 0.039 | 0.397 |
| Number of mixed episodes | 0.027 | 0.350 |
| History of psychosis | 0.050 | 0.077 |
| Use of antidepressants | 0.176 | <.001 |
| Use of anticonvulsants | -0.034 | 0.200 |
| Use of lithium | -0.168 | <.001 |
| Use of antipsychotics | 0.611 | <.001 |

| | | |
|---|---|---|
| Use of anxiolytics | 0.178 | <.001 |
| Use of antiparkinsonian drugs | 0.131 | <.001 |

MADRS: Montgomery-Asberg Depression Rating Scale
YMRS: Young Mania Rating Scale
CGI: Clinical Global Impressions scale.

| | AAS (sum) | | | AAS (max) | | | ADS (sum) | | | ADS (max) | | | AEC (sum) | | | AEC (max) | | |
|---|---|---|---|---|---|---|---|---|---|---|---|---|---|---|---|---|---|---|
| | OR (95% CI) | p-value | fmi | OR (95% CI) | p-value | fmi | OR (95% CI) | p-value | fmi | OR (95% CI) | p-value | fmi | OR (95% CI) | p-value | fmi | OR (95% CI) | p-value | fmi |
| Scale | 1.03 (0.92-1.16) | 0.609 | 0.11 | 1.03 (0.9-1.18) | 0.644 | 0.13 | 1 (0.9-1.12) | 0.946 | 0.10 | 1 (0.86-1.15) | 0.956 | 0.14 | 1 (0.87-1.15) | 0.983 | 0.14 | 1.07 (0.89-1.3) | 0.479 | 0.13 |
| MADRS | 0.98 (0.94-1.03) | 0.420 | 0.18 | 0.98 (0.94-1.02) | 0.357 | 0.16 | 0.98 (0.94-1.03) | 0.417 | 0.18 | 0.98 (0.94-1.02) | 0.351 | 0.16 | 0.98 (0.94-1.03) | 0.416 | 0.18 | 0.98 (0.94-1.02) | 0.358 | 0.16 |
| History of psychosis | 0.9 (0.65-1.26) | 0.548 | 0.21 | 0.92 (0.65-1.31) | 0.642 | 0.30 | 0.9 (0.65-1.25) | 0.534 | 0.21 | 0.92 (0.65-1.3) | 0.624 | 0.29 | 0.9 (0.65-1.25) | 0.532 | 0.21 | 0.92 (0.65-1.31) | 0.643 | 0.29 |
| Antidepressants | 0.82 (0.59-1.14) | 0.229 | 0.18 | 0.84 (0.61-1.15) | 0.269 | 0.12 | 0.83 (0.6-1.15) | 0.270 | 0.18 | 0.85 (0.62-1.16) | 0.307 | 0.11 | 0.83 (0.6-1.16) | 0.275 | 0.17 | 0.83 (0.61-1.14) | 0.253 | 0.11 |
| Lithium | **1.95 (1.45-2.61)** | **0.000** | 0.16 | **1.94 (1.46-2.57)** | **0.000** | 0.10 | **1.94 (1.43-2.63)** | **0.000** | 0.16 | **1.94 (1.45-2.6)** | **0.000** | 0.11 | **1.94 (1.41-2.67)** | **0.000** | 0.15 | **1.88 (1.4-2.53)** | **0.000** | 0.11 |
| Antipsychotics | **1.61 (1.16-2.24)** | **0.005** | 0.15 | **1.61 (1.16-2.22)** | **0.004** | 0.12 | **1.66 (1.19-2.33)** | **0.003** | 0.14 | **1.68 (1.21-2.33)** | **0.002** | 0.12 | **1.67 (1.18-2.37)** | **0.004** | 0.14 | **1.57 (1.13-2.18)** | **0.007** | 0.12 |
| Anxiolytics | 1.25 (0.91-1.72) | 0.173 | 0.14 | 1.25 (0.9-1.72) | 0.178 | 0.13 | 1.25 (0.91-1.72) | 0.174 | 0.13 | 1.25 (0.9-1.72) | 0.178 | 0.13 | 1.25 (0.9-1.73) | 0.182 | 0.13 | 1.23 (0.89-1.7) | 0.218 | 0.13 |
| Antiparkisonian | **2.6 (1.02-6.61)** | **0.046** | 0.07 | **2.6 (1.03-6.59)** | **0.044** | 0.06 | **2.68 (1.06-6.77)** | **0.037** | 0.06 | **2.69 (1.07-6.74)** | **0.035** | 0.05 | **2.69 (1.06-6.81)** | **0.037** | 0.06 | **2.58 (1.02-6.5)** | **0.045** | 0.05 |
| Education level | **0.87 (0.82-0.92)** | **0.000** | 0.13 | **0.87 (0.82-0.92)** | **0.000** | 0.14 | **0.87 (0.82-0.92)** | **0.000** | 0.13 | **0.87 (0.82-0.92)** | **0.000** | 0.14 | **0.87 (0.82-0.92)** | **0.000** | 0.13 | **0.87 (0.82-0.92)** | **0.000** | 0.14 |
| Age at first episode | 1 (0.99-1.02) | 0.809 | 0.23 | 1 (0.99-1.02) | 0.811 | 0.23 | 1 (0.99-1.02) | 0.815 | 0.23 | 1 (0.99-1.02) | 0.809 | 0.23 | 1 (0.99-1.02) | 0.813 | 0.23 | 1 (0.99-1.02) | 0.844 | 0.23 |
| Number of manic episode | 1.07 (1-1.14) | 0.059 | 0.16 | 1.07 (1-1.15) | 0.059 | 0.24 | 1.07 (1-1.14) | 0.060 | 0.16 | 1.07 (1-1.15) | 0.060 | 0.24 | 1.07 (1-1.14) | 0.060 | 0.16 | 1.07 (1-1.15) | 0.059 | 0.24 |
| Number of depressive episode | 1 (0.98-1.03) | 0.889 | 0.35 | 1 (0.98-1.03) | 0.888 | 0.34 | 1 (0.98-1.03) | 0.905 | 0.35 | 1 (0.98-1.03) | 0.906 | 0.34 | 1 (0.98-1.03) | 0.907 | 0.34 | 1 (0.98-1.03) | 0.900 | 0.34 |
| CGI | 1.09 (0.99-1.21) | 0.076 | 0.15 | 1.09 (0.99-1.21) | 0.085 | 0.18 | 1.09 (0.99-1.21) | 0.078 | 0.15 | 1.09 (0.99-1.21) | 0.085 | 0.17 | 1.09 (0.99-1.21) | 0.078 | 0.15 | 1.09 (0.99-1.21) | 0.085 | 0.18 |

| | AIS (sum) | | | AIS (max) | | | ALS (sum) | | | ALS (max) | | | ARS (sum) | | | ARS (max) | | | Salahudeen (sum) | | |
|---|---|---|---|---|---|---|---|---|---|---|---|---|---|---|---|---|---|---|---|---|---|
| | OR (95% CI) | p-value | fmi | OR (95% CI) | p-value | fmi | OR (95% CI) | p-value | fmi | OR (95% CI) | p-value | fmi | OR (95% CI) | p-value | fmi | OR (95% CI) | p-value | fmi | OR (95% CI) | p-value | fmi |
| Scale | 1.01 (0.88-1.16) | 0.918 | 0.12 | 1.03 (0.82-1.29) | 0.806 | 0.10 | 1.22 (0.95-1.55) | 0.117 | 0.11 | 1.32 (0.98-1.79) | 0.069 | 0.10 | 1.11 (0.93-1.34) | 0.250 | 0.11 | 1.1 (0.9-1.35) | 0.355 | 0.11 | 1.08 (0.92-1.26) | 0.356 | 0.14 |
| MADRS | 0.98 (0.94-1.03) | 0.418 | 0.18 | 0.98 (0.94-1.02) | 0.356 | 0.16 | 0.98 (0.94-1.03) | 0.425 | 0.18 | 0.98 (0.94-1.03) | 0.383 | 0.16 | 0.98 (0.94-1.03) | 0.412 | 0.18 | 0.98 (0.94-1.02) | 0.350 | 0.16 | 0.98 (0.94-1.03) | 0.399 | 0.18 |
| History of psychosis | 0.9 (0.65-1.25) | 0.533 | 0.21 | 0.92 (0.65-1.3) | 0.631 | 0.29 | 0.9 (0.65-1.25) | 0.521 | 0.21 | 0.91 (0.64-1.29) | 0.611 | 0.29 | 0.91 (0.65-1.26) | 0.565 | 0.21 | 0.92 (0.65-1.31) | 0.657 | 0.30 | 0.9 (0.65-1.25) | 0.528 | 0.21 |
| Antidepressants | 0.83 (0.57-1.19) | 0.308 | 0.19 | 0.84 (0.62-1.15) | 0.289 | 0.12 | 0.7 (0.47-1.04) | 0.076 | 0.18 | 0.73 (0.51-1.04) | 0.084 | 0.14 | 0.82 (0.6-1.13) | 0.224 | 0.17 | 0.84 (0.62-1.15) | 0.276 | 0.12 | 0.77 (0.54-1.11) | 0.161 | 0.18 |
| Lithium | **1.94 (1.43-2.63)** | **0.000** | 0.15 | **1.93 (1.46-2.57)** | **0.000** | 0.10 | **1.6 (1.1-2.33)** | **0.015** | 0.12 | **1.62 (1.15-2.28)** | **0.006** | 0.10 | **1.95 (1.45-2.61)** | **0.000** | 0.16 | **1.94 (1.46-2.57)** | **0.000** | 0.10 | **1.86 (1.37-2.53)** | **0.000** | 0.15 |
| Antipsychotics | **1.65 (1.14-2.39)** | **0.008** | 0.12 | **1.63 (1.17-2.27)** | **0.004** | 0.12 | **1.63 (1.22-2.19)** | **0.001** | 0.15 | **1.67 (1.25-2.22)** | **0.001** | 0.12 | **1.52 (1.09-2.13)** | **0.014** | 0.15 | **1.54 (1.11-2.15)** | **0.011** | 0.13 | **1.56 (1.14-2.15)** | **0.006** | 0.13 |
| Anxiolytics | 1.24 (0.87-1.76) | 0.234 | 0.12 | 1.24 (0.89-1.72) | 0.199 | 0.14 | 1.15 (0.83-1.61) | 0.398 | 0.12 | 1.2 (0.87-1.65) | 0.278 | 0.13 | 1.22 (0.88-1.68) | 0.237 | 0.13 | 1.22 (0.88-1.69) | 0.229 | 0.13 | 1.17 (0.82-1.66) | 0.379 | 0.13 |
| Antiparkisonian | 2.63 (0.95-7.3) | 0.063 | 0.07 | 2.59 (0.99-6.76) | 0.052 | 0.06 | 2.47 (0.98-6.25) | 0.056 | 0.06 | 2.48 (0.98-6.25) | 0.054 | 0.06 | **2.75 (1.09-6.91)** | **0.032** | 0.06 | **2.73 (1.09-6.86)** | **0.033** | 0.06 | 2.1 (0.73-6.01) | 0.168 | 0.06 |
| Education level | **0.87 (0.82-0.92)** | **0.000** | 0.13 | **0.87 (0.82-0.92)** | **0.000** | 0.14 | **0.87 (0.82-0.92)** | **0.000** | 0.13 | **0.87 (0.82-0.92)** | **0.000** | 0.14 | **0.87 (0.82-0.92)** | **0.000** | 0.13 | **0.87 (0.82-0.92)** | **0.000** | 0.14 | **0.87 (0.82-0.92)** | **0.000** | 0.13 |
| Age at first episode | 1 (0.99-1.02) | 0.816 | 0.24 | 1 (0.99-1.02) | 0.821 | 0.23 | 1 (0.99-1.02) | 0.780 | 0.24 | 1 (0.99-1.02) | 0.823 | 0.23 | 1 (0.99-1.02) | 0.815 | 0.23 | 1 (0.99-1.02) | 0.809 | 0.23 | 1 (0.99-1.02) | 0.845 | 0.24 |
| Number of manic episode | 1.07 (1-1.14) | 0.060 | 0.16 | 1.07 (1-1.15) | 0.059 | 0.24 | 1.07 (1-1.14) | 0.056 | 0.16 | 1.07 (1-1.15) | 0.060 | 0.23 | 1.07 (1-1.14) | 0.058 | 0.16 | 1.07 (1-1.15) | 0.059 | 0.24 | 1.07 (0.99-1.14) | 0.070 | 0.16 |
| Number of depressive episode | 1 (0.98-1.03) | 0.908 | 0.35 | 1 (0.98-1.03) | 0.908 | 0.34 | 1 (0.98-1.03) | 0.921 | 0.34 | 1 (0.98-1.03) | 0.910 | 0.34 | 1 (0.98-1.03) | 0.875 | 0.34 | 1 (0.98-1.03) | 0.883 | 0.34 | 1 (0.98-1.03) | 0.895 | 0.34 |
| CGI | 1.09 (0.99-1.21) | 0.078 | 0.15 | 1.09 (0.99-1.21) | 0.085 | 0.18 | 1.09 (0.99-1.21) | 0.077 | 0.15 | 1.09 (0.99-1.21) | 0.085 | 0.17 | 1.09 (0.99-1.21) | 0.079 | 0.15 | 1.09 (0.99-1.21) | 0.086 | 0.18 | 1.09 (0.99-1.2) | 0.082 | 0.15 |

| | BADS (sum) | | | BADS (max) | | | CALS (sum) | | | CALS (max) | | | Chew (sum) | | | CI (sum) | | |
|---|---|---|---|---|---|---|---|---|---|---|---|---|---|---|---|---|---|---|
| | OR (95% CI) | p-value | fmi | OR (95% CI) | p-value | fmi | OR (95% CI) | p-value | fmi | OR (95% CI) | p-value | fmi | OR (95% CI) | p-value | fmi | OR (95% CI) | p-value | fmi |
| Scale | 1.04 (0.93-1.17) | 0.457 | 0.12 | 1.06 (0.91-1.25) | 0.446 | 0.09 | 1.04 (0.9-1.2) | 0.608 | 0.15 | 1.07 (0.85-1.34) | 0.590 | 0.13 | 1.13 (0.74-1.74) | 0.193 | 0.15 | 1.01 (0.81-1.25) | 0.937 | 0.14 |
| MADRS | 0.98 (0.94-1.03) | 0.415 | 0.18 | 0.98 (0.94-1.02) | 0.358 | 0.16 | 0.98 (0.94-1.03) | 0.412 | 0.18 | 0.98 (0.94-1.02) | 0.354 | 0.16 | 0.98 (0.94-1.03) | 0.414 | 0.14 | 0.98 (0.94-1.03) | 0.416 | 0.18 |
| History of psychosis | 0.91 (0.65-1.26) | 0.553 | 0.21 | 0.92 (0.65-1.31) | 0.647 | 0.29 | 0.9 (0.65-1.25) | 0.537 | 0.21 | 0.92 (0.65-1.3) | 0.635 | 0.29 | 0.93 (0.67-1.29) | 0.653 | 0.20 | 0.9 (0.65-1.25) | 0.534 | 0.21 |
| Antidepressants | 0.79 (0.56-1.12) | 0.193 | 0.19 | 0.83 (0.6-1.14) | 0.241 | 0.12 | 0.8 (0.56-1.14) | 0.222 | 0.19 | 0.84 (0.61-1.14) | 0.266 | 0.12 | 0.79 (0.55-1.12) | 0.157 | 0.11 | 0.83 (0.61-1.14) | 0.261 | 0.17 |
| Lithium | **1.89 (1.39-2.56)** | **0.000** | 0.15 | **1.92 (1.44-2.55)** | **0.000** | 0.11 | **1.9 (1.4-2.58)** | **0.000** | 0.15 | **1.93 (1.45-2.57)** | **0.000** | 0.10 | **1.72 (1.23-2.41)** | **0.002** | 0.17 | **1.95 (1.45-2.61)** | **0.000** | 0.16 |
| Antipsychotics | **1.58 (1.15-2.19)** | **0.005** | 0.14 | **1.6 (1.18-2.17)** | **0.003** | 0.12 | **1.58 (1.11-2.25)** | **0.011** | 0.12 | **1.6 (1.16-2.21)** | **0.004** | 0.12 | **1.5 (1.09-2.06)** | **0.013** | 0.12 | **1.66 (1.21-2.29)** | **0.002** | 0.14 |
| Anxiolytics | 1.22 (0.88-1.69) | 0.230 | 0.14 | 1.24 (0.9-1.71) | 0.197 | 0.14 | 1.21 (0.86-1.7) | 0.271 | 0.13 | 1.23 (0.89-1.71) | 0.204 | 0.14 | 1.24 (0.88-1.74) | 0.216 | 0.21 | 1.25 (0.91-1.72) | 0.173 | 0.14 |
| Antiparkisonian | **2.57 (1.01-6.52)** | **0.047** | 0.06 | **2.61 (1.04-6.55)** | **0.042** | 0.05 | 2.39 (0.86-6.63) | 0.094 | 0.07 | 2.46 (0.94-6.46) | 0.067 | 0.05 | **2.78 (1.1-7.03)** | **0.030** | 0.06 | **2.68 (1.06-6.78)** | **0.037** | 0.06 |
| Education level | **0.87 (0.82-0.92)** | **0.000** | 0.13 | **0.87 (0.82-0.92)** | **0.000** | 0.14 | **0.87 (0.82-0.92)** | **0.000** | 0.13 | **0.87 (0.82-0.92)** | **0.000** | 0.14 | **0.87 (0.82-0.92)** | **0.000** | 0.15 | **0.87 (0.82-0.92)** | **0.000** | 0.13 |
| Age at first episode | 1 (0.99-1.02) | 0.832 | 0.23 | 1 (0.99-1.02) | 0.833 | 0.23 | 1 (0.99-1.02) | 0.832 | 0.23 | 1 (0.99-1.02) | 0.832 | 0.23 | 1 (0.99-1.02) | 0.769 | 0.20 | 1 (0.99-1.02) | 0.813 | 0.23 |
| Number of manic episode | 1.07 (1-1.14) | 0.064 | 0.16 | 1.07 (1-1.15) | 0.061 | 0.24 | 1.07 (1-1.14) | 0.064 | 0.16 | 1.07 (1-1.15) | 0.060 | 0.24 | 1.07 (0.99-1.14) | 0.072 | 0.20 | 1.07 (1-1.14) | 0.060 | 0.16 |
| Number of depressive episode | 1 (0.98-1.03) | 0.886 | 0.34 | 1 (0.98-1.03) | 0.889 | 0.34 | 1 (0.98-1.03) | 0.897 | 0.34 | 1 (0.98-1.03) | 0.903 | 0.34 | 1 (0.98-1.03) | 0.956 | 0.36 | 1 (0.98-1.03) | 0.907 | 0.34 |
| CGI | 1.09 (0.99-1.2) | 0.083 | 0.15 | 1.09 (0.99-1.21) | 0.090 | 0.17 | 1.09 (0.99-1.21) | 0.080 | 0.15 | 1.09 (0.99-1.21) | 0.087 | 0.18 | 1.09 (0.99-1.21) | 0.078 | 0.17 | 1.09 (0.99-1.21) | 0.078 | 0.15 |

| | CI (max) | | | DBI (sum) | | | DBI (max) | | | DRS (sum) | | | DRS (max) | | | Duran (sum) | | | Duran (max) | | |
|---|---|---|---|---|---|---|---|---|---|---|---|---|---|---|---|---|---|---|---|---|---|
| | OR (95% CI) | p-value | fmi | OR (95% CI) | p-value | fmi | OR (95% CI) | p-value | fmi | OR (95% CI) | p-value | fmi | OR (95% CI) | p-value | fmi | OR (95% CI) | p-value | fmi | OR (95% CI) | p-value | fmi |
| Scale | 1 (0.8-1.25) | 0.993 | 0.15 | 1.16 (0.74-1.8) | 0.518 | 0.17 | 1.79 (0.72-4.42) | 0.209 | 0.14 | 1.05 (0.95-1.15) | 0.350 | 0.11 | 1.14 (0.98-1.33) | 0.093 | 0.11 | 1.06 (0.94-1.18) | 0.357 | 0.15 | 1.11 (0.92-1.34) | 0.269 | 0.11 |
| MADRS | 0.98 (0.94-1.02) | 0.352 | 0.16 | 0.98 (0.94-1.03) | 0.396 | 0.18 | 0.98 (0.93-1.02) | 0.327 | 0.16 | 0.98 (0.94-1.03) | 0.418 | 0.18 | 0.98 (0.94-1.02) | 0.358 | 0.16 | 0.98 (0.94-1.03) | 0.418 | 0.18 | 0.98 (0.94-1.02) | 0.355 | 0.16 |
| History of psychosis | 0.92 (0.65-1.3) | 0.627 | 0.30 | 0.9 (0.65-1.25) | 0.540 | 0.21 | 0.91 (0.64-1.29) | 0.603 | 0.30 | 0.91 (0.65-1.26) | 0.561 | 0.21 | 0.92 (0.65-1.31) | 0.655 | 0.29 | 0.91 (0.65-1.26) | 0.554 | 0.21 | 0.92 (0.65-1.3) | 0.641 | 0.29 |
| Antidepressants | 0.85 (0.62-1.16) | 0.301 | 0.12 | 0.79 (0.54-1.14) | 0.204 | 0.20 | 0.79 (0.57-1.1) | 0.167 | 0.12 | 0.78 (0.55-1.11) | 0.167 | 0.17 | 0.79 (0.58-1.09) | 0.157 | 0.12 | 0.8 (0.57-1.12) | 0.188 | 0.18 | 0.83 (0.61-1.13) | 0.236 | 0.12 |
| Lithium | **1.94 (1.46-2.57)** | **0.000** | 0.10 | **1.84 (1.32-2.57)** | **0.000** | 0.14 | **1.79 (1.32-2.44)** | **0.000** | 0.10 | **1.81 (1.3-2.52)** | **0.000** | 0.15 | **1.76 (1.3-2.4)** | **0.000** | 0.13 | **1.76 (1.23-2.52)** | **0.002** | 0.14 | **1.77 (1.27-2.47)** | **0.001** | 0.14 |
| Antipsychotics | **1.67 (1.22-2.29)** | **0.001** | 0.11 | **1.6 (1.16-2.2)** | **0.004** | 0.14 | **1.61 (1.21-2.16)** | **0.001** | 0.12 | **1.59 (1.16-2.17)** | **0.004** | 0.15 | **1.58 (1.18-2.12)** | **0.002** | 0.12 | **1.56 (1.12-2.16)** | **0.008** | 0.14 | **1.6 (1.19-2.15)** | **0.002** | 0.12 |
| Anxiolytics | 1.25 (0.9-1.72) | 0.178 | 0.13 | 1.21 (0.87-1.69) | 0.252 | 0.13 | 1.23 (0.89-1.7) | 0.207 | 0.14 | 1.17 (0.83-1.66) | 0.366 | 0.14 | 1.19 (0.86-1.65) | 0.293 | 0.14 | 1.17 (0.83-1.66) | 0.362 | 0.13 | 1.21 (0.87-1.67) | 0.256 | 0.14 |
| Antiparkisonian | **2.69 (1.07-6.76)** | **0.036** | 0.05 | **2.61 (1.03-6.58)** | **0.042** | 0.06 | **2.65 (1.06-6.62)** | **0.038** | 0.05 | 2.54 (1-6.45) | 0.050 | 0.06 | 2.4 (0.95-6.09) | 0.066 | 0.05 | 2.28 (0.85-6.11) | 0.101 | 0.07 | 2.31 (0.89-5.98) | 0.084 | 0.05 |
| Education level | **0.87 (0.82-0.92)** | **0.000** | 0.14 | **0.87 (0.82-0.92)** | **0.000** | 0.13 | **0.87 (0.82-0.92)** | **0.000** | 0.14 | **0.87 (0.82-0.92)** | **0.000** | 0.13 | **0.87 (0.82-0.92)** | **0.000** | 0.14 | **0.87 (0.82-0.92)** | **0.000** | 0.13 | **0.87 (0.82-0.92)** | **0.000** | 0.14 |
| Age at first episode | 1 (0.99-1.02) | 0.811 | 0.23 | 1 (0.99-1.02) | 0.826 | 0.23 | 1 (0.99-1.02) | 0.819 | 0.23 | 1 (0.99-1.02) | 0.834 | 0.23 | 1 (0.99-1.02) | 0.866 | 0.24 | 1 (0.99-1.02) | 0.830 | 0.23 | 1 (0.99-1.02) | 0.828 | 0.23 |
| Number of manic episode | 1.07 (1-1.15) | 0.060 | 0.24 | 1.07 (1-1.14) | 0.068 | 0.16 | 1.07 (1-1.15) | 0.067 | 0.24 | 1.07 (1-1.14) | 0.064 | 0.16 | 1.07 (1-1.15) | 0.065 | 0.23 | 1.07 (1-1.14) | 0.062 | 0.16 | 1.07 (1-1.15) | 0.057 | 0.24 |
| Number of depressive episode | 1 (0.98-1.03) | 0.905 | 0.34 | 1 (0.98-1.03) | 0.918 | 0.34 | 1 (0.98-1.03) | 0.924 | 0.34 | 1 (0.98-1.03) | 0.872 | 0.34 | 1 (0.98-1.03) | 0.872 | 0.34 | 1 (0.98-1.03) | 0.887 | 0.34 | 1 (0.98-1.03) | 0.885 | 0.34 |
| CGI | 1.09 (0.99-1.21) | 0.085 | 0.18 | 1.09 (0.99-1.21) | 0.080 | 0.15 | 1.09 (0.99-1.21) | 0.085 | 0.18 | 1.09 (0.99-1.21) | 0.079 | 0.15 | 1.09 (0.99-1.21) | 0.090 | 0.18 | 1.09 (0.99-1.21) | 0.079 | 0.15 | 1.09 (0.99-1.21) | 0.086 | 0.18 |

| | GACB (sum) | | | GACB (max) | | | KABS (sum) | | | KABS (max) | | | MARANTE (sum) | | | MARANTE (max) | | |
|---|---|---|---|---|---|---|---|---|---|---|---|---|---|---|---|---|---|---|
| | OR (95% CI) | p-value | fmi | OR (95% CI) | p-value | fmi | OR (95% CI) | p-value | fmi | OR (95% CI) | p-value | fmi | OR (95% CI) | p-value | fmi | OR (95% CI) | p-value | fmi |
| Scale | 1.05 (0.92-1.2) | 0.494 | 0.13 | 1.06 (0.85-1.32) | 0.577 | 0.12 | 1.09 (0.95-1.25) | 0.233 | 0.14 | 1.04 (0.87-1.24) | 0.696 | 0.10 | 1.12 (0.98-1.28) | 0.086 | 0.15 | 1.19 (1-1.42) | 0.055 | 0.12 |
| MADRS | 0.98 (0.94-1.03) | 0.407 | 0.18 | 0.98 (0.94-1.02) | 0.351 | 0.16 | 0.98 (0.94-1.03) | 0.405 | 0.18 | 0.98 (0.94-1.02) | 0.354 | 0.16 | 0.98 (0.94-1.03) | 0.408 | 0.18 | 0.98 (0.94-1.02) | 0.349 | 0.16 |
| History of psychosis | 0.9 (0.65-1.25) | 0.535 | 0.21 | 0.92 (0.65-1.3) | 0.633 | 0.29 | 0.9 (0.65-1.26) | 0.549 | 0.21 | 0.92 (0.65-1.3) | 0.635 | 0.29 | 0.91 (0.66-1.27) | 0.582 | 0.21 | 0.92 (0.65-1.31) | 0.657 | 0.29 |
| Antidepressants | 0.79 (0.56-1.13) | 0.197 | 0.19 | 0.84 (0.61-1.14) | 0.266 | 0.12 | 0.76 (0.54-1.09) | 0.136 | 0.19 | 0.83 (0.6-1.15) | 0.268 | 0.11 | 0.76 (0.54-1.06) | 0.107 | 0.18 | 0.78 (0.57-1.08) | 0.138 | 0.12 |
| Lithium | **1.89 (1.4-2.56)** | **0.000** | 0.15 | **1.93 (1.45-2.57)** | **0.000** | 0.10 | **1.96 (1.46-2.63)** | **0.000** | 0.16 | **1.94 (1.46-2.58)** | **0.000** | 0.10 | **1.74 (1.26-2.4)** | **0.001** | 0.16 | **1.79 (1.33-2.41)** | **0.000** | 0.12 |
| Antipsychotics | **1.56 (1.11-2.21)** | **0.011** | 0.12 | **1.6 (1.17-2.21)** | **0.004** | 0.14 | 1.44 (0.99-2.1) | 0.059 | 0.14 | **1.59 (1.1-2.3)** | **0.013** | 0.11 | **1.47 (1.06-2.04)** | **0.021** | 0.16 | **1.5 (1.1-2.04)** | **0.010** | 0.12 |
| Anxiolytics | 1.21 (0.87-1.69) | 0.259 | 0.13 | 1.24 (0.89-1.71) | 0.199 | 0.14 | 1.18 (0.84-1.64) | 0.344 | 0.13 | 1.23 (0.89-1.71) | 0.205 | 0.13 | 1.17 (0.84-1.62) | 0.360 | 0.13 | 1.19 (0.86-1.65) | 0.294 | 0.13 |
| Antiparkisonian | 2.52 (0.98-6.45) | 0.054 | 0.06 | **2.6 (1.03-6.55)** | **0.043** | 0.05 | 2.48 (0.98-6.3) | 0.056 | 0.06 | **2.65 (1.05-6.64)** | **0.038** | 0.05 | 2.4 (0.94-6.12) | 0.067 | 0.06 | 2.37 (0.94-6) | 0.069 | 0.05 |
| Education level | **0.87 (0.82-0.92)** | **0.000** | 0.13 | **0.87 (0.82-0.92)** | **0.000** | 0.14 | **0.87 (0.82-0.92)** | **0.000** | 0.13 | **0.87 (0.82-0.92)** | **0.000** | 0.14 | **0.87 (0.82-0.92)** | **0.000** | 0.13 | **0.87 (0.82-0.92)** | **0.000** | 0.14 |
| Age at first episode | 1 (0.99-1.02) | 0.834 | 0.23 | 1 (0.99-1.02) | 0.836 | 0.23 | 1 (0.99-1.02) | 0.845 | 0.23 | 1 (0.99-1.02) | 0.819 | 0.23 | 1 (0.99-1.02) | 0.837 | 0.23 | 1 (0.99-1.02) | 0.837 | 0.23 |
| Number of manic episode | 1.07 (1-1.14) | 0.065 | 0.16 | 1.07 (1-1.15) | 0.060 | 0.24 | 1.07 (1-1.14) | 0.061 | 0.16 | 1.07 (1-1.15) | 0.060 | 0.24 | 1.07 (1-1.14) | 0.064 | 0.16 | 1.07 (1-1.15) | 0.062 | 0.23 |
| Number of depressive episode | 1 (0.98-1.03) | 0.895 | 0.35 | 1 (0.98-1.03) | 0.902 | 0.34 | 1 (0.98-1.03) | 0.882 | 0.35 | 1 (0.98-1.03) | 0.896 | 0.34 | 1 (0.98-1.03) | 0.862 | 0.34 | 1 (0.98-1.03) | 0.882 | 0.34 |
| CGI | 1.09 (0.99-1.21) | 0.080 | 0.15 | 1.09 (0.99-1.21) | 0.086 | 0.18 | 1.09 (0.99-1.2) | 0.082 | 0.15 | 1.09 (0.99-1.21) | 0.087 | 0.17 | 1.09 (0.99-1.21) | 0.076 | 0.15 | 1.09 (0.99-1.21) | 0.079 | 0.18 |

| | mARS (sum) | | | mARS (max) | | | Peters (sum) | | | Peters (max) | | | PI (sum) | | | PI (max) | | | Salahudeen (max) | | |
|---|---|---|---|---|---|---|---|---|---|---|---|---|---|---|---|---|---|---|---|---|---|
| | OR (95% CI) | p-value | fmi | OR (95% CI) | p-value | fmi | OR (95% CI) | p-value | fmi | OR (95% CI) | p-value | fmi | OR (95% CI) | p-value | fmi | OR (95% CI) | p-value | fmi | OR (95% CI) | p-value | fmi |
| Scale | 1.1 (0.93-1.32) | 0.265 | 0.12 | 1.11 (0.91-1.35) | 0.305 | 0.10 | 1.19 (0.76-1.87) | 0.438 | 0.08 | 1.18 (0.73-1.9) | 0.500 | 0.10 | 1.03 (0.98-1.08) | 0.303 | 0.25 | 1.03 (0.98-1.08) | 0.305 | 0.20 | 1.17 (0.89-1.52) | 0.255 | 0.12 |
| MADRS | 0.98 (0.94-1.03) | 0.420 | 0.18 | 0.98 (0.94-1.02) | 0.360 | 0.16 | 0.98 (0.94-1.03) | 0.391 | 0.18 | 0.98 (0.93-1.02) | 0.331 | 0.16 | 0.98 (0.94-1.03) | 0.422 | 0.18 | 0.98 (0.94-1.02) | 0.357 | 0.16 | 0.98 (0.94-1.02) | 0.338 | 0.16 |
| History of psychosis | 0.91 (0.65-1.26) | 0.564 | 0.21 | 0.92 (0.65-1.31) | 0.659 | 0.30 | 0.9 (0.65-1.25) | 0.528 | 0.21 | 0.92 (0.65-1.3) | 0.622 | 0.29 | 0.9 (0.65-1.26) | 0.551 | 0.21 | 0.92 (0.65-1.31) | 0.646 | 0.29 | 0.91 (0.64-1.3) | 0.614 | 0.29 |
| Antidepressants | 0.81 (0.59-1.12) | 0.204 | 0.18 | 0.83 (0.61-1.14) | 0.250 | 0.12 | 0.83 (0.6-1.14) | 0.243 | 0.17 | 0.84 (0.62-1.15) | 0.284 | 0.12 | 0.83 (0.61-1.15) | 0.263 | 0.17 | 0.85 (0.63-1.16) | 0.305 | 0.12 | 0.82 (0.6-1.13) | 0.224 | 0.12 |
| Lithium | **1.95 (1.45-2.62)** | **0.000** | 0.16 | **1.94 (1.46-2.58)** | **0.000** | 0.10 | **1.95 (1.45-2.61)** | **0.000** | 0.16 | **1.94 (1.46-2.57)** | **0.000** | 0.10 | **1.95 (1.45-2.62)** | **0.000** | 0.16 | **1.94 (1.46-2.58)** | **0.000** | 0.10 | **1.92 (1.44-2.55)** | **0.000** | 0.10 |
| Antipsychotics | **1.53 (1.1-2.13)** | **0.012** | 0.16 | **1.54 (1.11-2.13)** | **0.011** | 0.12 | **1.66 (1.24-2.23)** | **0.001** | 0.16 | **1.66 (1.25-2.21)** | **0.001** | 0.12 | **1.64 (1.22-2.2)** | **0.001** | 0.15 | **1.64 (1.23-2.18)** | **0.001** | 0.12 | **1.63 (1.22-2.18)** | **0.001** | 0.13 |
| Anxiolytics | 1.21 (0.88-1.68) | 0.243 | 0.14 | 1.21 (0.88-1.68) | 0.241 | 0.13 | 1.25 (0.91-1.72) | 0.170 | 0.14 | 1.25 (0.91-1.72) | 0.175 | 0.14 | 1.25 (0.9-1.72) | 0.180 | 0.14 | 1.24 (0.9-1.71) | 0.184 | 0.13 | 1.2 (0.86-1.67) | 0.275 | 0.14 |
| Antiparkisonian | **2.76 (1.1-6.94)** | **0.031** | 0.06 | **2.74 (1.09-6.9)** | **0.032** | 0.06 | 2.17 (0.73-6.45) | 0.164 | 0.09 | 2.23 (0.76-6.57) | 0.146 | 0.09 | **2.59 (1.03-6.54)** | **0.044** | 0.06 | **2.59 (1.03-6.52)** | **0.044** | 0.06 | 2.02 (0.72-5.68) | 0.183 | 0.06 |
| Education level | **0.87 (0.82-0.92)** | **0.000** | 0.13 | **0.87 (0.82-0.92)** | **0.000** | 0.14 | **0.87 (0.82-0.92)** | **0.000** | 0.13 | **0.87 (0.82-0.92)** | **0.000** | 0.14 | **0.87 (0.82-0.92)** | **0.000** | 0.13 | **0.87 (0.82-0.92)** | **0.000** | 0.14 | **0.87 (0.82-0.92)** | **0.000** | 0.14 |
| Age at first episode | 1 (0.99-1.02) | 0.840 | 0.23 | 1 (0.99-1.02) | 0.830 | 0.23 | 1 (0.99-1.02) | 0.827 | 0.23 | 1 (0.99-1.02) | 0.823 | 0.23 | 1 (0.99-1.02) | 0.789 | 0.24 | 1 (0.99-1.02) | 0.788 | 0.23 | 1 (0.99-1.02) | 0.869 | 0.23 |
| Number of manic episode | 1.07 (1-1.14) | 0.061 | 0.16 | 1.07 (1-1.15) | 0.060 | 0.24 | 1.07 (1-1.14) | 0.059 | 0.16 | 1.07 (1-1.15) | 0.059 | 0.24 | 1.06 (0.99-1.14) | 0.083 | 0.16 | 1.07 (0.99-1.15) | 0.080 | 0.24 | 1.07 (1-1.15) | 0.063 | 0.23 |
| Number of depressive episode | 1 (0.98-1.03) | 0.882 | 0.34 | 1 (0.98-1.03) | 0.890 | 0.34 | 1 (0.98-1.03) | 0.897 | 0.34 | 1 (0.98-1.03) | 0.895 | 0.34 | 1 (0.98-1.03) | 0.888 | 0.34 | 1 (0.98-1.03) | 0.887 | 0.34 | 1 (0.98-1.03) | 0.910 | 0.34 |
| CGI | 1.09 (0.99-1.21) | 0.080 | 0.15 | 1.09 (0.99-1.21) | 0.087 | 0.18 | 1.09 (0.99-1.21) | 0.076 | 0.15 | 1.09 (0.99-1.21) | 0.084 | 0.18 | 1.1 (0.99-1.21) | 0.071 | 0.16 | 1.1 (0.99-1.21) | 0.077 | 0.18 | 1.09 (0.99-1.21) | 0.087 | 0.18 |

Supplementary Table 2. Multiple logistic regression models with a non-significant association between the scale and cognitive impairment.

Significant associations are in bold.

AAS: Anticholinergic Activity Scale (Ehrt et al., 2010); ADS: Anticholinergic Drug Scale (Carnahan et al., 2006); AEC: Anticholinergic Effect on Cognition (Bishara et al., 2016); AIS: Anticholinergic Impregnation Scale (Briet et al., 2017); ALS: Anticholinergic Loading Scale (Sittironnarit et al., 2011); ARS: Anticholinergic Risk Scale (Rudolph et al., 2008); BADS: Brazilian Anticholinergic Drug Scale (Nery et al., 2019); CALS: CRIDECO Anticholinergic Load Scale (Ramos et al., 2022); Chew: Chew's scale (Chew et al., 2008); CI: Clinical index (Minzenberg et al., 2004); DBI: Drug burden index, WHO version (Dauphinot et al., 2014); DRS: Drug Delirogenic Scale (Hefner et al., 2015); Duran: Durán Scale (Durán et al., 2013); fmi: fraction of missing information; GACB: German Anticholinergic Burden Scale (Kiesel et al., 2018); KABS: Korean Anticholinergic Burden Scale (Jun et al., 2019); MARANTE: Muscarinic Acetylcholinergic Receptor ANTagonist Exposure Scale (Klamer et al., 2017); mARS: modified Anticholinergic Risk Scale (Sumukadas et al., 2016); Peters: Peters's scale (Peters, 1989); PI: Pharmacological Index (Minzenberg et al., 2004); Salahudeen: Salahudeen's scale (Salahudeen et al., 2015).

Supplementary Table 3. Multiple logistic regression models of cognitive impairment with alternative measures of iatrogenic cognitive burden as main predictors.

| | OR (95% CI) | p-value | fmi |
|---|---|---|---|
| CPZeq | 1 (1-1) | 0.818 | 0.16 |
| MADRS | 0.99 (0.95-1.04) | 0.728 | 0.18 |
| History of psychosis | 0.93 (0.68-1.29) | 0.672 | 0.23 |
| Antidepressants | 0.82 (0.61-1.12) | 0.212 | 0.15 |
| Lithium | **1.8 (1.36-2.39)** | **0.000** | 0.15 |
| Antipsychotics | **1.57 (1.1-2.23)** | **0.012** | 0.12 |
| Anxiolytics | 1.18 (0.85-1.62) | 0.325 | 0.14 |
| Antiparkisonian | 2.45 (0.96-6.27) | 0.061 | 0.06 |
| Education level | **0.87 (0.82-0.91)** | **0.000** | 0.14 |
| Age at first episode | 1.01 (0.99-1.02) | 0.398 | 0.23 |
| Number of manic episode | **1.08 (1.01-1.15)** | **0.032** | 0.16 |
| Number of depressive episode | 1.01 (0.98-1.03) | 0.661 | 0.33 |
| CGI | 1.08 (0.98-1.19) | 0.102 | 0.15 |

| | OR (95% CI) | p-value | fmi |
|---|---|---|---|
| Lorazepam equivalents | 1.09 (0.91-1.3) | 0.341 | 0.11 |
| MADRS | 0.98 (0.94-1.03) | 0.425 | 0.18 |
| History of psychosis | 0.9 (0.65-1.26) | 0.547 | 0.21 |
| Antidepressants | 0.84 (0.61-1.15) | 0.273 | 0.17 |
| Lithium | **1.95 (1.45-2.62)** | **0.000** | 0.16 |
| Antipsychotics | **1.66 (1.24-2.23)** | **0.001** | 0.15 |
| Anxiolytics | 1.15 (0.81-1.65) | 0.428 | 0.10 |
| Antiparkisonian | **2.58 (1.02-6.5)** | **0.045** | 0.06 |
| Education level | **0.87 (0.82-0.92)** | **0.000** | 0.13 |
| Age at first episode | 1 (0.99-1.02) | 0.829 | 0.23 |
| Number of manic episode | 1.07 (1-1.14) | 0.057 | 0.16 |
| Number of depressive episode | 1 (0.98-1.03) | 0.931 | 0.34 |
| CGI | 1.09 (0.99-1.21) | 0.078 | 0.15 |

| | OR (95% CI) | p-value | fmi |
|---|---|---|---|
| Number of antipsychotics | 1.71 (1-2.92) | 0.052 | 0.13 |
| MADRS | 0.98 (0.94-1.03) | 0.420 | 0.18 |
| History of psychosis | 0.89 (0.64-1.24) | 0.483 | 0.21 |
| Antidepressants | 0.82 (0.6-1.13) | 0.226 | 0.18 |
| Lithium | **1.98 (1.47-2.66)** | **0.000** | 0.16 |
| Antipsychotics | 0.93 (0.48-1.8) | 0.824 | 0.13 |
| Anxiolytics | 1.22 (0.88-1.68) | 0.230 | 0.13 |
| Antiparkisonian | **2.6 (1.03-6.53)** | **0.042** | 0.06 |
| Education level | **0.87 (0.82-0.92)** | **0.000** | 0.13 |
| Age at first episode | 1 (0.99-1.02) | 0.782 | 0.23 |
| Number of manic episode | 1.07 (1-1.14) | 0.062 | 0.16 |
| Number of depressive episode | 1 (0.98-1.03) | 0.862 | 0.35 |
| CGI | 1.09 (0.99-1.2) | 0.085 | 0.15 |

| | OR (95% CI) | p-value | fmi |
|---|---|---|---|
| Number of treatments | 1.03 (0.84-1.28) | 0.758 | 0.19 |
| MADRS | 0.98 (0.94-1.03) | 0.413 | 0.18 |
| History of psychosis | 0.9 (0.65-1.25) | 0.531 | 0.21 |
| Antidepressants | 0.81 (0.55-1.18) | 0.273 | 0.21 |
| Lithium | **1.91 (1.39-2.62)** | **0.000** | 0.15 |
| Antipsychotics | **1.62 (1.15-2.29)** | **0.006** | 0.15 |
| Anxiolytics | 1.19 (0.77-1.84) | 0.421 | 0.17 |
| Antiparkisonian | 2.56 (0.98-6.7) | 0.056 | 0.05 |
| Education level | **0.87 (0.82-0.92)** | **0.000** | 0.14 |
| Age at first episode | 1 (0.99-1.02) | 0.827 | 0.24 |
| Number of manic episode | 1.07 (1-1.14) | 0.060 | 0.16 |
| Number of depressive episode | 1 (0.98-1.03) | 0.915 | 0.35 |
| CGI | 1.09 (0.99-1.21) | 0.080 | 0.15 |

| Var | ABC (sum)[1] | | | ABC (max)[2] | | |
| --- | --- | --- | --- | --- | --- | --- |
| | Beta (95% CI) | p-value | fmi | Beta (95% CI) | p-value | fmi |
| Scale | **0.21 (0.08-0.35)** | **0.001** | 0.04 | **0.22 (0.06-0.37)** | **0.006** | 0.04 |
| MADRS | **0.16 (0.12-0.2)** | **<.001** | 0.07 | **0.16 (0.12-0.2)** | **<.001** | 0.07 |
| History of psychosis | -0.13 (-0.43-0.17) | 0.383 | 0.21 | -0.13 (-0.42-0.16) | 0.377 | 0.15 |
| Antidepressants | **0.28 (0.01-0.54)** | **0.043** | 0.06 | **0.28 (0.02-0.55)** | **0.037** | 0.04 |
| Lithium | 0.21 (-0.04-0.47) | 0.106 | 0.05 | 0.2 (-0.05-0.46) | 0.124 | 0.04 |
| Antipsychotics | 0.12 (-0.14-0.37) | 0.37 | 0.07 | 0.11 (-0.14-0.36) | 0.383 | 0.05 |
| Anxiolytics | 0.22 (-0.12-0.55) | 0.201 | 0.07 | 0.25 (-0.08-0.57) | 0.143 | 0.04 |
| Antiparkisonian | -0.11 (-1.13-0.91) | 0.832 | 0.01 | -0.08 (-1.11-0.95) | 0.882 | 0.01 |
| Education level | **-0.05 (-0.1-0)** | **0.035** | 0.05 | **-0.05 (-0.1-0)** | **0.037** | 0.05 |
| Age at first episode | 0.01 (-0.01-0.02) | 0.399 | 0.11 | 0.01 (-0.01-0.02) | 0.466 | 0.11 |
| Number of manic episode | -0.06 (-0.12-0.01) | 0.085 | 0.12 | -0.06 (-0.12-0.01) | 0.082 | 0.11 |
| Number of depressive episode | 0.01 (-0.01-0.03) | 0.344 | 0.27 | 0.01 (-0.01-0.03) | 0.31 | 0.24 |
| CGI | 0.03 (-0.06-0.12) | 0.496 | 0.09 | 0.03 (-0.06-0.12) | 0.537 | 0.08 |

[1] R2 = 0.096; adj R2 = 0.087; AIC = 6332
[2] R2 = 0.093; adj R2 = 0.085; AIC = 6331

| Var | ACB (sum)[1] | | | ACB (max)[2] | | |
| --- | --- | --- | --- | --- | --- | --- |
| | Beta (95% CI) | p-value | fmi | Beta (95% CI) | p-value | fmi |
| Scale | **0.16 (0.06-0.26)** | **0.002** | 0.05 | **0.2 (0.08-0.32)** | **0.001** | 0.05 |
| MADRS | **0.16 (0.12-0.2)** | **<.001** | 0.07 | **0.16 (0.12-0.2)** | **<.001** | 0.07 |
| History of psychosis | -0.13 (-0.43-0.17) | 0.396 | 0.21 | -0.12 (-0.41-0.17) | 0.416 | 0.15 |
| Antidepressants | 0.2 (-0.07-0.48) | 0.149 | 0.05 | 0.24 (-0.03-0.5) | 0.085 | 0.04 |
| Lithium | 0.22 (-0.04-0.47) | 0.097 | 0.05 | 0.21 (-0.04-0.47) | 0.1 | 0.04 |
| Antipsychotics | -0.2 (-0.53-0.13) | 0.225 | 0.07 | -0.21 (-0.53-0.11) | 0.199 | 0.06 |
| Anxiolytics | **0.39 (0.1-0.69)** | **0.009** | 0.07 | **0.43 (0.14-0.71)** | **0.004** | 0.05 |
| Antiparkisonian | 0.35 (-0.61-1.31) | 0.474 | 0.01 | 0.43 (-0.52-1.38) | 0.372 | 0.01 |
| Education level | **-0.05 (-0.1-0)** | **0.031** | 0.05 | **-0.05 (-0.1-0)** | **0.035** | 0.05 |
| Age at first episode | 0.01 (-0.01-0.02) | 0.404 | 0.11 | 0.01 (-0.01-0.02) | 0.474 | 0.11 |
| Number of manic episode | -0.05 (-0.12-0.01) | 0.103 | 0.11 | -0.05 (-0.12-0.01) | 0.092 | 0.10 |
| Number of depressive episode | 0.01 (-0.01-0.03) | 0.281 | 0.27 | 0.01 (-0.01-0.03) | 0.274 | 0.24 |
| CGI | 0.03 (-0.06-0.12) | 0.521 | 0.09 | 0.02 (-0.06-0.11) | 0.587 | 0.08 |

[1] R2 = 0.095; adj R2 = 0.087; AIC = 6332
[2] R2 = 0.095; adj R2 = 0.086; AIC = 6328

| Var | ADS (sum)[1] | | |
| --- | --- | --- | --- |
| | Beta (95% CI) | p-value | fmi |
| Scale | 0.06 (-0.04-0.15) | 0.251 | 0.04 |
| MADRS | **0.16 (0.12-0.2)** | **<.001** | 0.07 |
| History of psychosis | -0.13 (-0.43-0.17) | 0.385 | 0.21 |
| Antidepressants | 0.27 (0-0.55) | 0.052 | 0.06 |
| Lithium | 0.16 (-0.11-0.42) | 0.247 | 0.05 |
| Antipsychotics | 0.03 (-0.26-0.33) | 0.829 | 0.06 |
| Anxiolytics | **0.47 (0.18-0.76)** | **0.002** | 0.06 |
| Antiparkisonian | 0.41 (-0.55-1.37) | 0.399 | 0.01 |
| Education level | **-0.05 (-0.1-0)** | **0.03** | 0.05 |
| Age at first episode | 0.01 (-0.01-0.02) | 0.372 | 0.11 |
| Number of manic episode | -0.05 (-0.12-0.01) | 0.092 | 0.11 |
| Number of depressive episode | 0.01 (-0.01-0.03) | 0.258 | 0.27 |
| CGI | 0.03 (-0.06-0.12) | 0.525 | 0.09 |

[1] R2 = 0.09; adj R2 = 0.081; AIC = 6341

| Var | AEC (sum)[1] | | | AEC (max)[2] | | |
| --- | --- | --- | --- | --- | --- | --- |
| | Beta (95% CI) | p-value | fmi | Beta (95% CI) | p-value | fmi |
| Scale | 0.09 (-0.03-0.22) | 0.156 | 0.05 | 0.15 (-0.01-0.31) | 0.071 | 0.04 |
| MADRS | **0.16 (0.12-0.2)** | **<.001** | 0.07 | **0.16 (0.12-0.2)** | **<.001** | 0.07 |
| History of psychosis | -0.14 (-0.43-0.16) | 0.375 | 0.21 | -0.13 (-0.42-0.16) | 0.382 | 0.15 |
| Antidepressants | 0.26 (-0.02-0.54) | 0.067 | 0.05 | **0.27 (0-0.54)** | **0.049** | 0.04 |
| Lithium | 0.12 (-0.16-0.4) | 0.414 | 0.05 | 0.12 (-0.15-0.39) | 0.378 | 0.05 |
| Antipsychotics | -0.01 (-0.32-0.3) | 0.971 | 0.07 | -0.03 (-0.32-0.27) | 0.859 | 0.05 |
| Anxiolytics | **0.43 (0.14-0.73)** | **0.004** | 0.07 | **0.43 (0.14-0.72)** | **0.004** | 0.05 |
| Antiparkisonian | 0.39 (-0.57-1.35) | 0.425 | 0.01 | 0.38 (-0.58-1.34) | 0.437 | 0.01 |
| Education level | **-0.05 (-0.1--0.01)** | **0.03** | 0.05 | **-0.05 (-0.1-0)** | **0.031** | 0.05 |
| Age at first episode | 0.01 (-0.01-0.02) | 0.372 | 0.11 | 0.01 (-0.01-0.02) | 0.461 | 0.11 |
| Number of manic episode | -0.05 (-0.12-0.01) | 0.09 | 0.11 | -0.06 (-0.12-0.01) | 0.084 | 0.11 |
| Number of depressive episode | 0.01 (-0.01-0.03) | 0.264 | 0.27 | 0.01 (-0.01-0.03) | 0.265 | 0.24 |
| CGI | 0.03 (-0.06-0.12) | 0.515 | 0.09 | 0.03 (-0.06-0.11) | 0.561 | 0.08 |

[1] R2 = 0.09; adj R2 = 0.082; AIC = 6340
[2] R2 = 0.09; adj R2 = 0.082; AIC = 6335

| Var | AIS (sum)[1] | | | AIS (max)[2] | | |
| --- | --- | --- | --- | --- | --- | --- |
| | Beta (95% CI) | p-value | fmi | Beta (95% CI) | p-value | fmi |
| Scale | **0.24 (0.11-0.36)** | **<.001** | 0.04 | **0.27 (0.07-0.47)** | **0.008** | 0.04 |
| MADRS | **0.16 (0.12-0.2)** | **<.001** | 0.07 | **0.16 (0.12-0.2)** | **<.001** | 0.07 |
| History of psychosis | -0.12 (-0.42-0.17) | 0.415 | 0.20 | -0.13 (-0.42-0.16) | 0.374 | 0.15 |
| Antidepressants | 0.03 (-0.27-0.33) | 0.851 | 0.04 | 0.25 (-0.02-0.52) | 0.067 | 0.04 |
| Lithium | 0.06 (-0.2-0.33) | 0.63 | 0.04 | 0.18 (-0.08-0.43) | 0.168 | 0.04 |
| Antipsychotics | -0.28 (-0.6-0.05) | 0.097 | 0.05 | -0.09 (-0.38-0.21) | 0.561 | 0.04 |
| Anxiolytics | 0.21 (-0.1-0.53) | 0.186 | 0.05 | **0.41 (0.12-0.7)** | **0.006** | 0.05 |
| Antiparkisonian | -0.29 (-1.33-0.74) | 0.577 | 0.02 | 0.13 (-0.85-1.12) | 0.793 | 0.01 |
| Education level | **-0.05 (-0.1-0)** | **0.034** | 0.05 | **-0.05 (-0.1-0)** | **0.037** | 0.05 |
| Age at first episode | 0.01 (-0.01-0.02) | 0.449 | 0.11 | 0 (-0.01-0.02) | 0.487 | 0.11 |
| Number of manic episode | -0.06 (-0.12-0) | 0.071 | 0.12 | -0.06 (-0.12-0.01) | 0.081 | 0.10 |
| Number of depressive episode | 0.01 (-0.01-0.03) | 0.293 | 0.27 | 0.01 (-0.01-0.03) | 0.288 | 0.23 |
| CGI | 0.03 (-0.06-0.11) | 0.568 | 0.08 | 0.03 (-0.06-0.11) | 0.571 | 0.08 |

[1] R2 = 0.098; adj R2 = 0.09; AIC = 6327
[2] R2 = 0.093; adj R2 = 0.084; AIC = 6331

| Var | ALS (sum)[1] | | | ALS (max)[2] | | |
| --- | --- | --- | --- | --- | --- | --- |
| | Beta (95% CI) | p-value | fmi | Beta (95% CI) | p-value | fmi |
| Scale | 0.13 (-0.1-0.35) | 0.268 | 0.07 | 0.04 (-0.22-0.31) | 0.756 | 0.03 |
| MADRS | **0.16 (0.12-0.2)** | **<.001** | 0.07 | **0.16 (0.12-0.2)** | **<.001** | 0.07 |
| History of psychosis | -0.14 (-0.44-0.15) | 0.347 | 0.21 | -0.14 (-0.43-0.15) | 0.345 | 0.15 |
| Antidepressants | 0.21 (-0.12-0.53) | 0.222 | 0.05 | 0.29 (-0.01-0.6) | 0.057 | 0.03 |
| Lithium | 0.07 (-0.27-0.41) | 0.683 | 0.04 | 0.16 (-0.14-0.47) | 0.296 | 0.03 |
| Antipsychotics | 0.11 (-0.15-0.36) | 0.413 | 0.06 | 0.11 (-0.14-0.36) | 0.386 | 0.05 |
| Anxiolytics | **0.43 (0.12-0.73)** | **0.006** | 0.06 | **0.47 (0.18-0.76)** | **0.002** | 0.05 |
| Antiparkisonian | 0.41 (-0.55-1.37) | 0.407 | 0.01 | 0.45 (-0.5-1.41) | 0.353 | 0.01 |
| Education level | **-0.05 (-0.1-0)** | **0.031** | 0.05 | **-0.05 (-0.1-0)** | **0.034** | 0.05 |
| Age at first episode | 0.01 (-0.01-0.02) | 0.324 | 0.11 | 0.01 (-0.01-0.02) | 0.417 | 0.11 |
| Number of manic episode | -0.05 (-0.12-0.01) | 0.094 | 0.11 | -0.06 (-0.12-0.01) | 0.081 | 0.11 |
| Number of depressive episode | 0.01 (-0.01-0.03) | 0.283 | 0.27 | 0.01 (-0.01-0.03) | 0.269 | 0.24 |
| CGI | 0.03 (-0.06-0.12) | 0.523 | 0.09 | 0.03 (-0.06-0.11) | 0.561 | 0.08 |

[1] R2 = 0.09; adj R2 = 0.081; AIC = 6341
[2] R2 = 0.088; adj R2 = 0.08; AIC = 6339

| Var | ARS (sum)[1] | | | ARS (max)[2] | | |
| --- | --- | --- | --- | --- | --- | --- |
| | Beta (95% CI) | p-value | fmi | Beta (95% CI) | p-value | fmi |
| Scale | **0.26 (0.09-0.44)** | **0.003** | 0.07 | **0.3 (0.11-0.49)** | **0.002** | 0.05 |
| MADRS | **0.16 (0.12-0.2)** | **<.001** | 0.07 | **0.16 (0.12-0.2)** | **<.001** | 0.07 |
| History of psychosis | -0.12 (-0.42-0.18) | 0.42 | 0.21 | -0.12 (-0.41-0.17) | 0.423 | 0.16 |
| Antidepressants | **0.28 (0.01-0.55)** | **0.04** | 0.06 | **0.29 (0.03-0.55)** | **0.032** | 0.04 |
| Lithium | 0.2 (-0.06-0.45) | 0.131 | 0.05 | 0.19 (-0.06-0.45) | 0.143 | 0.04 |
| Antipsychotics | -0.1 (-0.4-0.19) | 0.495 | 0.08 | -0.13 (-0.42-0.17) | 0.398 | 0.05 |
| Anxiolytics | **0.41 (0.12-0.71)** | **0.006** | 0.07 | **0.41 (0.12-0.7)** | **0.005** | 0.05 |
| Antiparkisonian | 0.51 (-0.44-1.46) | 0.295 | 0.01 | 0.52 (-0.43-1.47) | 0.287 | 0.01 |
| Education level | **-0.05 (-0.1-0)** | **0.036** | 0.05 | **-0.05 (-0.1-0)** | **0.037** | 0.05 |
| Age at first episode | 0.01 (-0.01-0.02) | 0.351 | 0.11 | 0.01 (-0.01-0.02) | 0.411 | 0.11 |
| Number of manic episode | -0.05 (-0.12-0.01) | 0.091 | 0.11 | -0.06 (-0.12-0.01) | 0.081 | 0.11 |
| Number of depressive episode | 0.01 (-0.01-0.04) | 0.229 | 0.27 | 0.01 (-0.01-0.04) | 0.225 | 0.24 |
| CGI | 0.03 (-0.06-0.12) | 0.533 | 0.09 | 0.03 (-0.06-0.11) | 0.572 | 0.07 |

[1] R2 = 0.095; adj R2 = 0.086; AIC = 6333
[2] R2 = 0.094; adj R2 = 0.086; AIC = 6329

| Var | BADS (sum)[1] | | | BADS (max)[2] | | |
| --- | --- | --- | --- | --- | --- | --- |
| | Beta (95% CI) | p-value | fmi | Beta (95% CI) | p-value | fmi |
| Scale | **0.13 (0.03-0.23)** | **0.014** | 0.04 | **0.19 (0.04-0.33)** | **0.011** | 0.04 |
| MADRS | **0.16 (0.12-0.2)** | **<.001** | 0.07 | **0.16 (0.12-0.2)** | **<.001** | 0.07 |
| History of psychosis | -0.13 (-0.43-0.17) | 0.406 | 0.21 | -0.13 (-0.41-0.16) | 0.393 | 0.15 |
| Antidepressants | 0.17 (-0.12-0.46) | 0.252 | 0.05 | 0.23 (-0.04-0.5) | 0.091 | 0.04 |
| Lithium | 0.11 (-0.16-0.38) | 0.417 | 0.04 | 0.16 (-0.1-0.41) | 0.228 | 0.04 |
| Antipsychotics | -0.03 (-0.32-0.25) | 0.812 | 0.06 | -0.01 (-0.28-0.26) | 0.931 | 0.05 |
| Anxiolytics | **0.41 (0.12-0.7)** | **0.006** | 0.06 | **0.45 (0.16-0.73)** | **0.002** | 0.05 |
| Antiparkisonian | 0.33 (-0.63-1.29) | 0.505 | 0.01 | 0.38 (-0.57-1.33) | 0.435 | 0.01 |
| Education level | **-0.05 (-0.1-0)** | **0.033** | 0.05 | **-0.05 (-0.1-0)** | **0.031** | 0.05 |
| Age at first episode | 0.01 (-0.01-0.02) | 0.396 | 0.11 | 0.01 (-0.01-0.02) | 0.469 | 0.11 |
| Number of manic episode | -0.06 (-0.12-0.01) | 0.075 | 0.11 | -0.06 (-0.12-0) | 0.07 | 0.11 |
| Number of depressive episode | 0.01 (-0.01-0.04) | 0.24 | 0.27 | 0.01 (-0.01-0.03) | 0.245 | 0.24 |
| CGI | 0.02 (-0.06-0.11) | 0.578 | 0.09 | 0.02 (-0.07-0.11) | 0.62 | 0.08 |

[1] R2 = 0.093; adj R2 = 0.084; AIC = 6336
[2] R2 = 0.092; adj R2 = 0.084; AIC = 6332

| Var | CALS (sum)[1] | | | CALS (max)[2] | | |
| --- | --- | --- | --- | --- | --- | --- |
| | Beta (95% CI) | p-value | fmi | Beta (95% CI) | p-value | fmi |
| Scale | **0.19 (0.06-0.31)** | **0.004** | 0.05 | **0.25 (0.06-0.45)** | **0.012** | 0.04 |
| MADRS | **0.16 (0.12-0.2)** | **<.001** | 0.07 | **0.16 (0.12-0.2)** | **<.001** | 0.07 |
| History of psychosis | -0.13 (-0.43-0.17) | 0.386 | 0.21 | -0.13 (-0.42-0.15) | 0.367 | 0.15 |
| Antidepressants | 0.12 (-0.18-0.42) | 0.431 | 0.05 | 0.25 (-0.02-0.52) | 0.069 | 0.04 |
| Lithium | 0.08 (-0.19-0.35) | 0.551 | 0.04 | 0.17 (-0.09-0.43) | 0.195 | 0.04 |
| Antipsychotics | -0.15 (-0.47-0.16) | 0.333 | 0.05 | -0.06 (-0.34-0.23) | 0.701 | 0.05 |
| Anxiolytics | **0.32 (0.02-0.63)** | **0.038** | 0.06 | **0.43 (0.14-0.72)** | **0.004** | 0.05 |
| Antiparkisonian | -0.12 (-1.16-0.91) | 0.813 | 0.02 | 0.12 (-0.87-1.11) | 0.809 | 0.01 |
| Education level | **-0.05 (-0.1--0.01)** | **0.028** | 0.05 | **-0.05 (-0.1-0)** | **0.03** | 0.05 |
| Age at first episode | 0.01 (-0.01-0.02) | 0.426 | 0.12 | 0 (-0.01-0.02) | 0.485 | 0.11 |
| Number of manic episode | -0.06 (-0.12-0) | 0.066 | 0.12 | -0.06 (-0.12-0.01) | 0.074 | 0.10 |
| Number of depressive episode | 0.01 (-0.01-0.04) | 0.244 | 0.27 | 0.01 (-0.01-0.03) | 0.265 | 0.24 |
| CGI | 0.03 (-0.06-0.11) | 0.574 | 0.08 | 0.02 (-0.06-0.11) | 0.583 | 0.08 |

[1] R2 = 0.094; adj R2 = 0.086; AIC = 6333
[2] R2 = 0.092; adj R2 = 0.084; AIC = 6332

| Var | CrAS (sum)[1] | | | CrAS (max)[2] | | |
| --- | --- | --- | --- | --- | --- | --- |
| | Beta (95% CI) | p-value | fmi | Beta (95% CI) | p-value | fmi |
| Scale | 0.14 (-0.01-0.29) | 0.069 | 0.04 | 0.17 (-0.01-0.34) | 0.059 | 0.04 |
| MADRS | **0.16 (0.12-0.2)** | **<.001** | 0.07 | **0.16 (0.12-0.2)** | **<.001** | 0.07 |
| History of psychosis | -0.13 (-0.43-0.17) | 0.388 | 0.21 | -0.12 (-0.41-0.17) | 0.402 | 0.16 |
| Antidepressants | 0.22 (-0.06-0.51) | 0.128 | 0.05 | 0.24 (-0.03-0.52) | 0.082 | 0.04 |
| Lithium | 0.19 (-0.06-0.45) | 0.137 | 0.05 | 0.19 (-0.06-0.45) | 0.143 | 0.04 |
| Antipsychotics | -0.02 (-0.32-0.28) | 0.891 | 0.07 | -0.03 (-0.32-0.26) | 0.854 | 0.05 |
| Anxiolytics | **0.45 (0.16-0.74)** | **0.002** | 0.06 | **0.46 (0.17-0.75)** | **0.002** | 0.05 |
| Antiparkisonian | 0.37 (-0.59-1.33) | 0.453 | 0.01 | 0.37 (-0.58-1.33) | 0.444 | 0.01 |
| Education level | **-0.05 (-0.1-0)** | **0.031** | 0.05 | **-0.05 (-0.1-0)** | **0.031** | 0.05 |
| Age at first episode | 0.01 (-0.01-0.02) | 0.337 | 0.11 | 0.01 (-0.01-0.02) | 0.403 | 0.11 |
| Number of manic episode | -0.05 (-0.11-0.01) | 0.115 | 0.11 | -0.05 (-0.11-0.01) | 0.105 | 0.10 |
| Number of depressive episode | 0.01 (-0.01-0.03) | 0.28 | 0.27 | 0.01 (-0.01-0.03) | 0.268 | 0.24 |
| CGI | 0.03 (-0.06-0.12) | 0.534 | 0.09 | 0.03 (-0.06-0.11) | 0.574 | 0.08 |

[1] R2 = 0.091; adj R2 = 0.083; AIC = 6339
[2] R2 = 0.09; adj R2 = 0.082; AIC = 6335

| Var | DBI-WHO (sum)[1] | | | DBI-WHO (max)[2] | | |
| --- | --- | --- | --- | --- | --- | --- |
| | Beta (95% CI) | p-value | fmi | Beta (95% CI) | p-value | fmi |
| Scale | 0.38 (-0.01-0.77) | 0.054 | 0.05 | 0.27 (-0.48-1.03) | 0.478 | 0.05 |
| MADRS | **0.16 (0.12-0.2)** | **<.001** | 0.07 | **0.16 (0.12-0.2)** | **<.001** | 0.07 |
| History of psychosis | -0.14 (-0.43-0.16) | 0.374 | 0.21 | -0.14 (-0.43-0.15) | 0.334 | 0.15 |
| Antidepressants | 0.17 (-0.13-0.47) | 0.275 | 0.05 | **0.29 (0.01-0.56)** | **0.045** | 0.04 |
| Lithium | 0.06 (-0.24-0.35) | 0.705 | 0.04 | 0.15 (-0.12-0.43) | 0.278 | 0.03 |
| Antipsychotics | 0 (-0.28-0.28) | 0.986 | 0.05 | 0.09 (-0.16-0.35) | 0.474 | 0.05 |
| Anxiolytics | **0.4 (0.1-0.7)** | **0.009** | 0.05 | **0.47 (0.18-0.75)** | **0.002** | 0.05 |
| Antiparkisonian | 0.38 (-0.58-1.34) | 0.441 | 0.01 | 0.46 (-0.5-1.41) | 0.347 | 0.01 |
| Education level | **-0.05 (-0.1-0)** | **0.033** | 0.05 | **-0.05 (-0.1-0)** | **0.035** | 0.05 |
| Age at first episode | 0.01 (-0.01-0.02) | 0.378 | 0.11 | 0.01 (-0.01-0.02) | 0.419 | 0.11 |
| Number of manic episode | -0.06 (-0.12-0) | 0.071 | 0.11 | -0.06 (-0.12-0.01) | 0.077 | 0.11 |
| Number of depressive episode | 0.01 (-0.01-0.03) | 0.293 | 0.27 | 0.01 (-0.01-0.03) | 0.273 | 0.24 |
| CGI | 0.03 (-0.06-0.12) | 0.553 | 0.09 | 0.03 (-0.06-0.11) | 0.561 | 0.08 |

[1] R2 = 0.091; adj R2 = 0.083; AIC = 6338
[2] R2 = 0.088; adj R2 = 0.08; AIC = 6338

| Var | DRS (sum)[1] | | | DRS (max)[2] | | |
| --- | --- | --- | --- | --- | --- | --- |
| | Beta (95% CI) | p-value | fmi | Beta (95% CI) | p-value | fmi |
| Scale | 0.05 (-0.04-0.14) | 0.26 | 0.03 | **0.14 (0.01-0.27)** | **0.037** | 0.03 |
| MADRS | **0.16 (0.12-0.2)** | **<.001** | 0.07 | **0.16 (0.12-0.2)** | **<.001** | 0.07 |
| History of psychosis | -0.13 (-0.43-0.16) | 0.379 | 0.21 | -0.13 (-0.42-0.16) | 0.372 | 0.15 |
| Antidepressants | 0.25 (-0.04-0.54) | 0.093 | 0.05 | 0.24 (-0.04-0.51) | 0.089 | 0.04 |
| Lithium | 0.12 (-0.17-0.41) | 0.429 | 0.04 | 0.08 (-0.19-0.36) | 0.563 | 0.04 |
| Antipsychotics | 0.07 (-0.21-0.34) | 0.639 | 0.06 | 0.05 (-0.21-0.31) | 0.709 | 0.05 |
| Anxiolytics | **0.41 (0.1-0.72)** | **0.01** | 0.05 | **0.42 (0.13-0.71)** | **0.004** | 0.04 |
| Antiparkisonian | 0.4 (-0.56-1.36) | 0.414 | 0.01 | 0.35 (-0.61-1.3) | 0.481 | 0.01 |
| Education level | **-0.05 (-0.1-0)** | **0.031** | 0.05 | **-0.05 (-0.1-0)** | **0.033** | 0.05 |
| Age at first episode | 0.01 (-0.01-0.02) | 0.365 | 0.11 | 0.01 (-0.01-0.02) | 0.464 | 0.11 |
| Number of manic episode | -0.06 (-0.12-0.01) | 0.083 | 0.12 | -0.06 (-0.12-0) | 0.07 | 0.11 |
| Number of depressive episode | 0.01 (-0.01-0.03) | 0.255 | 0.27 | 0.01 (-0.01-0.03) | 0.249 | 0.24 |
| CGI | 0.03 (-0.06-0.12) | 0.525 | 0.09 | 0.02 (-0.06-0.11) | 0.581 | 0.08 |

[1] R2 = 0.09; adj R2 = 0.081; AIC = 6341
[2] R2 = 0.091; adj R2 = 0.082; AIC = 6334

| Var | Duran (sum)[1] | | | Duran (max)[2] | | |
| --- | --- | --- | --- | --- | --- | --- |
| | Beta (95% CI) | p-value | fmi | Beta (95% CI) | p-value | fmi |
| Scale | **0.18 (0.08-0.28)** | **<.001** | 0.03 | **0.25 (0.09-0.41)** | **0.002** | 0.03 |
| MADRS | **0.16 (0.12-0.2)** | **<.001** | 0.07 | **0.16 (0.12-0.2)** | **<.001** | 0.07 |
| History of psychosis | -0.12 (-0.42-0.17) | 0.419 | 0.20 | -0.13 (-0.41-0.16) | 0.383 | 0.15 |
| Antidepressants | 0.18 (-0.1-0.45) | 0.207 | 0.05 | 0.24 (-0.02-0.51) | 0.073 | 0.04 |
| Lithium | -0.13 (-0.44-0.19) | 0.424 | 0.04 | -0.05 (-0.34-0.25) | 0.753 | 0.04 |
| Antipsychotics | -0.11 (-0.39-0.18) | 0.451 | 0.06 | -0.01 (-0.27-0.25) | 0.935 | 0.05 |
| Anxiolytics | 0.28 (-0.03-0.59) | 0.08 | 0.06 | **0.39 (0.1-0.68)** | **0.009** | 0.05 |
| Antiparkisonian | -0.09 (-1.09-0.91) | 0.862 | 0.01 | 0.11 (-0.86-1.09) | 0.82 | 0.01 |
| Education level | **-0.05 (-0.1--0.01)** | **0.023** | 0.05 | **-0.05 (-0.1--0.01)** | **0.026** | 0.05 |
| Age at first episode | 0.01 (-0.01-0.02) | 0.382 | 0.12 | 0.01 (-0.01-0.02) | 0.431 | 0.11 |
| Number of manic episode | -0.06 (-0.12-0.01) | 0.076 | 0.11 | -0.05 (-0.12-0.01) | 0.086 | 0.10 |
| Number of depressive episode | 0.01 (-0.01-0.04) | 0.234 | 0.27 | 0.01 (-0.01-0.03) | 0.245 | 0.24 |
| CGI | 0.03 (-0.06-0.12) | 0.53 | 0.08 | 0.02 (-0.06-0.11) | 0.578 | 0.08 |

[1] R2 = 0.097; adj R2 = 0.088; AIC = 6330
[2] R2 = 0.094; adj R2 = 0.086; AIC = 6329

| Var | GACB (sum)[1] | | | GACB (max)[2] | | |
| --- | --- | --- | --- | --- | --- | --- |
| | Beta (95% CI) | p-value | fmi | Beta (95% CI) | p-value | fmi |
| Scale | **0.15 (0.03-0.27)** | **0.017** | 0.03 | **0.23 (0.04-0.42)** | **0.016** | 0.05 |
| MADRS | **0.16 (0.12-0.2)** | **<.001** | 0.07 | **0.16 (0.12-0.2)** | **<.001** | 0.07 |
| History of psychosis | -0.14 (-0.43-0.16) | 0.369 | 0.21 | -0.13 (-0.42-0.15) | 0.36 | 0.15 |
| Antidepressants | 0.16 (-0.13-0.46) | 0.28 | 0.05 | 0.25 (-0.01-0.52) | 0.063 | 0.04 |
| Lithium | 0.11 (-0.16-0.37) | 0.428 | 0.05 | 0.17 (-0.09-0.42) | 0.196 | 0.04 |
| Antipsychotics | -0.09 (-0.39-0.22) | 0.581 | 0.05 | -0.04 (-0.32-0.24) | 0.795 | 0.05 |
| Anxiolytics | **0.38 (0.08-0.68)** | **0.014** | 0.06 | **0.44 (0.15-0.72)** | **0.003** | 0.05 |
| Antiparkisonian | 0.26 (-0.71-1.23) | 0.596 | 0.01 | 0.35 (-0.6-1.31) | 0.469 | 0.01 |
| Education level | **-0.05 (-0.1-0)** | **0.03** | 0.05 | **-0.05 (-0.1-0)** | **0.032** | 0.05 |
| Age at first episode | 0.01 (-0.01-0.02) | 0.404 | 0.12 | 0 (-0.01-0.02) | 0.493 | 0.11 |
| Number of manic episode | -0.06 (-0.12-0) | 0.069 | 0.12 | -0.06 (-0.12-0.01) | 0.072 | 0.11 |
| Number of depressive episode | 0.01 (-0.01-0.03) | 0.253 | 0.27 | 0.01 (-0.01-0.03) | 0.262 | 0.24 |
| CGI | 0.03 (-0.06-0.11) | 0.551 | 0.09 | 0.03 (-0.06-0.11) | 0.573 | 0.08 |

[1] R2 = 0.093; adj R2 = 0.084; AIC = 6336
[2] R2 = 0.092; adj R2 = 0.083; AIC = 6333

| Var | KABS (sum)[1] | | | KABS (max)[2] | | |
| --- | --- | --- | --- | --- | --- | --- |
| | Beta (95% CI) | p-value | fmi | Beta (95% CI) | p-value | fmi |
| Scale | **0.21 (0.09-0.34)** | **0.001** | 0.06 | **0.26 (0.1-0.42)** | **0.002** | 0.04 |
| MADRS | **0.16 (0.12-0.2)** | **<.001** | 0.07 | **0.16 (0.12-0.2)** | **<.001** | 0.07 |
| History of psychosis | -0.13 (-0.43-0.17) | 0.397 | 0.21 | -0.12 (-0.41-0.16) | 0.398 | 0.15 |
| Antidepressants | 0.1 (-0.19-0.4) | 0.487 | 0.06 | 0.17 (-0.11-0.45) | 0.232 | 0.04 |
| Lithium | 0.21 (-0.04-0.47) | 0.106 | 0.05 | 0.21 (-0.05-0.46) | 0.11 | 0.04 |
| Antipsychotics | -0.25 (-0.58-0.09) | 0.149 | 0.06 | -0.22 (-0.54-0.11) | 0.192 | 0.05 |
| Anxiolytics | **0.33 (0.03-0.63)** | **0.033** | 0.07 | **0.4 (0.11-0.69)** | **0.007** | 0.05 |
| Antiparkisonian | 0.26 (-0.7-1.22) | 0.596 | 0.01 | 0.36 (-0.59-1.32) | 0.456 | 0.01 |
| Education level | **-0.05 (-0.1-0)** | **0.032** | 0.05 | **-0.05 (-0.1-0)** | **0.035** | 0.05 |
| Age at first episode | 0.01 (-0.01-0.02) | 0.411 | 0.11 | 0.01 (-0.01-0.02) | 0.468 | 0.11 |
| Number of manic episode | -0.06 (-0.12-0.01) | 0.086 | 0.11 | -0.06 (-0.12-0.01) | 0.085 | 0.11 |
| Number of depressive episode | 0.01 (-0.01-0.03) | 0.247 | 0.27 | 0.01 (-0.01-0.03) | 0.241 | 0.24 |
| CGI | 0.03 (-0.06-0.11) | 0.554 | 0.08 | 0.02 (-0.06-0.11) | 0.613 | 0.08 |

[1] R2 = 0.096; adj R2 = 0.088; AIC = 6331
[2] R2 = 0.095; adj R2 = 0.086; AIC = 6328

| Var | mACB1 (sum)[1] | | | mACB1 (max)[2] | | |
| --- | --- | --- | --- | --- | --- | --- |
| | Beta (95% CI) | p-value | fmi | Beta (95% CI) | p-value | fmi |
| Scale | 0.09 (-0.02-0.19) | 0.095 | 0.04 | 0.12 (0-0.25) | 0.052 | 0.04 |
| MADRS | **0.16 (0.12-0.2)** | **<.001** | 0.07 | **0.16 (0.12-0.2)** | **<.001** | 0.07 |
| History of psychosis | -0.13 (-0.43-0.17) | 0.382 | 0.21 | -0.13 (-0.41-0.16) | 0.393 | 0.15 |
| Antidepressants | 0.23 (-0.06-0.51) | 0.116 | 0.05 | 0.25 (-0.03-0.52) | 0.077 | 0.04 |
| Lithium | 0.21 (-0.05-0.46) | 0.116 | 0.05 | 0.2 (-0.05-0.46) | 0.117 | 0.04 |
| Antipsychotics | -0.06 (-0.39-0.27) | 0.741 | 0.06 | -0.09 (-0.41-0.24) | 0.6 | 0.05 |
| Anxiolytics | **0.47 (0.18-0.76)** | **0.002** | 0.06 | **0.47 (0.19-0.76)** | **0.001** | 0.05 |
| Antiparkisonian | 0.41 (-0.55-1.36) | 0.407 | 0.01 | 0.44 (-0.51-1.4) | 0.362 | 0.01 |
| Education level | **-0.05 (-0.1-0)** | **0.03** | 0.05 | **-0.05 (-0.1-0)** | **0.031** | 0.05 |
| Age at first episode | 0.01 (-0.01-0.02) | 0.365 | 0.11 | 0.01 (-0.01-0.02) | 0.433 | 0.11 |
| Number of manic episode | -0.05 (-0.12-0.01) | 0.091 | 0.11 | -0.06 (-0.12-0.01) | 0.082 | 0.11 |
| Number of depressive episode | 0.01 (-0.01-0.03) | 0.268 | 0.27 | 0.01 (-0.01-0.03) | 0.262 | 0.24 |
| CGI | 0.03 (-0.06-0.12) | 0.544 | 0.09 | 0.02 (-0.06-0.11) | 0.607 | 0.08 |

[1] R2 = 0.091; adj R2 = 0.082; AIC = 6339
[2] R2 = 0.09; adj R2 = 0.082; AIC = 6335

| Var | mACB2 (sum)[1] | | | mACB2 (max)[2] | | |
| --- | --- | --- | --- | --- | --- | --- |
| | Beta (95% CI) | p-value | fmi | Beta (95% CI) | p-value | fmi |
| Scale | **0.15 (0.04-0.25)** | **0.005** | 0.04 | **0.18 (0.05-0.32)** | **0.007** | 0.05 |
| MADRS | **0.16 (0.12-0.2)** | **<.001** | 0.07 | **0.16 (0.12-0.2)** | **<.001** | 0.07 |
| History of psychosis | -0.12 (-0.42-0.17) | 0.418 | 0.21 | -0.12 (-0.4-0.17) | 0.428 | 0.15 |
| Antidepressants | 0.14 (-0.15-0.44) | 0.332 | 0.05 | 0.23 (-0.04-0.5) | 0.102 | 0.04 |
| Lithium | 0.25 (0-0.51) | 0.055 | 0.05 | 0.24 (-0.02-0.49) | 0.071 | 0.04 |
| Antipsychotics | -0.15 (-0.48-0.17) | 0.346 | 0.06 | -0.13 (-0.43-0.18) | 0.418 | 0.06 |
| Anxiolytics | **0.37 (0.07-0.67)** | **0.015** | 0.06 | **0.44 (0.15-0.73)** | **0.003** | 0.05 |
| Antiparkisonian | 0.31 (-0.65-1.27) | 0.524 | 0.01 | 0.44 (-0.52-1.39) | 0.369 | 0.01 |
| Education level | **-0.05 (-0.1-0)** | **0.032** | 0.05 | **-0.05 (-0.1-0)** | **0.033** | 0.05 |
| Age at first episode | 0.01 (-0.01-0.02) | 0.389 | 0.11 | 0.01 (-0.01-0.02) | 0.466 | 0.11 |
| Number of manic episode | -0.06 (-0.12-0.01) | 0.081 | 0.11 | -0.06 (-0.12-0.01) | 0.082 | 0.10 |
| Number of depressive episode | 0.01 (-0.01-0.04) | 0.247 | 0.27 | 0.01 (-0.01-0.03) | 0.245 | 0.24 |
| CGI | 0.03 (-0.06-0.11) | 0.555 | 0.09 | 0.02 (-0.06-0.11) | 0.607 | 0.08 |

[1] R2 = 0.094; adj R2 = 0.086; AIC = 6334
[2] R2 = 0.093; adj R2 = 0.084; AIC = 6331

| Var | Marante (sum)[1] | | | Marante (max)[2] | | |
| --- | --- | --- | --- | --- | --- | --- |
| | Beta (95% CI) | p-value | fmi | Beta (95% CI) | p-value | fmi |
| Scale | 0.09 (-0.03-0.21) | 0.123 | 0.03 | **0.16 (0.01-0.32)** | **0.04** | 0.02 |
| MADRS | **0.16 (0.12-0.2)** | **<.001** | 0.07 | **0.16 (0.12-0.2)** | **<.001** | 0.07 |
| History of psychosis | -0.13 (-0.43-0.17) | 0.386 | 0.21 | -0.13 (-0.42-0.16) | 0.372 | 0.15 |
| Antidepressants | 0.24 (-0.04-0.52) | 0.097 | 0.05 | 0.24 (-0.04-0.51) | 0.09 | 0.04 |
| Lithium | 0.11 (-0.17-0.39) | 0.457 | 0.04 | 0.11 (-0.16-0.37) | 0.434 | 0.04 |
| Antipsychotics | 0.02 (-0.26-0.31) | 0.887 | 0.06 | 0.01 (-0.27-0.28) | 0.964 | 0.05 |
| Anxiolytics | **0.42 (0.13-0.72)** | **0.005** | 0.06 | **0.43 (0.14-0.72)** | **0.004** | 0.04 |
| Antiparkisonian | 0.37 (-0.59-1.33) | 0.451 | 0.01 | 0.35 (-0.61-1.31) | 0.477 | 0.01 |
| Education level | **-0.05 (-0.1-0)** | **0.032** | 0.05 | **-0.05 (-0.1-0)** | **0.035** | 0.05 |
| Age at first episode | 0.01 (-0.01-0.02) | 0.359 | 0.11 | 0.01 (-0.01-0.02) | 0.432 | 0.11 |
| Number of manic episode | -0.06 (-0.12-0.01) | 0.085 | 0.11 | -0.06 (-0.12-0.01) | 0.076 | 0.11 |
| Number of depressive episode | 0.01 (-0.01-0.03) | 0.257 | 0.27 | 0.01 (-0.01-0.03) | 0.254 | 0.24 |
| CGI | 0.03 (-0.06-0.12) | 0.511 | 0.09 | 0.03 (-0.06-0.12) | 0.539 | 0.08 |

[1] R2 = 0.09; adj R2 = 0.082; AIC = 6340
[2] R2 = 0.091; adj R2 = 0.082; AIC = 6334

| Var | mARS (sum)[1] | | | mARS (max)[2] | | |
| --- | --- | --- | --- | --- | --- | --- |
| | Beta (95% CI) | p-value | fmi | Beta (95% CI) | p-value | fmi |
| Scale | **0.24 (0.07-0.4)** | **0.005** | 0.06 | **0.31 (0.12-0.49)** | **0.001** | 0.05 |
| MADRS | **0.16 (0.12-0.2)** | **<.001** | 0.07 | **0.16 (0.12-0.2)** | **<.001** | 0.07 |
| History of psychosis | -0.12 (-0.42-0.18) | 0.418 | 0.21 | -0.12 (-0.4-0.17) | 0.428 | 0.16 |
| Antidepressants | 0.26 (-0.01-0.52) | 0.062 | 0.06 | 0.26 (-0.01-0.52) | 0.056 | 0.04 |
| Lithium | 0.2 (-0.05-0.46) | 0.121 | 0.05 | 0.19 (-0.06-0.45) | 0.134 | 0.04 |
| Antipsychotics | -0.08 (-0.38-0.21) | 0.574 | 0.08 | -0.13 (-0.42-0.16) | 0.377 | 0.05 |
| Anxiolytics | **0.41 (0.12-0.71)** | **0.006** | 0.07 | **0.4 (0.11-0.69)** | **0.007** | 0.05 |
| Antiparkisonian | 0.52 (-0.44-1.47) | 0.287 | 0.01 | 0.53 (-0.42-1.48) | 0.277 | 0.01 |
| Education level | **-0.05 (-0.1-0)** | **0.033** | 0.05 | **-0.05 (-0.1-0)** | **0.033** | 0.05 |
| Age at first episode | 0.01 (-0.01-0.02) | 0.399 | 0.11 | 0.01 (-0.01-0.02) | 0.467 | 0.11 |
| Number of manic episode | -0.06 (-0.12-0.01) | 0.084 | 0.11 | -0.06 (-0.12-0.01) | 0.077 | 0.11 |
| Number of depressive episode | 0.01 (-0.01-0.04) | 0.247 | 0.27 | 0.01 (-0.01-0.03) | 0.249 | 0.24 |
| CGI | 0.03 (-0.06-0.12) | 0.53 | 0.09 | 0.02 (-0.06-0.11) | 0.576 | 0.08 |

[1] R2 = 0.094; adj R2 = 0.086; AIC = 6334
[2] R2 = 0.095; adj R2 = 0.087; AIC = 6327

| Var | Salahudeen (sum)[1] | | | Salahudeen (max)[2] | | |
| --- | --- | --- | --- | --- | --- | --- |
| | Beta (95% CI) | p-value | fmi | Beta (95% CI) | p-value | fmi |
| Scale | **0.23 (0.09-0.38)** | **0.001** | 0.05 | **0.3 (0.06-0.54)** | **0.013** | 0.06 |
| MADRS | **0.16 (0.12-0.2)** | **<.001** | 0.07 | **0.16 (0.12-0.2)** | **<.001** | 0.07 |
| History of psychosis | -0.14 (-0.44-0.16) | 0.354 | 0.21 | -0.15 (-0.43-0.14) | 0.32 | 0.15 |
| Antidepressants | 0.09 (-0.21-0.39) | 0.56 | 0.05 | 0.25 (-0.02-0.51) | 0.073 | 0.04 |
| Lithium | 0.05 (-0.22-0.32) | 0.717 | 0.05 | 0.16 (-0.1-0.41) | 0.231 | 0.04 |
| Antipsychotics | -0.09 (-0.37-0.2) | 0.546 | 0.06 | 0.05 (-0.2-0.31) | 0.678 | 0.05 |
| Anxiolytics | 0.28 (-0.04-0.59) | 0.084 | 0.06 | **0.4 (0.11-0.69)** | **0.007** | 0.05 |
| Antiparkisonian | -0.31 (-1.38-0.76) | 0.569 | 0.02 | -0.09 (-1.14-0.96) | 0.869 | 0.02 |
| Education level | **-0.05 (-0.1--0.01)** | **0.028** | 0.05 | **-0.05 (-0.1-0)** | **0.033** | 0.05 |
| Age at first episode | 0.01 (-0.01-0.02) | 0.44 | 0.12 | 0 (-0.01-0.02) | 0.517 | 0.11 |
| Number of manic episode | -0.06 (-0.13-0) | 0.055 | 0.12 | -0.06 (-0.12-0) | 0.067 | 0.11 |
| Number of depressive episode | 0.01 (-0.01-0.03) | 0.252 | 0.27 | 0.01 (-0.01-0.03) | 0.279 | 0.24 |
| CGI | 0.02 (-0.06-0.11) | 0.588 | 0.08 | 0.02 (-0.06-0.11) | 0.592 | 0.08 |

[1] R2 = 0.096; adj R2 = 0.087; AIC = 6332
[2] R2 = 0.092; adj R2 = 0.084; AIC = 6332

| Var | Summers (sum)[1] | | | Summers (max)[2] | | |
| --- | --- | --- | --- | --- | --- | --- |
| | Beta (95% CI) | p-value | fmi | Beta (95% CI) | p-value | fmi |
| Scale | 0 (-0.13-0.14) | 0.961 | 0.25 | -0.01 (-0.14-0.12) | 0.904 | 0.06 |
| MADRS | **0.16 (0.12-0.2)** | **<.001** | 0.07 | **0.16 (0.12-0.2)** | **<.001** | 0.07 |
| History of psychosis | -0.14 (-0.44-0.16) | 0.351 | 0.21 | -0.14 (-0.43-0.15) | 0.347 | 0.15 |
| Antidepressants | **0.32 (0.05-0.58)** | **0.02** | 0.06 | **0.32 (0.06-0.58)** | **0.018** | 0.04 |
| Lithium | 0.2 (-0.06-0.45) | 0.131 | 0.04 | 0.19 (-0.06-0.45) | 0.14 | 0.04 |
| Antipsychotics | 0.12 (-0.13-0.38) | 0.354 | 0.06 | 0.11 (-0.14-0.37) | 0.384 | 0.05 |
| Anxiolytics | **0.47 (0.18-0.77)** | **0.002** | 0.06 | **0.48 (0.18-0.77)** | **0.002** | 0.05 |
| Antiparkisonian | 0.46 (-0.55-1.46) | 0.374 | 0.03 | 0.48 (-0.51-1.47) | 0.34 | 0.01 |
| Education level | **-0.05 (-0.1-0)** | **0.031** | 0.05 | **-0.05 (-0.1-0)** | **0.033** | 0.05 |
| Age at first episode | 0.01 (-0.01-0.02) | 0.346 | 0.11 | 0.01 (-0.01-0.02) | 0.415 | 0.11 |
| Number of manic episode | -0.05 (-0.12-0.01) | 0.091 | 0.11 | -0.06 (-0.12-0.01) | 0.081 | 0.11 |
| Number of depressive episode | 0.01 (-0.01-0.03) | 0.277 | 0.27 | 0.01 (-0.01-0.03) | 0.268 | 0.24 |
| CGI | 0.03 (-0.06-0.12) | 0.524 | 0.09 | 0.03 (-0.06-0.11) | 0.561 | 0.08 |

[1] R2 = 0.089; adj R2 = 0.081; AIC = 6342
[2] R2 = 0.088; adj R2 = 0.08; AIC = 6339

**Supplementary Table 4. Multiple logistic regression models of peripheral anticholinergic side effects subscore with the 21 scales significantly associated with the peripheral subscore in bivariable regression analysis as main predictor.**

**Significant associations are in bold. ABC: Anticholinergic Burden Classification; ACB: Anticholinergic Cognitive Burden Scale; ADS: Anticholinergic Drug Scale; AEC: Anticholinergic Effect on Cognition; AIS: Anticholinergic Impregnation Scale; ALS: Anticholinergic Loading Scale; ARS: Anticholminergic Risk Scale; BADS: Brazilian Anticholinergic Activity Drug Scale; CALS: the CRIDECO Anticholinergic Load Scale; Chew: Chew's scale; CrAS: Clinician-rated Anticholinergic Scale; DBI-WHO: Drug Burden Index, WHO version; DRS: Delirogenic Risk Scale; Duran: Durán Scale; GACB: German Anticholinergic Burden Scale; KABS: Korean Anticholinergic Burden Scale; mACB1: modified Anticholinergic Cognitive Burden scale 1; mACB2: modified Anticholinergic Cognitive Burden scale 2; Marante: Muscarinic Acetylcholinergic Receptor ANTagonist Exposure Scale; mARS: modified Anticholinergic Risk Scale; Salahudeen: Salahudeen's scale; fmi: fraction of missing information.**

| | AAS (sum)[1] | | | AAS (max)[2] | | |
|---|---|---|---|---|---|---|
| Var | Beta (95% CI) | p-value | fmi | Beta (95% CI) | p-value | fmi |
| Scale | 0.09 (-0.05-0.23) | 0.192 | 0.04 | 0.13 (-0.02-0.28) | 0.089 | 0.04 |
| MADRS | **0.37 (0.32-0.42)** | **<.001** | 0.06 | **0.37 (0.32-0.42)** | **<.001** | 0.06 |
| History of psychosis | -0.27 (-0.65-0.12) | 0.174 | 0.18 | -0.25 (-0.65-0.15) | 0.219 | 0.24 |
| Antidepressants | **0.53 (0.18-0.89)** | **0.003** | 0.05 | **0.53 (0.18-0.88)** | **0.003** | 0.04 |
| Lithium | -0.27 (-0.6-0.07) | 0.118 | 0.06 | -0.27 (-0.6-0.06) | 0.113 | 0.03 |
| Antipsychotics | **0.53 (0.16-0.9)** | **0.006** | 0.06 | **0.49 (0.12-0.87)** | **0.01** | 0.06 |
| Anxiolytics | **0.46 (0.09-0.84)** | **0.015** | 0.04 | **0.47 (0.1-0.85)** | **0.013** | 0.04 |
| Antiparkisonian | 0.85 (-0.41-2.11) | 0.189 | 0.02 | 0.83 (-0.43-2.08) | 0.196 | 0.01 |
| Education level | -0.05 (-0.11-0.01) | 0.107 | 0.04 | -0.05 (-0.11-0.01) | 0.107 | 0.05 |
| Age at first episode | 0 (-0.02-0.02) | 0.924 | 0.07 | 0 (-0.02-0.02) | 0.936 | 0.10 |
| Number of manic episode | -0.07 (-0.15-0.01) | 0.099 | 0.09 | -0.07 (-0.15-0.01) | 0.09 | 0.12 |
| Number of depressive episode | 0.02 (-0.01-0.05) | 0.185 | 0.22 | 0.02 (-0.01-0.05) | 0.148 | 0.22 |
| CGI | **0.15 (0.03-0.27)** | **0.013** | 0.14 | **0.15 (0.03-0.26)** | **0.012** | 0.08 |

[1] R2 = 0.211; adj R2 = 0.204; AIC = 7096
[2] R2 = 0.212; adj R2 = 0.205; AIC = 7088

| | ABC (sum)[1] | | | ABC (max)[2] | | |
|---|---|---|---|---|---|---|
| Var | Beta (95% CI) | p-value | fmi | Beta (95% CI) | p-value | fmi |
| Scale | 1.18 (0.99-1.4) | 0.058 | 0.04 | 1.17 (0.95-1.43) | 0.131 | 0.05 |
| MADRS | **1.45 (1.38-1.52)** | **<.001** | 0.06 | **1.45 (1.38-1.52)** | **<.001** | 0.06 |
| History of psychosis | 0.76 (0.52-1.12) | 0.166 | 0.18 | 0.77 (0.52-1.15) | 0.201 | 0.24 |
| Antidepressants | **1.75 (1.24-2.48)** | **0.002** | 0.05 | **1.77 (1.25-2.49)** | **0.001** | 0.04 |
| Lithium | 0.77 (0.55-1.08) | 0.131 | 0.06 | 0.77 (0.55-1.07) | 0.118 | 0.03 |
| Antipsychotics | **1.9 (1.36-2.65)** | **<.001** | 0.06 | **1.9 (1.37-2.65)** | **<.001** | 0.05 |
| Anxiolytics | 1.3 (0.85-2) | 0.228 | 0.05 | 1.36 (0.89-2.08) | 0.16 | 0.03 |
| Antiparkisonian | 1.66 (0.44-6.3) | 0.454 | 0.02 | 1.74 (0.45-6.69) | 0.418 | 0.02 |
| Education level | 0.95 (0.9-1.01) | 0.111 | 0.04 | 0.95 (0.9-1.01) | 0.118 | 0.05 |
| Age at first episode | 1 (0.98-1.02) | 0.99 | 0.07 | 1 (0.98-1.02) | 0.984 | 0.10 |
| Number of manic episode | 0.93 (0.86-1.01) | 0.093 | 0.09 | 0.93 (0.86-1.01) | 0.088 | 0.12 |
| Number of depressive episode | 1.02 (0.99-1.05) | 0.236 | 0.23 | 1.02 (0.99-1.05) | 0.187 | 0.22 |
| CGI | **1.16 (1.03-1.31)** | **0.013** | 0.14 | **1.16 (1.03-1.3)** | **0.011** | 0.08 |

[1] R2 = 0.212; adj R2 = 0.205; AIC = 7094
[2] R2 = 0.212; adj R2 = 0.205; AIC = 7088

| | ACB (sum)[1] | | | ACB (max)[2] | | |
|---|---|---|---|---|---|---|
| Var | Beta (95% CI) | p-value | fmi | Beta (95% CI) | p-value | fmi |
| Scale | 0.13 (0-0.26) | 0.051 | 0.04 | **0.22 (0.06-0.38)** | **0.006** | 0.06 |
| MADRS | **0.37 (0.32-0.42)** | **<.001** | 0.06 | **0.37 (0.32-0.42)** | **<.001** | 0.06 |
| History of psychosis | -0.27 (-0.65-0.12) | 0.172 | 0.18 | -0.24 (-0.64-0.15) | 0.228 | 0.24 |
| Antidepressants | 0.5 (0.14-0.86) | 0.006 | 0.05 | 0.5 (0.15-0.85) | 0.005 | 0.04 |
| Lithium | -0.25 (-0.59-0.08) | 0.138 | 0.06 | -0.25 (-0.58-0.09) | 0.146 | 0.03 |
| Antipsychotics | **0.38 (-0.05-0.81)** | **0.083** | 0.06 | **0.28 (-0.14-0.7)** | **0.192** | 0.07 |
| Anxiolytics | **0.4 (0.01-0.78)** | **0.042** | 0.05 | **0.42 (0.05-0.79)** | **0.028** | 0.04 |
| Antiparkisonian | 0.86 (-0.39-2.11) | 0.179 | 0.02 | 0.91 (-0.33-2.16) | 0.149 | 0.01 |
| Education level | -0.05 (-0.11-0.01) | 0.105 | 0.04 | -0.05 (-0.11-0.01) | 0.116 | 0.05 |
| Age at first episode | 0 (-0.02-0.02) | 0.997 | 0.08 | 0 (-0.02-0.02) | 0.986 | 0.10 |
| Number of manic episode | -0.07 (-0.15-0.01) | 0.105 | 0.09 | -0.07 (-0.15-0.01) | 0.097 | 0.11 |
| Number of depressive episode | 0.02 (-0.01-0.05) | 0.205 | 0.22 | 0.02 (-0.01-0.05) | 0.174 | 0.22 |
| CGI | **0.15 (0.03-0.27)** | **0.014** | 0.13 | **0.14 (0.03-0.26)** | **0.013** | 0.08 |

[1] R2 = 0.212; adj R2 = 0.205; AIC = 7094
[2] R2 = 0.215; adj R2 = 0.208; AIC = 7083

| | ADS (sum)[1] | | | ADS (max)[2] | | |
|---|---|---|---|---|---|---|
| Var | Beta (95% CI) | p-value | fmi | Beta (95% CI) | p-value | fmi |
| Scale | 0.09 (-0.04-0.21) | 0.181 | 0.03 | 0.16 (-0.01-0.33) | 0.066 | 0.05 |
| MADRS | **0.37 (0.32-0.42)** | **<.001** | 0.06 | **0.37 (0.32-0.42)** | **<.001** | 0.06 |
| History of psychosis | -0.26 (-0.65-0.12) | 0.179 | 0.18 | -0.25 (-0.64-0.15) | 0.224 | 0.24 |
| Antidepressants | **0.53 (0.17-0.89)** | **0.004** | 0.05 | **0.54 (0.19-0.89)** | **0.002** | 0.04 |
| Lithium | -0.33 (-0.68-0.02) | 0.062 | 0.06 | -0.33 (-0.67-0.01) | 0.055 | 0.03 |
| Antipsychotics | **0.51 (0.13-0.9)** | **0.009** | 0.06 | **0.46 (0.07-0.84)** | **0.02** | 0.06 |
| Anxiolytics | **0.46 (0.08-0.83)** | **0.017** | 0.04 | **0.47 (0.1-0.85)** | **0.013** | 0.04 |
| Antiparkisonian | 0.88 (-0.38-2.13) | 0.171 | 0.02 | 0.89 (-0.36-2.13) | 0.162 | 0.01 |
| Education level | -0.05 (-0.11-0.01) | 0.103 | 0.04 | -0.05 (-0.11-0.01) | 0.103 | 0.05 |
| Age at first episode | 0 (-0.02-0.02) | 0.989 | 0.07 | 0 (-0.02-0.02) | 0.988 | 0.10 |
| Number of manic episode | -0.07 (-0.15-0.01) | 0.098 | 0.09 | -0.07 (-0.15-0.01) | 0.092 | 0.12 |
| Number of depressive episode | 0.02 (-0.01-0.05) | 0.185 | 0.22 | 0.02 (-0.01-0.05) | 0.16 | 0.22 |
| CGI | **0.15 (0.03-0.27)** | **0.014** | 0.13 | **0.15 (0.03-0.26)** | **0.013** | 0.08 |

[1] R2 = 0.211; adj R2 = 0.204; AIC = 7096
[2] R2 = 0.213; adj R2 = 0.205; AIC = 7087

| | AEC (sum)[1] | | | AEC (max)[2] | | |
|---|---|---|---|---|---|---|
| Var | Beta (95% CI) | p-value | fmi | Beta (95% CI) | p-value | fmi |
| Scale | 0.15 (-0.02-0.31) | 0.076 | 0.04 | **0.29 (0.07-0.5)** | **0.008** | 0.05 |
| MADRS | **0.37 (0.32-0.42)** | **<.001** | 0.06 | **0.37 (0.32-0.42)** | **<.001** | 0.06 |
| History of psychosis | -0.27 (-0.65-0.12) | 0.172 | 0.18 | -0.25 (-0.64-0.15) | 0.223 | 0.24 |
| Antidepressants | **0.5 (0.14-0.86)** | **0.006** | 0.05 | **0.5 (0.15-0.85)** | **0.005** | 0.04 |
| Lithium | **-0.4 (-0.77--0.03)** | **0.033** | 0.06 | **-0.41 (-0.75--0.06)** | **0.021** | 0.04 |
| Antipsychotics | **0.44 (0.04-0.84)** | **0.032** | 0.05 | 0.38 (-0.01-0.76) | 0.054 | 0.06 |
| Anxiolytics | **0.4 (0.01-0.78)** | **0.043** | 0.05 | **0.4 (0.02-0.77)** | **0.039** | 0.04 |
| Antiparkisonian | 0.84 (-0.42-2.09) | 0.192 | 0.02 | 0.79 (-0.46-2.03) | 0.216 | 0.01 |
| Education level | -0.05 (-0.11-0.01) | 0.099 | 0.04 | -0.05 (-0.11-0.01) | 0.104 | 0.05 |
| Age at first episode | 0 (-0.02-0.02) | 0.991 | 0.08 | 0 (-0.02-0.02) | 0.966 | 0.10 |
| Number of manic episode | -0.07 (-0.15-0.01) | 0.096 | 0.09 | -0.07 (-0.15-0.01) | 0.091 | 0.12 |
| Number of depressive episode | 0.02 (-0.01-0.05) | 0.19 | 0.22 | 0.02 (-0.01-0.05) | 0.168 | 0.22 |
| CGI | **0.15 (0.03-0.27)** | **0.013** | 0.13 | **0.15 (0.03-0.26)** | **0.012** | 0.08 |

[1] R2 = 0.212; adj R2 = 0.204; AIC = 7095
[2] R2 = 0.215; adj R2 = 0.207; AIC = 7084

| | AIS (sum)[1] | | | AIS (max)[2] | | |
|---|---|---|---|---|---|---|
| Var | Beta (95% CI) | p-value | fmi | Beta (95% CI) | p-value | fmi |
| Scale | **0.21 (0.05-0.37)** | **0.012** | 0.05 | **0.32 (0.06-0.58)** | **0.017** | 0.05 |
| MADRS | **0.37 (0.32-0.42)** | **<.001** | 0.06 | **0.37 (0.32-0.42)** | **<.001** | 0.06 |
| History of psychosis | -0.26 (-0.64-0.12) | 0.181 | 0.18 | -0.25 (-0.65-0.14) | 0.206 | 0.23 |
| Antidepressants | 0.34 (-0.05-0.74) | 0.091 | 0.05 | **0.51 (0.17-0.86)** | **0.004** | 0.04 |
| Lithium | **-0.39 (-0.73--0.04)** | **0.03** | 0.06 | -0.29 (-0.62-0.05) | 0.091 | 0.03 |
| Antipsychotics | 0.3 (-0.13-0.73) | 0.169 | 0.06 | **0.41 (0.03-0.79)** | **0.036** | 0.05 |
| Anxiolytics | 0.24 (-0.18-0.65) | 0.261 | 0.04 | **0.4 (0.02-0.77)** | **0.04** | 0.04 |
| Antiparkisonian | 0.29 (-1.06-1.64) | 0.673 | 0.03 | 0.56 (-0.73-1.84) | 0.396 | 0.01 |
| Education level | -0.05 (-0.11-0.01) | 0.11 | 0.04 | -0.05 (-0.11-0.01) | 0.122 | 0.05 |
| Age at first episode | 0 (-0.02-0.02) | 0.956 | 0.07 | 0 (-0.02-0.02) | 0.967 | 0.10 |
| Number of manic episode | -0.07 (-0.15-0.01) | 0.083 | 0.10 | -0.07 (-0.15-0.01) | 0.087 | 0.12 |
| Number of depressive episode | 0.02 (-0.01-0.05) | 0.212 | 0.23 | 0.02 (-0.01-0.05) | 0.184 | 0.22 |
| CGI | **0.15 (0.03-0.26)** | **0.016** | 0.14 | **0.15 (0.03-0.26)** | **0.012** | 0.08 |

[1] R2 = 0.214; adj R2 = 0.206; AIC = 7091
[2] R2 = 0.214; adj R2 = 0.207; AIC = 7085

| | ALS (sum)[1] | | |
|---|---|---|---|
| Var | Beta (95% CI) | p-value | fmi |
| Scale | 0.11 (-0.19-0.4) | 0.472 | 0.08 |
| MADRS | **0.37 (0.32-0.42)** | **<.001** | 0.06 |
| History of psychosis | -0.28 (-0.66-0.1) | 0.154 | 0.18 |
| Antidepressants | **0.5 (0.07-0.93)** | **0.023** | 0.05 |
| Lithium | -0.38 (-0.83-0.07) | 0.1 | 0.08 |
| Antipsychotics | **0.63 (0.3-0.97)** | **<.001** | 0.06 |
| Anxiolytics | **0.42 (0.03-0.82)** | **0.035** | 0.05 |
| Antiparkisonian | 0.9 (-0.35-2.16) | 0.159 | 0.02 |
| Education level | -0.05 (-0.11-0.01) | 0.104 | 0.04 |
| Age at first episode | 0 (-0.02-0.02) | 0.918 | 0.07 |
| Number of manic episode | -0.07 (-0.15-0.01) | 0.099 | 0.09 |
| Number of depressive episode | 0.02 (-0.01-0.05) | 0.207 | 0.22 |
| CGI | **0.15 (0.03-0.27)** | **0.014** | 0.13 |

[1] R2 = 0.21; adj R2 = 0.203; AIC = 7097

| | ARS (sum)[1] | | | ARS (max)[2] | | |
|---|---|---|---|---|---|---|
| Var | Beta (95% CI) | p-value | fmi | Beta (95% CI) | p-value | fmi |
| Scale | 0.21 (-0.02-0.44) | 0.069 | 0.06 | **0.29 (0.04-0.54)** | **0.021** | 0.05 |
| MADRS | **0.37 (0.32-0.42)** | **<.001** | 0.06 | **0.37 (0.32-0.42)** | **<.001** | 0.06 |
| History of psychosis | -0.26 (-0.64-0.12) | 0.18 | 0.18 | -0.24 (-0.64-0.15) | 0.227 | 0.24 |
| Antidepressants | **0.56 (0.22-0.91)** | **0.001** | 0.05 | **0.57 (0.22-0.91)** | **0.001** | 0.04 |
| Lithium | -0.27 (-0.61-0.07) | 0.115 | 0.06 | -0.27 (-0.6-0.06) | 0.108 | 0.03 |
| Antipsychotics | **0.47 (0.08-0.85)** | **0.018** | 0.06 | **0.41 (0.02-0.8)** | **0.037** | 0.06 |
| Anxiolytics | **0.42 (0.04-0.79)** | **0.031** | 0.05 | **0.41 (0.04-0.79)** | **0.032** | 0.04 |
| Antiparkisonian | 0.99 (-0.26-2.24) | 0.12 | 0.01 | 1 (-0.24-2.24) | 0.115 | 0.01 |
| Education level | -0.05 (-0.11-0.01) | 0.113 | 0.04 | -0.05 (-0.11-0.01) | 0.121 | 0.05 |
| Age at first episode | 0 (-0.02-0.02) | 0.947 | 0.07 | 0 (-0.02-0.02) | 0.942 | 0.10 |
| Number of manic episode | -0.07 (-0.15-0.01) | 0.097 | 0.09 | -0.07 (-0.15-0.01) | 0.088 | 0.12 |
| Number of depressive episode | 0.02 (-0.01-0.05) | 0.179 | 0.22 | 0.02 (-0.01-0.05) | 0.147 | 0.22 |
| CGI | **0.15 (0.03-0.27)** | **0.014** | 0.13 | **0.15 (0.03-0.26)** | **0.012** | 0.08 |

[1] R2 = 0.212; adj R2 = 0.205; AIC = 7094
[2] R2 = 0.214; adj R2 = 0.207; AIC = 7085

| | BADS (sum)[1] | | | BADS (max)[2] | | |
|---|---|---|---|---|---|---|
| Var | Beta (95% CI) | p-value | fmi | Beta (95% CI) | p-value | fmi |
| Scale | **0.14 (0-0.27)** | **0.043** | 0.05 | **0.28 (0.1-0.47)** | **0.003** | 0.05 |
| MADRS | **0.37 (0.32-0.42)** | **<.001** | 0.06 | **0.37 (0.32-0.42)** | **<.001** | 0.06 |
| History of psychosis | -0.26 (-0.64-0.12) | 0.182 | 0.18 | -0.24 (-0.64-0.15) | 0.224 | 0.24 |
| Antidepressants | **0.43 (0.06-0.81)** | **0.025** | 0.05 | **0.47 (0.11-0.82)** | **0.01** | 0.04 |
| Lithium | **-0.37 (-0.71--0.02)** | **0.04** | 0.06 | -0.32 (-0.65-0.01) | 0.057 | 0.03 |
| Antipsychotics | **0.48 (0.11-0.85)** | **0.012** | 0.06 | **0.45 (0.1-0.81)** | **0.012** | 0.06 |
| Anxiolytics | **0.39 (0.01-0.77)** | **0.043** | 0.04 | **0.43 (0.06-0.8)** | **0.024** | 0.04 |
| Antiparkisonian | 0.81 (-0.45-2.06) | 0.21 | 0.02 | 0.82 (-0.42-2.06) | 0.195 | 0.01 |
| Education level | -0.05 (-0.11-0.01) | 0.11 | 0.04 | -0.05 (-0.11-0.01) | 0.106 | 0.05 |
| Age at first episode | 0 (-0.02-0.02) | 0.995 | 0.08 | 0 (-0.02-0.02) | 0.97 | 0.10 |
| Number of manic episode | -0.07 (-0.15-0.01) | 0.083 | 0.10 | -0.08 (-0.16-0.01) | 0.074 | 0.12 |
| Number of depressive episode | 0.02 (-0.01-0.05) | 0.178 | 0.23 | 0.02 (-0.01-0.05) | 0.151 | 0.22 |
| CGI | **0.14 (0.03-0.26)** | **0.017** | 0.13 | **0.14 (0.03-0.26)** | **0.016** | 0.08 |

[1] R2 = 0.212; adj R2 = 0.205; AIC = 7094
[2] R2 = 0.216; adj R2 = 0.209; AIC = 7082

| | CALS (sum)[1] | | | CALS (max)[2] | | |
|---|---|---|---|---|---|---|
| Var | Beta (95% CI) | p-value | fmi | Beta (95% CI) | p-value | fmi |
| Scale | **0.23 (0.07-0.4)** | **0.006** | 0.05 | **0.35 (0.09-0.61)** | **0.008** | 0.06 |
| MADRS | **0.37 (0.32-0.42)** | **<.001** | 0.06 | **0.37 (0.32-0.42)** | **<.001** | 0.06 |
| History of psychosis | -0.26 (-0.65-0.12) | 0.174 | 0.18 | -0.26 (-0.65-0.14) | 0.204 | 0.23 |
| Antidepressants | 0.35 (-0.04-0.74) | 0.077 | 0.05 | **0.5 (0.15-0.85)** | **0.005** | 0.04 |
| Lithium | **-0.41 (-0.76--0.06)** | **0.021** | 0.06 | -0.3 (-0.63-0.03) | 0.074 | 0.03 |
| Antipsychotics | 0.31 (-0.11-0.72) | 0.145 | 0.06 | **0.41 (0.04-0.78)** | **0.031** | 0.06 |
| Anxiolytics | 0.28 (-0.12-0.67) | 0.169 | 0.04 | **0.41 (0.03-0.78)** | **0.032** | 0.04 |
| Antiparkisonian | 0.23 (-1.12-1.58) | 0.741 | 0.02 | 0.48 (-0.81-1.77) | 0.47 | 0.01 |
| Education level | -0.05 (-0.11-0.01) | 0.096 | 0.04 | -0.05 (-0.11-0.01) | 0.104 | 0.05 |
| Age at first episode | 0 (-0.02-0.02) | 0.946 | 0.08 | 0 (-0.02-0.02) | 0.957 | 0.10 |
| Number of manic episode | -0.08 (-0.16-0.01) | 0.072 | 0.10 | -0.07 (-0.16-0.01) | 0.079 | 0.12 |
| Number of depressive episode | 0.02 (-0.01-0.05) | 0.177 | 0.23 | 0.02 (-0.01-0.05) | 0.167 | 0.22 |
| CGI | **0.14 (0.03-0.26)** | **0.017** | 0.13 | **0.14 (0.03-0.26)** | **0.013** | 0.08 |

[1] R2 = 0.214; adj R2 = 0.207; AIC = 7090
[2] R2 = 0.215; adj R2 = 0.208; AIC = 7083

| | CrAS (sum)[1] | | | CrAS (max)[2] | | |
|---|---|---|---|---|---|---|
| Var | Beta (95% CI) | p-value | fmi | Beta (95% CI) | p-value | fmi |
| Scale | 0.09 (-0.11-0.28) | 0.389 | 0.04 | 0.11 (-0.12-0.34) | 0.339 | 0.06 |
| MADRS | **0.37 (0.32-0.42)** | **<.001** | 0.06 | **0.37 (0.32-0.42)** | **<.001** | 0.06 |
| History of psychosis | -0.27 (-0.65-0.11) | 0.166 | 0.18 | -0.25 (-0.65-0.14) | 0.209 | 0.24 |
| Antidepressants | **0.53 (0.16-0.9)** | **0.005** | 0.05 | **0.55 (0.19-0.9)** | **0.003** | 0.05 |
| Lithium | -0.27 (-0.61-0.06) | 0.114 | 0.06 | -0.27 (-0.6-0.06) | 0.109 | 0.03 |
| Antipsychotics | **0.56 (0.17-0.95)** | **0.005** | 0.06 | **0.55 (0.17-0.93)** | **0.005** | 0.07 |
| Anxiolytics | **0.45 (0.07-0.83)** | **0.019** | 0.04 | **0.46 (0.09-0.84)** | **0.015** | 0.04 |
| Antiparkisonian | 0.89 (-0.36-2.15) | 0.164 | 0.02 | 0.89 (-0.36-2.14) | 0.163 | 0.01 |
| Education level | -0.05 (-0.11-0.01) | 0.105 | 0.04 | -0.05 (-0.11-0.01) | 0.109 | 0.05 |
| Age at first episode | 0 (-0.02-0.02) | 0.934 | 0.07 | 0 (-0.02-0.02) | 0.937 | 0.10 |
| Number of manic episode | -0.07 (-0.15-0.01) | 0.108 | 0.09 | -0.07 (-0.15-0.01) | 0.1 | 0.12 |
| Number of depressive episode | 0.02 (-0.01-0.05) | 0.205 | 0.22 | 0.02 (-0.01-0.05) | 0.17 | 0.22 |
| CGI | **0.15 (0.03-0.27)** | **0.014** | 0.13 | **0.15 (0.03-0.26)** | **0.012** | 0.08 |

[1] R2 = 0.21; adj R2 = 0.203; AIC = 7097
[2] R2 = 0.211; adj R2 = 0.204; AIC = 7090

| | DBI-WHO (sum)[1] | | |
|---|---|---|---|
| Var | Beta (95% CI) | p-value | fmi |
| Scale | 0.18 (-0.32-0.69) | 0.473 | 0.04 |
| MADRS | **0.37 (0.32-0.42)** | **<.001** | 0.06 |
| History of psychosis | -0.27 (-0.66-0.11) | 0.161 | 0.18 |
| Antidepressants | **0.52 (0.13-0.92)** | **0.01** | 0.04 |
| Lithium | -0.34 (-0.72-0.05) | 0.085 | 0.06 |
| Antipsychotics | **0.59 (0.22-0.95)** | **0.002** | 0.05 |
| Anxiolytics | **0.43 (0.04-0.81)** | **0.03** | 0.04 |
| Antiparkisonian | 0.91 (-0.34-2.17) | 0.155 | 0.02 |
| Education level | -0.05 (-0.11-0.01) | 0.106 | 0.04 |
| Age at first episode | 0 (-0.02-0.02) | 0.959 | 0.07 |
| Number of manic episode | -0.07 (-0.15-0.01) | 0.089 | 0.09 |
| Number of depressive episode | 0.02 (-0.01-0.05) | 0.208 | 0.22 |
| CGI | **0.15 (0.03-0.27)** | **0.015** | 0.14 |

[1] R2 = 0.21; adj R2 = 0.203; AIC = 7097

| | DRS (sum)[1] | | | DRS (max)[2] | | |
|---|---|---|---|---|---|---|
| Var | Beta (95% CI) | p-value | fmi | Beta (95% CI) | p-value | fmi |
| Scale | 0.08 (-0.03-0.2) | 0.15 | 0.03 | **0.23 (0.06-0.4)** | **0.009** | 0.04 |
| MADRS | **0.37 (0.32-0.42)** | **<.001** | 0.06 | **0.37 (0.32-0.42)** | **<.001** | 0.06 |
| History of psychosis | -0.26 (-0.65-0.12) | 0.176 | 0.18 | -0.25 (-0.65-0.14) | 0.21 | 0.24 |
| Antidepressants | **0.48 (0.1-0.86)** | **0.013** | 0.05 | **0.46 (0.11-0.82)** | **0.011** | 0.04 |
| Lithium | **-0.4 (-0.79--0.02)** | **0.039** | 0.05 | **-0.45 (-0.81--0.09)** | **0.013** | 0.03 |
| Antipsychotics | **0.55 (0.2-0.91)** | **0.002** | 0.06 | **0.54 (0.2-0.88)** | **0.002** | 0.05 |
| Anxiolytics | 0.35 (-0.05-0.76) | 0.087 | 0.04 | **0.39 (0.01-0.77)** | **0.042** | 0.04 |
| Antiparkisonian | 0.85 (-0.41-2.11) | 0.186 | 0.02 | 0.76 (-0.49-2.01) | 0.236 | 0.01 |
| Education level | -0.05 (-0.11-0.01) | 0.104 | 0.04 | -0.05 (-0.11-0.01) | 0.11 | 0.05 |
| Age at first episode | 0 (-0.02-0.02) | 0.981 | 0.07 | 0 (-0.02-0.02) | 0.974 | 0.10 |
| Number of manic episode | -0.07 (-0.15-0.01) | 0.086 | 0.09 | -0.08 (-0.16-0.01) | 0.072 | 0.12 |
| Number of depressive episode | 0.02 (-0.01-0.05) | 0.18 | 0.22 | 0.02 (-0.01-0.05) | 0.154 | 0.22 |
| CGI | **0.15 (0.03-0.27)** | **0.014** | 0.13 | **0.14 (0.03-0.26)** | **0.013** | 0.08 |

[1] R2 = 0.211; adj R2 = 0.204; AIC = 7096
[2] R2 = 0.215; adj R2 = 0.207; AIC = 7084

| | Duran (sum)[1] | | | Duran (max)[2] | | |
|---|---|---|---|---|---|---|
| Var | Beta (95% CI) | p-value | fmi | Beta (95% CI) | p-value | fmi |
| Scale | **0.16 (0.03-0.3)** | **0.015** | 0.03 | **0.31 (0.1-0.52)** | **0.004** | 0.05 |
| MADRS | **0.37 (0.32-0.42)** | **<.001** | 0.06 | **0.37 (0.32-0.42)** | **<.001** | 0.06 |
| History of psychosis | -0.26 (-0.64-0.12) | 0.183 | 0.18 | -0.25 (-0.65-0.14) | 0.213 | 0.24 |
| Antidepressants | **0.47 (0.11-0.83)** | **0.011** | 0.05 | **0.5 (0.16-0.85)** | **0.004** | 0.04 |
| Lithium | **-0.57 (-0.98--0.15)** | **0.007** | 0.06 | **-0.57 (-0.95--0.18)** | **0.004** | 0.04 |
| Antipsychotics | **0.43 (0.06-0.81)** | **0.022** | 0.06 | **0.49 (0.15-0.84)** | **0.005** | 0.06 |
| Anxiolytics | 0.28 (-0.12-0.68) | 0.166 | 0.04 | 0.37 (-0.01-0.75) | 0.055 | 0.04 |
| Antiparkisonian | 0.45 (-0.87-1.76) | 0.504 | 0.02 | 0.52 (-0.76-1.79) | 0.425 | 0.01 |
| Education level | -0.05 (-0.11-0.01) | 0.088 | 0.04 | -0.05 (-0.11-0.01) | 0.092 | 0.05 |
| Age at first episode | 0 (-0.02-0.02) | 0.981 | 0.08 | 0 (-0.02-0.02) | 0.966 | 0.10 |
| Number of manic episode | -0.07 (-0.15-0.01) | 0.087 | 0.09 | -0.07 (-0.15-0.01) | 0.093 | 0.12 |
| Number of depressive episode | 0.02 (-0.01-0.05) | 0.178 | 0.22 | 0.02 (-0.01-0.05) | 0.155 | 0.22 |
| CGI | **0.15 (0.03-0.27)** | **0.014** | 0.13 | **0.15 (0.03-0.26)** | **0.013** | 0.08 |

[1] R2 = 0.213; adj R2 = 0.206; AIC = 7092
[2] R2 = 0.216; adj R2 = 0.208; AIC = 7082

| | GACB (sum)[1] | | | GACB (max)[2] | | |
|---|---|---|---|---|---|---|
| Var | Beta (95% CI) | p-value | fmi | Beta (95% CI) | p-value | fmi |
| Scale | **0.17 (0.02-0.33)** | **0.032** | 0.03 | **0.33 (0.09-0.58)** | **0.008** | 0.05 |
| MADRS | **0.37 (0.32-0.42)** | **<.001** | 0.06 | **0.37 (0.32-0.42)** | **<.001** | 0.06 |
| History of psychosis | -0.27 (-0.65-0.11) | 0.164 | 0.18 | -0.26 (-0.65-0.14) | 0.199 | 0.23 |
| Antidepressants | **0.41 (0.03-0.79)** | **0.035** | 0.05 | **0.5 (0.15-0.85)** | **0.005** | 0.04 |
| Lithium | **-0.38 (-0.73--0.03)** | **0.036** | 0.06 | -0.3 (-0.64-0.03) | 0.073 | 0.03 |
| Antipsychotics | **0.4 (0-0.8)** | **0.049** | 0.06 | **0.43 (0.06-0.8)** | **0.022** | 0.06 |
| Anxiolytics | 0.35 (-0.04-0.74) | 0.077 | 0.04 | **0.42 (0.05-0.79)** | **0.028** | 0.04 |
| Antiparkisonian | 0.72 (-0.55-1.98) | 0.269 | 0.02 | 0.79 (-0.45-2.04) | 0.213 | 0.01 |
| Education level | -0.05 (-0.11-0.01) | 0.103 | 0.04 | -0.05 (-0.11-0.01) | 0.11 | 0.05 |
| Age at first episode | 0 (-0.02-0.02) | 0.98 | 0.08 | 0 (-0.02-0.02) | 0.943 | 0.10 |
| Number of manic episode | -0.07 (-0.16-0.01) | 0.076 | 0.10 | -0.07 (-0.16-0.01) | 0.077 | 0.11 |
| Number of depressive episode | 0.02 (-0.01-0.05) | 0.185 | 0.23 | 0.02 (-0.01-0.05) | 0.165 | 0.22 |
| CGI | **0.15 (0.03-0.26)** | **0.015** | 0.13 | **0.15 (0.03-0.26)** | **0.013** | 0.08 |

[1] R2 = 0.213; adj R2 = 0.205; AIC = 7093
[2] R2 = 0.215; adj R2 = 0.207; AIC = 7084

| | KABS (sum)[1] | | | KABS (max)[2] | | |
|---|---|---|---|---|---|---|
| Var | Beta (95% CI) | p-value | fmi | Beta (95% CI) | p-value | fmi |
| Scale | **0.22 (0.06-0.39)** | **0.008** | 0.05 | **0.3 (0.08-0.51)** | **0.007** | 0.06 |
| MADRS | **0.37 (0.32-0.42)** | **<.001** | 0.06 | **0.37 (0.32-0.42)** | **<.001** | 0.06 |
| History of psychosis | -0.26 (-0.64-0.12) | 0.177 | 0.18 | -0.25 (-0.64-0.15) | 0.219 | 0.24 |
| Antidepressants | 0.37 (-0.01-0.75) | 0.059 | 0.05 | **0.43 (0.06-0.79)** | **0.022** | 0.05 |
| Lithium | -0.26 (-0.59-0.08) | 0.136 | 0.06 | -0.25 (-0.58-0.08) | 0.135 | 0.03 |
| Antipsychotics | 0.26 (-0.18-0.7) | 0.251 | 0.06 | 0.27 (-0.16-0.7) | 0.218 | 0.07 |
| Anxiolytics | 0.31 (-0.08-0.7) | 0.119 | 0.04 | **0.39 (0.01-0.76)** | **0.045** | 0.04 |
| Antiparkisonian | 0.74 (-0.52-2) | 0.25 | 0.02 | 0.83 (-0.41-2.08) | 0.189 | 0.01 |
| Education level | -0.05 (-0.11-0.01) | 0.107 | 0.04 | -0.05 (-0.11-0.01) | 0.117 | 0.05 |
| Age at first episode | 0 (-0.02-0.02) | 0.978 | 0.08 | 0 (-0.02-0.02) | 0.992 | 0.10 |
| Number of manic episode | -0.07 (-0.15-0.01) | 0.093 | 0.09 | -0.07 (-0.15-0.01) | 0.09 | 0.12 |
| Number of depressive episode | 0.02 (-0.01-0.05) | 0.183 | 0.22 | 0.02 (-0.01-0.05) | 0.153 | 0.22 |
| CGI | **0.15 (0.03-0.26)** | **0.015** | 0.13 | **0.14 (0.03-0.26)** | **0.014** | 0.08 |

[1] R2 = 0.214; adj R2 = 0.207; AIC = 7091
[2] R2 = 0.215; adj R2 = 0.208; AIC = 7083

| | mACB1 (sum)[1] | | | mACB1 (max)[2] | | |
|---|---|---|---|---|---|---|
| Var | Beta (95% CI) | p-value | fmi | Beta (95% CI) | p-value | fmi |
| Scale | 0.07 (-0.06-0.21) | 0.285 | 0.04 | 0.13 (-0.03-0.29) | 0.118 | 0.05 |
| MADRS | **0.37 (0.32-0.42)** | **<.001** | 0.06 | **0.37 (0.32-0.42)** | **<.001** | 0.06 |
| History of psychosis | -0.27 (-0.65-0.11) | 0.167 | 0.18 | -0.25 (-0.65-0.15) | 0.215 | 0.24 |
| Antidepressants | **0.52 (0.15-0.89)** | **0.006** | 0.05 | **0.52 (0.16-0.88)** | **0.004** | 0.05 |
| Lithium | -0.26 (-0.6-0.07) | 0.126 | 0.06 | -0.26 (-0.59-0.07) | 0.129 | 0.03 |
| Antipsychotics | **0.5 (0.07-0.93)** | **0.023** | 0.06 | **0.43 (0.01-0.86)** | **0.044** | 0.06 |
| Anxiolytics | **0.46 (0.08-0.83)** | **0.017** | 0.04 | **0.47 (0.1-0.84)** | **0.013** | 0.04 |
| Antiparkisonian | 0.9 (-0.35-2.16) | 0.158 | 0.02 | 0.93 (-0.32-2.17) | 0.144 | 0.01 |
| Education level | -0.05 (-0.11-0.01) | 0.103 | 0.04 | -0.05 (-0.11-0.01) | 0.107 | 0.05 |
| Age at first episode | 0 (-0.02-0.02) | 0.96 | 0.08 | 0 (-0.02-0.02) | 0.965 | 0.10 |
| Number of manic episode | -0.07 (-0.15-0.01) | 0.097 | 0.09 | -0.07 (-0.15-0.01) | 0.088 | 0.12 |
| Number of depressive episode | 0.02 (-0.01-0.05) | 0.199 | 0.22 | 0.02 (-0.01-0.05) | 0.167 | 0.22 |
| CGI | **0.15 (0.03-0.27)** | **0.015** | 0.13 | **0.14 (0.03-0.26)** | **0.014** | 0.08 |

[1] R2 = 0.211; adj R2 = 0.203; AIC = 7097
[2] R2 = 0.212; adj R2 = 0.205; AIC = 7088

| | mACB2 (sum)[1] | | | mACB2 (max)[2] | | |
|---|---|---|---|---|---|---|
| Var | Beta (95% CI) | p-value | fmi | Beta (95% CI) | p-value | fmi |
| Scale | **0.15 (0.02-0.29)** | **0.026** | 0.04 | **0.23 (0.05-0.4)** | **0.01** | 0.06 |
| MADRS | **0.37 (0.32-0.42)** | **<.001** | 0.06 | **0.37 (0.32-0.42)** | **<.001** | 0.06 |
| History of psychosis | -0.26 (-0.64-0.12) | 0.187 | 0.18 | -0.24 (-0.63-0.16) | 0.24 | 0.24 |
| Antidepressants | **0.41 (0.03-0.79)** | **0.034** | 0.05 | **0.48 (0.13-0.83)** | **0.008** | 0.04 |
| Lithium | -0.21 (-0.55-0.13) | 0.224 | 0.06 | -0.22 (-0.55-0.12) | 0.206 | 0.03 |
| Antipsychotics | 0.36 (-0.06-0.78) | 0.095 | 0.06 | 0.35 (-0.06-0.75) | 0.092 | 0.07 |
| Anxiolytics | 0.35 (-0.03-0.74) | 0.073 | 0.04 | **0.43 (0.06-0.8)** | **0.024** | 0.04 |
| Antiparkisonian | 0.79 (-0.46-2.05) | 0.215 | 0.02 | 0.91 (-0.33-2.16) | 0.149 | 0.01 |
| Education level | -0.05 (-0.11-0.01) | 0.108 | 0.04 | -0.05 (-0.11-0.01) | 0.111 | 0.05 |
| Age at first episode | 0 (-0.02-0.02) | 0.996 | 0.08 | 0 (-0.02-0.02) | 0.989 | 0.10 |
| Number of manic episode | -0.07 (-0.15-0.01) | 0.088 | 0.09 | -0.07 (-0.15-0.01) | 0.088 | 0.11 |
| Number of depressive episode | 0.02 (-0.01-0.05) | 0.183 | 0.22 | 0.02 (-0.01-0.05) | 0.154 | 0.22 |
| CGI | **0.15 (0.03-0.26)** | **0.015** | 0.13 | **0.14 (0.03-0.26)** | **0.014** | 0.08 |

[1] R2 = 0.213; adj R2 = 0.206; AIC = 7093
[2] R2 = 0.215; adj R2 = 0.207; AIC = 7084

| | MARANTE (sum)[1] | | | MARANTE (max)[2] | | |
|---|---|---|---|---|---|---|
| Var | Beta (95% CI) | p-value | fmi | Beta (95% CI) | p-value | fmi |
| Scale | 0.09 (-0.06-0.25) | 0.244 | 0.03 | 0.18 (-0.02-0.38) | 0.085 | 0.03 |
| MADRS | **0.37 (0.32-0.42)** | **<.001** | 0.06 | **0.37 (0.32-0.42)** | **<.001** | 0.06 |
| History of psychosis | -0.27 (-0.65-0.11) | 0.171 | 0.18 | -0.26 (-0.65-0.14) | 0.204 | 0.24 |
| Antidepressants | **0.52 (0.15-0.89)** | **0.006** | 0.05 | **0.51 (0.15-0.86)** | **0.006** | 0.04 |
| Lithium | -0.36 (-0.73-0.01) | 0.056 | 0.06 | **-0.36 (-0.71--0.02)** | **0.041** | 0.03 |
| Antipsychotics | **0.55 (0.18-0.92)** | **0.004** | 0.06 | **0.53 (0.17-0.88)** | **0.004** | 0.05 |
| Anxiolytics | **0.41 (0.03-0.8)** | **0.035** | 0.04 | **0.42 (0.05-0.8)** | **0.028** | 0.04 |
| Antiparkisonian | 0.86 (-0.4-2.12) | 0.18 | 0.02 | 0.82 (-0.43-2.07) | 0.198 | 0.01 |
| Education level | -0.05 (-0.11-0.01) | 0.106 | 0.04 | -0.05 (-0.11-0.01) | 0.116 | 0.05 |
| Age at first episode | 0 (-0.02-0.02) | 0.957 | 0.07 | 0 (-0.02-0.02) | 0.964 | 0.10 |
| Number of manic episode | -0.07 (-0.15-0.01) | 0.092 | 0.09 | -0.07 (-0.16-0.01) | 0.083 | 0.12 |
| Number of depressive episode | 0.02 (-0.01-0.05) | 0.191 | 0.22 | 0.02 (-0.01-0.05) | 0.163 | 0.22 |
| CGI | **0.15 (0.03-0.27)** | **0.013** | 0.13 | **0.15 (0.03-0.26)** | **0.011** | 0.08 |

[1] R2 = 0.211; adj R2 = 0.203; AIC = 7097
[2] R2 = 0.212; adj R2 = 0.205; AIC = 7088

| | mARS (sum)[1] | | | mARS (max)[2] | | |
|---|---|---|---|---|---|---|
| Var | Beta (95% CI) | p-value | fmi | Beta (95% CI) | p-value | fmi |
| Scale | 0.18 (-0.03-0.39) | 0.1 | 0.06 | **0.3 (0.06-0.53)** | **0.015** | 0.05 |
| MADRS | **0.37 (0.32-0.42)** | **<.001** | 0.06 | **0.37 (0.32-0.42)** | **<.001** | 0.06 |
| History of psychosis | -0.26 (-0.65-0.12) | 0.178 | 0.18 | -0.24 (-0.64-0.15) | 0.228 | 0.24 |
| Antidepressants | **0.55 (0.2-0.9)** | **0.002** | 0.05 | **0.54 (0.19-0.88)** | **0.002** | 0.04 |
| Lithium | -0.27 (-0.6-0.07) | 0.121 | 0.06 | -0.27 (-0.6-0.06) | 0.113 | 0.03 |
| Antipsychotics | **0.49 (0.11-0.87)** | **0.011** | 0.06 | **0.41 (0.03-0.79)** | **0.035** | 0.06 |
| Anxiolytics | **0.42 (0.04-0.8)** | **0.031** | 0.05 | **0.4 (0.03-0.78)** | **0.036** | 0.04 |
| Antiparkisonian | 0.99 (-0.26-2.24) | 0.119 | 0.01 | 1.01 (-0.23-2.25) | 0.111 | 0.01 |
| Education level | -0.05 (-0.11-0.01) | 0.108 | 0.04 | -0.05 (-0.11-0.01) | 0.112 | 0.05 |
| Age at first episode | 0 (-0.02-0.02) | 0.988 | 0.08 | 0 (-0.02-0.02) | 0.997 | 0.10 |
| Number of manic episode | -0.07 (-0.15-0.01) | 0.093 | 0.09 | -0.07 (-0.15-0.01) | 0.085 | 0.12 |
| Number of depressive episode | 0.02 (-0.01-0.05) | 0.189 | 0.22 | 0.02 (-0.01-0.05) | 0.16 | 0.22 |
| CGI | **0.15 (0.03-0.27)** | **0.014** | 0.13 | **0.15 (0.03-0.26)** | **0.013** | 0.08 |

[1] R2 = 0.211; adj R2 = 0.204; AIC = 7095
[2] R2 = 0.214; adj R2 = 0.207; AIC = 7085

| | Salahudeen (sum)[1] | | | Salahudeen (max)[2] | | |
|---|---|---|---|---|---|---|
| Var | Beta (95% CI) | p-value | fmi | Beta (95% CI) | p-value | fmi |
| Scale | **0.26 (0.07-0.44)** | **0.007** | 0.05 | **0.35 (0.04-0.66)** | **0.025** | 0.06 |
| MADRS | **0.37 (0.32-0.42)** | **<.001** | 0.06 | **0.37 (0.32-0.42)** | **<.001** | 0.06 |
| History of psychosis | -0.28 (-0.66-0.1) | 0.156 | 0.18 | -0.27 (-0.67-0.12) | 0.175 | 0.23 |
| Antidepressants | 0.34 (-0.05-0.73) | 0.086 | 0.05 | **0.51 (0.16-0.86)** | **0.004** | 0.04 |
| Lithium | **-0.43 (-0.79--0.08)** | **0.017** | 0.06 | -0.31 (-0.64-0.02) | 0.066 | 0.03 |
| Antipsychotics | **0.42 (0.04-0.79)** | **0.028** | 0.06 | **0.58 (0.24-0.91)** | **0.001** | 0.05 |
| Anxiolytics | 0.25 (-0.16-0.65) | 0.234 | 0.04 | **0.39 (0.01-0.77)** | **0.046** | 0.04 |
| Antiparkisonian | 0.1 (-1.3-1.5) | 0.886 | 0.03 | 0.3 (-1.06-1.67) | 0.666 | 0.02 |
| Education level | -0.05 (-0.11-0.01) | 0.098 | 0.04 | -0.05 (-0.11-0.01) | 0.111 | 0.05 |
| Age at first episode | 0 (-0.02-0.02) | 0.942 | 0.08 | 0 (-0.02-0.02) | 0.936 | 0.10 |
| Number of manic episode | -0.08 (-0.16-0) | 0.064 | 0.10 | -0.08 (-0.16-0.01) | 0.073 | 0.12 |
| Number of depressive episode | 0.02 (-0.01-0.05) | 0.185 | 0.22 | 0.02 (-0.01-0.05) | 0.177 | 0.22 |
| CGI | **0.14 (0.03-0.26)** | **0.017** | 0.14 | **0.14 (0.03-0.26)** | **0.013** | 0.08 |

[1] R2 = 0.214; adj R2 = 0.207; AIC = 7090
[2] R2 = 0.214; adj R2 = 0.206; AIC = 7086

| | Summers (sum)[1] | | | Summers (max)[2] | | |
|---|---|---|---|---|---|---|
| Var | Beta (95% CI) | p-value | fmi | Beta (95% CI) | p-value | fmi |
| Scale | 0.07 (-0.11-0.24) | 0.452 | 0.23 | 0.06 (-0.11-0.24) | 0.465 | 0.08 |
| MADRS | **0.37 (0.32-0.42)** | **<.001** | 0.06 | **0.37 (0.32-0.42)** | **<.001** | 0.06 |
| History of psychosis | -0.28 (-0.66-0.1) | 0.152 | 0.18 | -0.27 (-0.66-0.13) | 0.188 | 0.24 |
| Antidepressants | **0.59 (0.24-0.94)** | **0.001** | 0.05 | **0.59 (0.25-0.93)** | **0.001** | 0.04 |
| Lithium | -0.28 (-0.62-0.06) | 0.104 | 0.05 | -0.28 (-0.61-0.05) | 0.1 | 0.03 |
| Antipsychotics | **0.64 (0.31-0.97)** | **<.001** | 0.06 | **0.64 (0.31-0.97)** | **<.001** | 0.05 |
| Anxiolytics | **0.43 (0.04-0.81)** | **0.029** | 0.04 | **0.44 (0.05-0.82)** | **0.026** | 0.04 |
| Antiparkisonian | 0.8 (-0.5-2.11) | 0.227 | 0.03 | 0.82 (-0.47-2.11) | 0.215 | 0.01 |
| Education level | -0.05 (-0.11-0.01) | 0.107 | 0.04 | -0.05 (-0.11-0.01) | 0.115 | 0.05 |
| Age at first episode | 0 (-0.02-0.02) | 0.964 | 0.07 | 0 (-0.02-0.02) | 0.967 | 0.10 |
| Number of manic episode | -0.07 (-0.15-0.01) | 0.099 | 0.09 | -0.07 (-0.15-0.01) | 0.089 | 0.12 |
| Number of depressive episode | 0.02 (-0.01-0.05) | 0.2 | 0.22 | 0.02 (-0.01-0.05) | 0.168 | 0.22 |
| CGI | **0.15 (0.03-0.27)** | **0.014** | 0.13 | **0.15 (0.03-0.26)** | **0.012** | 0.08 |

[1] R2 = 0.21; adj R2 = 0.203; AIC = 7097
[2] R2 = 0.211; adj R2 = 0.204; AIC = 7090

**Supplementary Table 5. Multiple logistic regression models of the central anticholinergic side effects subscore with the 22 scales significantly associated with the central subscore in bivariable regression analysis as main predictors.**

**Significant associations are in bold. AAS: Anticholinergic Activity Scale; ABC: Anticholinergic Burden Classification; ACB: Anticholinergic Cognitive Burden Scale; ADS: Anticholinergic Drug Scale; AEC: Anticholinergic Effect on Cognition; AIS: Anticholinergic Impregnation Scale; ARS: Anticholinergic Risk Scale;BADS: Brazilian Anticholinergic Activity Drug Scale; CALS: the CRIDECO Anticholinergic Load Scale; CI: Clinical Index; CrAS: Clinician-rated Anticholinergic Scale; DBI-WHO: Drug Burden Index-WHO version; DRS: Delirogenic Risk Scale; Duran: Durán Scale; GACB: German Anticholinergic Burden Scale; KABS: Korean Anticholinergic Burden Scale; mACB1: modified Anticholinergic Cognitive Burden Scale 1; mACB2: modified Anticholinergic Cognitive Burden scale 2; Marante: Muscarinic Acetylcholinergic Receptor ANTagonist Exposure Scale; mARS: modified Anticholinergic Risk Scale; Salahudeen: Salahudeen's scale; Summers: Summers's scale.**

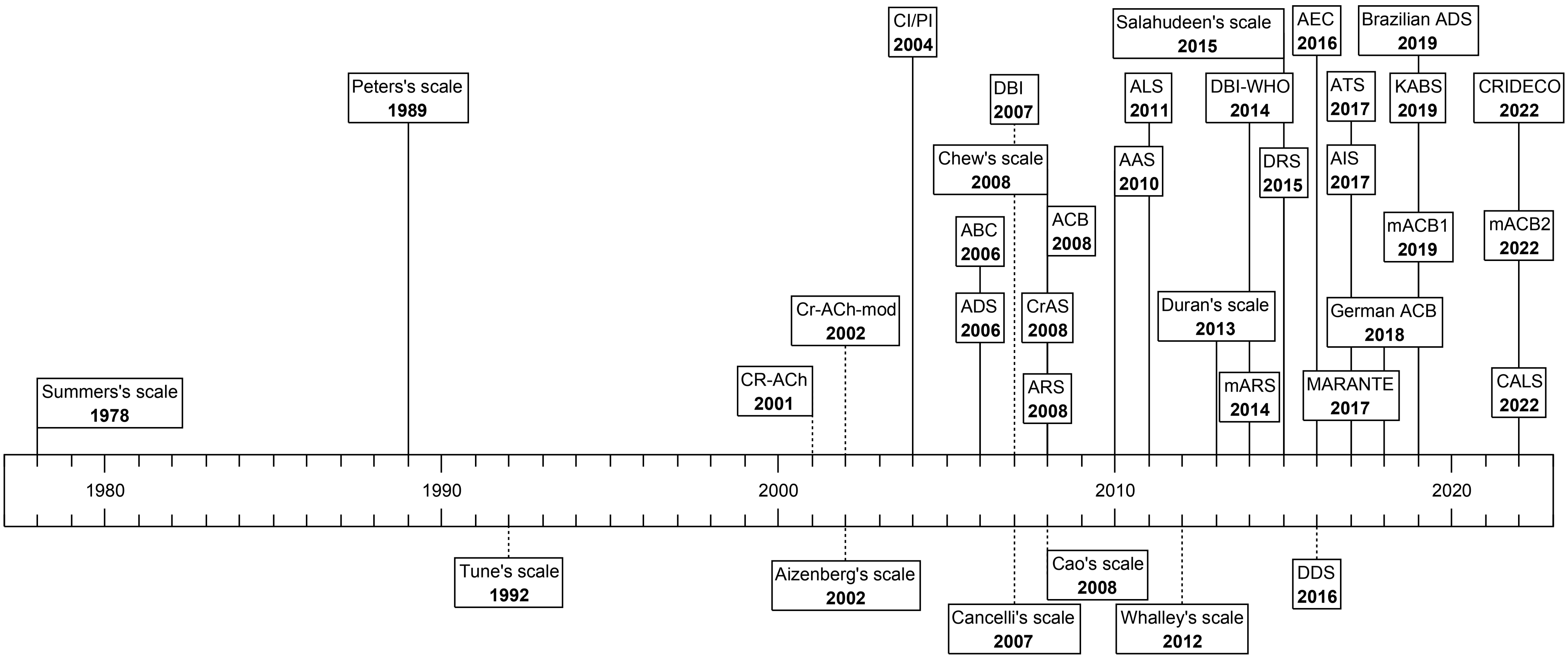


Supplementary Figure 1. Timeline of the 36 identified anticholinergic burden scales.

Dates correspond to the year of creation of the scale. Scales identified with a dotted line were excluded from our study.

**Supplementary Figure 2. Proportion of patients with a non−zero score in each scale (n=2031).**

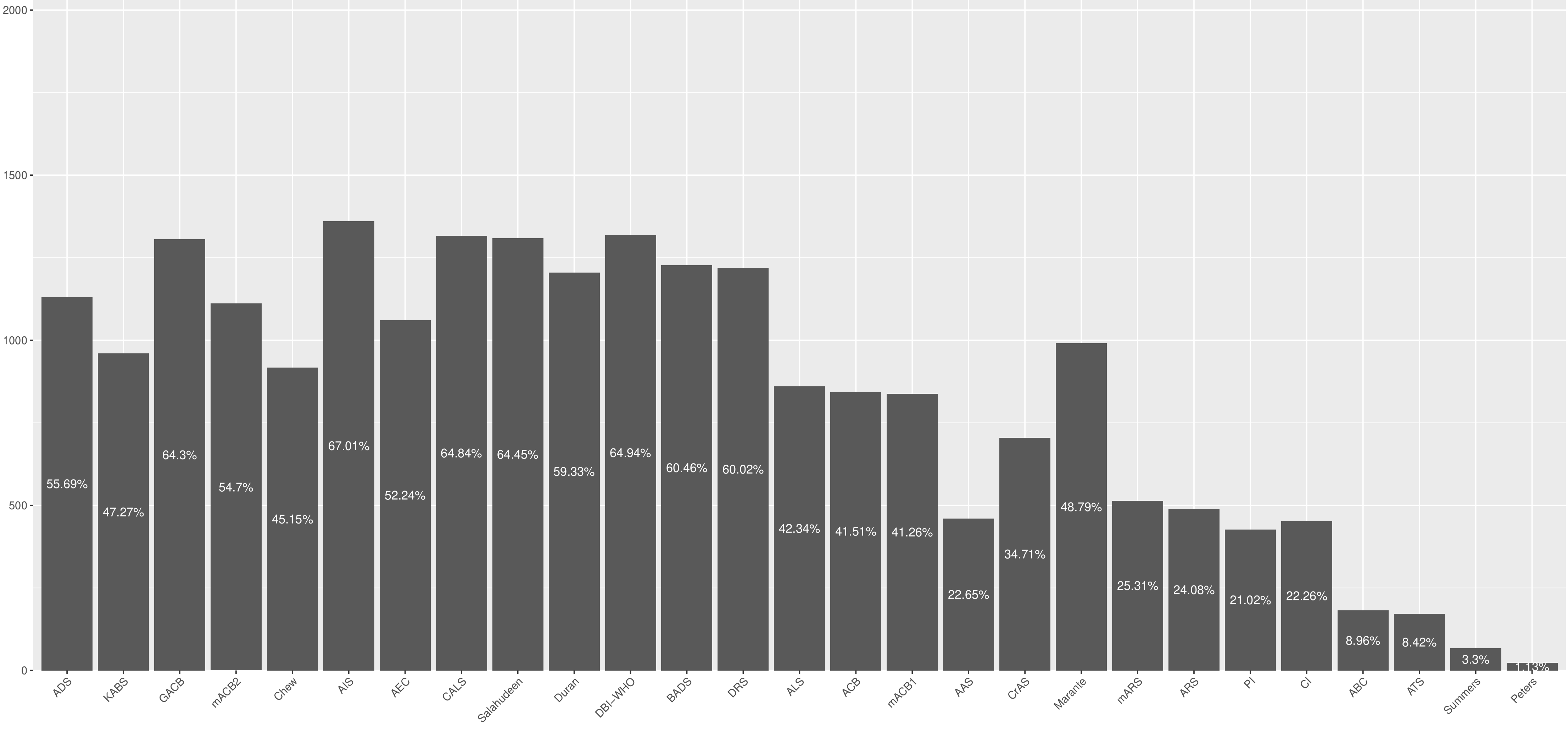


**X−axis is ordered from the scale listing the most drugs (i.e., ADS) to the scale listing the least drugs (i.e., Peters).**

| ABS | SUM: Peripheral subscore Beta (95% CI) | SUM: Peripheral subscore p-value | SUM: Central subscore Beta (95% CI) | SUM: Central subscore p-value | MAX: Peripheral subscore Beta (95% CI) | MAX: Peripheral subscore p-value | MAX: Central subscore Beta (95% CI) | MAX: Central subscore p-value |
|---|---|---|---|---|---|---|---|---|
| ADS | 0.09 (0.01 - 0.18) | 0.029 | 0.20 (0.08 - 0.31) | <.001 | 0.11 (-0.00 - 0.22) | 0.06 | 0.26 (0.10 - 0.42) | 0.001 |
| KABS | 0.22 (0.14 - 0.31) | <.001 | 0.45 (0.32 - 0.57) | <.001 | 0.26 (0.13 - 0.38) | <.001 | 0.54 (0.37 - 0.71) | <.001 |
| GACB | 0.22 (0.12 - 0.31) | <.001 | 0.39 (0.26 - 0.52) | <.001 | 0.30 (0.13 - 0.47) | <.001 | 0.60 (0.36 - 0.84) | <.001 |
| mACB2 | 0.18 (0.10 - 0.25) | <.001 | 0.37 (0.27 - 0.48) | <.001 | 0.19 (0.08 - 0.30) | <.001 | 0.44 (0.29 - 0.59) | <.001 |
| Chew | 0.13 (-0.00 - 0.27) | 0.055 | 0.13 (-0.06 - 0.32) | 0.192 | 0.17 (-0.01 - 0.36) | 0.061 | 0.20 (-0.05 - 0.46) | 0.116 |
| AIS | 0.24 (0.16 - 0.32) | <.001 | 0.38 (0.27 - 0.49) | <.001 | 0.31 (0.15 - 0.48) | <.001 | 0.60 (0.36 - 0.83) | <.001 |
| AEC | 0.15 (0.05 - 0.25) | 0.004 | 0.26 (0.13 - 0.40) | <.001 | 0.20 (0.06 - 0.34) | 0.005 | 0.41 (0.21 - 0.60) | <.001 |
| CALS | 0.22 (0.14 - 0.31) | <.001 | 0.40 (0.28 - 0.52) | <.001 | 0.31 (0.14 - 0.47) | <.001 | 0.61 (0.38 - 0.85) | <.001 |
| Salahudeen | 0.29 (0.19 - 0.39) | <.001 | 0.46 (0.32 - 0.60) | <.001 | 0.46 (0.25 - 0.67) | <.001 | 0.71 (0.42 - 1.00) | <.001 |
| Duran | 0.18 (0.11 - 0.26) | <.001 | 0.20 (0.09 - 0.30) | <.001 | 0.28 (0.15 - 0.42) | <.001 | 0.28 (0.09 - 0.47) | 0.004 |
| DBI-WHO | 0.74 (0.44 - 1.03) | <.001 | 0.92 (0.51 - 1.34) | <.001 | 0.92 (0.23 - 1.61) | 0.009 | 0.83 (-0.13 - 1.80) | 0.092 |
| BADS | 0.20 (0.11 - 0.28) | <.001 | 0.31 (0.19 - 0.43) | <.001 | 0.25 (0.12 - 0.38) | <.001 | 0.47 (0.28 - 0.66) | <.001 |
| DRS | 0.14 (0.07 - 0.21) | <.001 | 0.20 (0.10 - 0.29) | <.001 | 0.23 (0.11 - 0.35) | <.001 | 0.31 (0.14 - 0.48) | <.001 |
| ALS | 0.30 (0.14 - 0.45) | <.001 | 0.23 (0.02 - 0.44) | 0.033 | 0.26 (0.06 - 0.47) | 0.013 | 0.16 (-0.13 - 0.45) | 0.288 |
| ACB | 0.16 (0.09 - 0.24) | <.001 | 0.31 (0.21 - 0.42) | <.001 | 0.18 (0.08 - 0.27) | <.001 | 0.38 (0.24 - 0.51) | <.001 |
| mACB1 | 0.11 (0.03 - 0.19) | 0.006 | 0.27 (0.16 - 0.37) | <.001 | 0.11 (0.02 - 0.21) | 0.022 | 0.30 (0.17 - 0.44) | <.001 |
| AAS | 0.09 (-0.00 - 0.19) | 0.051 | 0.25 (0.12 - 0.38) | <.001 | 0.09 (-0.01 - 0.20) | 0.074 | 0.28 (0.14 - 0.43) | <.001 |
| CrAS | 0.24 (0.11 - 0.36) | <.001 | 0.45 (0.28 - 0.63) | <.001 | 0.25 (0.10 - 0.40) | 0.001 | 0.47 (0.26 - 0.68) | <.001 |
| Marante | 0.18 (0.08 - 0.28) | <.001 | 0.26 (0.12 - 0.39) | <.001 | 0.26 (0.12 - 0.40) | <.001 | 0.37 (0.17 - 0.57) | <.001 |
| mARS | 0.28 (0.13 - 0.42) | <.001 | 0.44 (0.24 - 0.64) | <.001 | 0.33 (0.16 - 0.49) | <.001 | 0.55 (0.33 - 0.77) | <.001 |
| ARS | 0.29 (0.14 - 0.44) | <.001 | 0.48 (0.26 - 0.69) | <.001 | 0.31 (0.15 - 0.48) | <.001 | 0.56 (0.33 - 0.80) | <.001 |
| PI | 0.03 (-0.02 - 0.07) | 0.307 | 0.03 (-0.04 - 0.10) | 0.388 | 0.02 (-0.02 - 0.07) | 0.335 | 0.03 (-0.04 - 0.10) | 0.407 |
| CI | 0.14 (-0.04 - 0.32) | 0.139 | 0.23 (-0.03 - 0.48) | 0.082 | 0.12 (-0.06 - 0.31) | 0.202 | 0.23 (-0.03 - 0.49) | 0.085 |
| ABC | 0.33 (0.22 - 0.44) | <.001 | 0.42 (0.27 - 0.58) | <.001 | 0.36 (0.23 - 0.49) | <.001 | 0.48 (0.30 - 0.66) | <.001 |
| ATS | 0.05 (-0.05 - 0.15) | 0.297 | 0.12 (-0.02 - 0.26) | 0.086 | 0.05 (-0.06 - 0.15) | 0.385 | 0.10 (-0.04 - 0.25) | 0.163 |
| Summers | 0.13 (0.01 - 0.25) | 0.042 | 0.30 (0.12 - 0.47) | <.001 | 0.13 (0.01 - 0.26) | 0.041 | 0.29 (0.12 - 0.47) | 0.001 |
| Peters | -0.04 (-0.46 - 0.37) | 0.84 | 0.34 (-0.24 - 0.92) | 0.25 | -0.09 (-0.53 - 0.35) | 0.692 | 0.30 (-0.32 - 0.91) | 0.346 |

**Supplementary Figure 3. Bivariable logistic regression models of peripheral and central anticholinergic side effects subscores with the scales as predictor.**

**Significant (p<.05) Odds Ratio are in red. AAS: Anticholinergic Activity Scale; ABC: Anticholinergic Burden Classification; ACB: Anticholinergic Cognitive Burden Scale; ADS: Anticholinergic Drug Scale; AEC: Anticholinergic Effect on Cognition; AIS: Anticholinergic Impregnation Scale; ALS: Anticholinergic Loading Scale; ARS: Anticholinergic Risk Scale; ATS: Anticholinergic Toxicity Scale; BADS: Brazilian Anticholinergic Activity Drug Scale; CALS: the CRIDECO Anticholinergic Load Scale; Chew: Chew's scale; CI: Clinical Index; CrAS: Clinician-rated Anticholinergic Scale; DBI-WHO: Drug Burden Index-WHO version; DRS: Delirogenic Risk Scale; Durán: Durán Scale; GACB: German Anticholinergic Burden Scale; KABS: Korean Anticholinergic Burden Scale; mACB1: modified Anticholinergic Cognitive Burden Scale 1; mACB2: modified Anticholinergic Cognitive Burden scale 2; Marante: Muscarinic Acetylcholinergic Receptor ANTagonist Exposure Scale; mARS: modified Anticholinergic Risk Scale; Peters: Peters's scale; PI: Pharmacological Index; Salahudeen: Salahudeen's scale; Summer: Summers's scale.**
